# Does blood pressure level matter more in individuals with higher genetic risk of dementia? A polygenic interaction study in the UK Biobank

**DOI:** 10.1101/2025.07.14.25331521

**Authors:** Phazha Bothongo, Ryan Mattick, Victoria Taylor-Bateman, Patrick G. Kehoe, Yoav Ben-Shlomo, Emma L. Anderson

## Abstract

**Background & Aims:** The causal role of blood pressure (BP) in dementia remains debated. BP may act as an effect modifier; whereby elevated pressure accelerates cognitive decline in the presence of existing neurodegenerative or cerebrovascular pathology, through vascular injury and impaired cerebral homeostasis. We examine whether genetic liability to dementia interacts with BP to influence risk of dementia diagnosis.

**Methods:** In 334,759 UK Biobank participants, we calculated weighted polygenic risk scores (PRSs) for Alzheimer’s disease (AD), vascular dementia (VaD), white matter hyperintensity (WMH) volume (a neuroimaging marker of cerebral small vessel disease), systolic (SBP), and diastolic (DBP) blood pressure. Logistic regression models examined main effects of the BP PRSs on dementia outcomes, and tested interactions between genetic liability to dementia and SBP/DBP levels to see whether they affect risk of dementia diagnoses.

**Results:** The SBP PRS increased dementia risk across all subtypes: AD (OR = 1.04, 95%CI: 1.01–1.08), VaD (OR = 1.06, 95%CI: 1.00–1.11), and all-cause dementia (OR = 1.05, 95%CI: 1.03–1.08). The DBP PRS showed little evidence of association with any dementia outcome. There was little evidence of multiplicative interaction between BP and genetic liability to dementia. Associations of the SBP PRS with dementia risk were consistent across high and low dementia genetic liability groups. For example, for all-cause dementia, the SBP PRS showed similar associations across both high (OR = 1.05, 95% CI: 1.02–1.07) and low (OR = 1.05, 95% CI: 1.02–1.08) AD genetic liability groups (interaction P value = 0.467). Similarly, no strong evidence of interaction was observed between the DBP PRS and dementia genetic liability.

**Conclusions:** Our study supports elevated SBP as an independent risk factor for dementia across subtypes, with little evidence that this effect is modified by genetic predisposition to Alzheimer’s disease or vascular dementia. Our findings reinforce the importance of population-wide strategies to lower SBP as a means of reducing dementia risk at the population level.

## Introduction

Dementia affects over 55 million individuals globally, with projections exceeding 150 million by 2050, driven largely by population ageing (1). It imposes devastating personal costs and a major economic burden, with global costs exceeding $1.3 trillion annually (2). Alzheimer’s disease (AD) and vascular dementia (VaD) are the two most common causes, often co-occurring as mixed pathology in older adults (3). Despite their public health importance, modifiable risk factors and underlying mechanisms for AD and VaD remain incompletely understood.

Elevated blood pressure (BP) is one of the most consistently implicated cardiovascular risk factors for dementia in observational studies (4–8). Proposed mechanisms differ by dementia subtype. In AD, elevated BP may exacerbate cerebral beta-amyloid (Aβ) deposition and promote vascular dysfunction, contributing to cerebral amyloid angiopathy (CAA), impaired Aβ clearance, and blood–brain barrier breakdown (9,10). Autopsy studies also reveal disruptions in the brain renin-angiotensin system (RAS), which normally regulates BP, with links to both AD pathology and increased Aβ and tau burden (11–13). In VaD, elevated BP is a well-established contributor to small vessel disease, lacunar infarcts, and chronic cerebral hypoperfusion - key mechanisms underlying vascular cognitive impairment (6,14).

Causal evidence linking elevated BP to dementia varies by subtype. Recent meta-analyses show mixed findings for AD, with systolic blood pressure (SBP) >140 mmHg linked to 18% higher risk (95% CI: 1.02–1.35), but limited evidence for diastolic blood pressure (DBP) (15). For VaD, more consistent dose-response relationships have been demonstrated. For example, a nationwide Korean study of 4.5 million adults found 23% increased VaD risk for SBP ≥160 mmHg, without protective effects at lower pressures, while DBP ≥90mmHg showed a 2-37% increased risk and DBP <80mmHg reduced risk by up to 13% (7).

Mendelian randomisation (MR) studies, which use genetic variants as unconfounded proxies for exposures, underscore this subtype distinction. Several MR studies report little causal evidence linking BP to AD (16,17), with some even suggesting protective effects of higher SBP (18,19), contradicting observational findings and World Health Organisation (WHO) guidelines (16). Conversely, limited MR research on VaD indicates stronger causal relationships; one recent study reported higher SBP as a significant risk factor for VaD (OR: 1.56, 95%CI, 1.25--1.93), but not AD (OR: 1.10, 95%CI, 0.95–1.26) (17).

Randomized controlled trials (RCTs) of BP-lowering medication for dementia prevention have also produced inconsistent results. The SPRINT-MIND trial found intensive BP reduction (<120LmmHg) decreased mild cognitive impairment by 19% but did not significantly reduce dementia onset (20,21). One individual participant data meta-analysis of 17 studies (34,519 older adults) did find a benefit of antihypertensive medication in reducing dementia risk (22). However, systematic reviews of antihypertensive RCTs generally report minimal or null effects on AD and VaD risk (23,24), often based on post-hoc analyses where cognitive outcomes were not primary endpoints.

These inconsistencies suggest that the effect of BP on dementia may depend critically on the underlying neuropathological context. Rather than operating uniformly, BP may interact with pre-existing neurodegenerative or cerebrovascular vulnerability, to accelerate vascular injury, neuroinflammation, and cerebral dysfunction. To explore this, we investigated whether genetic liability to dementia modifies the association between BP and clinical diagnoses of AD, VaD, and all-cause dementia, where BP is instrumented by polygenic risk scores (PRSs) for both SBP and DBP, to minimize potential bias due to confounding and reverse causation.

## Methods

### Study participants

Our study analysed data from the UK Biobank, a large-scale prospective cohort of approximately 500,000 participants aged 40 to 69 years at recruitment between 2006 and 2010 (25). The cohort provides extensive biological, lifestyle, and health data with long-term follow-up. Eligibility criteria included available genotype data, dementia outcome information, and complete demographic data. We excluded participants based on standard quality control metrics: sex discordance, putative sex chromosome aneuploidy, extreme heterozygosity, non-white British ancestry, relatedness to other participants, or consent withdrawal.

Following these exclusions, 334,759 individuals remained for analysis (Figure 1).

**Figure 1:**
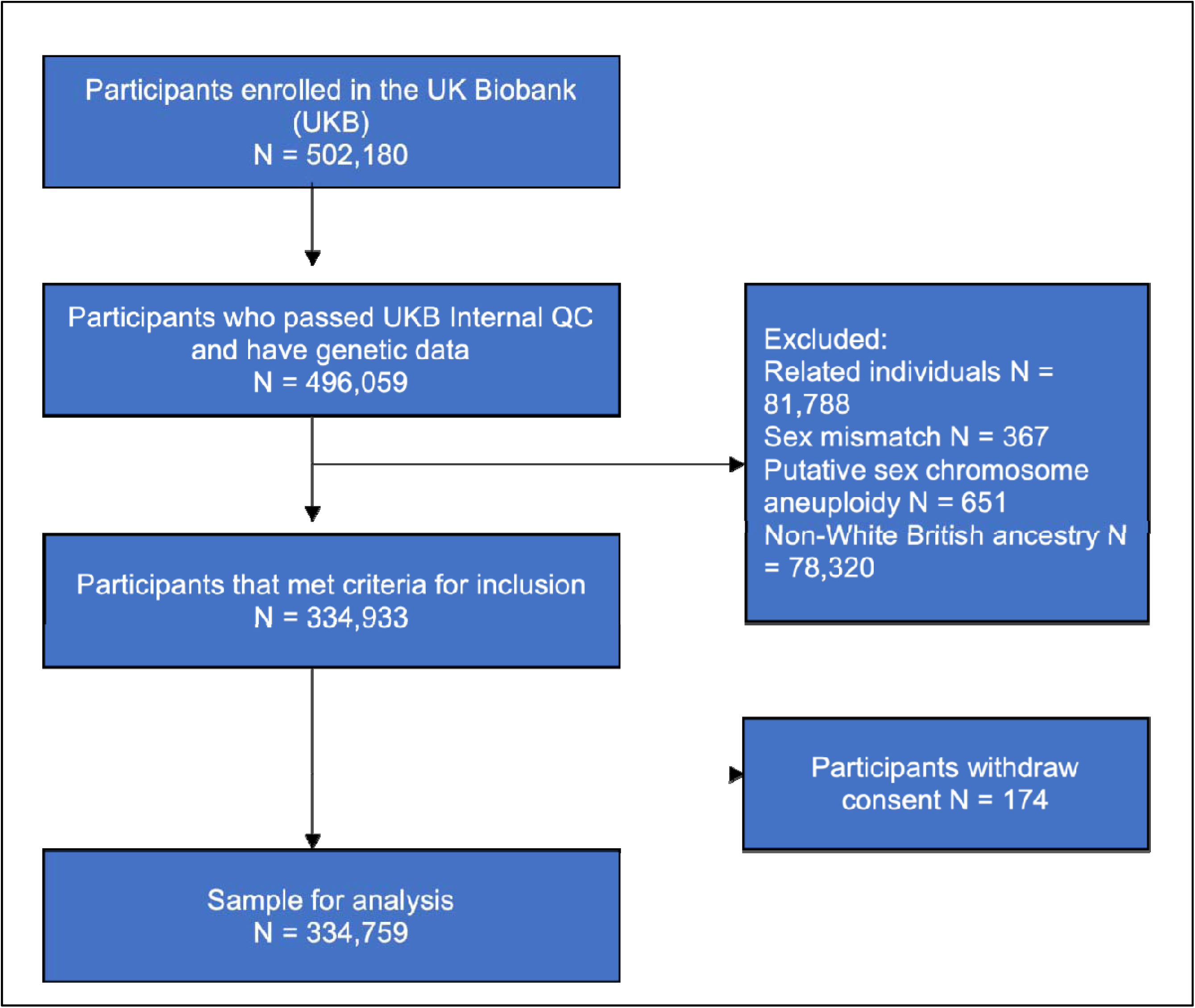
Participant flow chart.

### Genetic data

We used genotyping data in the UK Biobank (described previously) (26) to construct weighted PRSs, using summary statistics from large-scale genome-wide association studies (GWAS). All analyses were restricted to European ancestry participants to minimize population stratification confounding. For AD, we used the most recent two-stage GWAS by Bellenguez et al. (2022), incorporating 75 associated variants (27). The Bellenguez GWAS summary statistics excluded the *APOE* variants, but given the well-established large effects of *APOE* ε4 on AD risk, we obtained effect estimates for this variant from the Kunkle et al. (2019) GWAS (28). The VaD PRS utilised the recent GWAS meta-analysis by Taylor-Bateman et al. (2025), which represents the largest genetic study of VaD to date, identifying four genome-wide significant (p<5×10^-08^) and three suggestively associated (P<1×10^-06^) loci (29). The strongest association was observed for *APOE* (30), followed by genome-wide significant loci at *NECTIN2*, *APOC4*, and *APOC2*. Further details of the VaD PRS have been published previously (29). For WMH volume, we incorporated 27 independent genome-wide significant loci from Sargurupremraj et al. (2020) (31). SBP and DBP PRSs were constructed using summary statistics from Evangelou et al. (2018), encompassing 362 SBP and 405 DBP variants identified in over 750,000 individuals (32). See Supplementary tables 25-28 for variant information used in all PRSs.

### Dementia outcomes

All dementia diagnosis outcomes (AD, VaD and all-cause dementia) were obtained through algorithmic combination of self-reported data at baseline assessment, hospital admission diagnoses, and ICD-10 codes in death register records. Cases were defined using UK Biobank’s algorithmically defined diagnostic variables (UK Biobank category 47), with binary coding for presence or absence of each outcome (33).

### Covariates

We included covariates on sex, age and the first 10 genetic principal components in our analysis. Genetic principal components were included to control for genetic confounding due to ancestral differences among participants. SBP and DBP measures at baseline were also used to validate that the SBP and DBP PRSs were robustly associated with measured SBP and DBP.

### Statistical analysis

All analyses were conducted using R version 4.3.2. PRS were standardised to enable interpretation of results per standard deviation increase.

### Generation and validation of polygenic risk scores

Weighted PRSs were calculated by multiplying the dosage of risk-increasing alleles for each independent genetic variant by their respective GWAS-derived effect estimates, then summing across all variants. Included variants were independent and achieved genome-wide significance (P< 5×10^-08,^ R^2^<0.001, distance >10,000kb), except for VaD, where we included five suggestively associated variants to enhance statistical power, given the limited number of genome-wide significant loci. Given the Evangelou et al. SBP and DBP GWAS adjusts for BMI, potentially introducing collider bias given BMI’s position on the causal pathway between genetic variants and BP, we adjusted for a BMI PRS in all our models, using independent genome-wide significant loci (P< 5×10^-08,^ R^2^<0.001, distance >10,000kb) from the summary statistics by Yengo et al. (2018) (34) (supplementary Table 28).

To examine whether the PRSs used in our analysis were robustly associated with their respective trait, we used logistic regression to examine the effects of the AD, VaD and WMH PRSs on all dementia outcomes (AD, VaD and all-cause dementia diagnoses). We also used linear regression to examine whether the SBP and DBP PRSs were robustly associated with SBP and DBP measured at baseline. Models were adjusted for age, sex, the first ten genetic principal components and a BMI PRS to minimise risk of collider bias.

### Main effects of SBP and DBP on dementia risk

First, we examined main effects of the SBP and DBP PRSs on each dementia outcome (AD, VaD, and all-cause dementia) using logistic regression models, adjusted for age, sex, the first ten genetic principal components and the BMI PRS.

### Interaction analyses

To facilitate clinical interpretation, we binarised the dementia-specific PRS (AD, VaD and WMH PRSs) at the 75^th^ percentile, creating higher-risk (top quartile) versus lower-risk (bottom three quartiles) groups. Thus, we examined interactions between the SBP or DBP PRS and: (i) the binarised AD PRS for the Alzheimer’s disease diagnosis outcome, (ii) the binarised WMH PRS for the vascular dementia diagnosis outcome, (iii) the binarised VaD PRS for the vascular dementia diagnosis outcome, and (iv) both the binarised AD PRS and the binarised VaD PRS separately for the all-cause dementia diagnosis outcome. Both BP PRS were kept as continuous variables. The rationale for testing both AD and VaD PRS interactions with all-cause dementia reflects the clinical reality that mixed neurodegenerative and vascular pathology commonly contributes to dementia in older adults. Again, all models were adjusted for age, sex, the first ten genetic principal components and the BMI PRS.

### Sensitivity analyses

We conducted several sensitivity analyses to examine the robustness of interaction effects. First, we repeated analyses treating dementia PRSs as continuous variables (rather than binary strata) to test for multiplicative interactions with BP PRSs. Second, we assessed departures from additivity using the relative excess risk due to interaction (RERI), calculated from linear models with interaction terms and standard error propagation for 95% CIs. We defined RERI as *OR_11_ – OR_10_ – OR_01_ + 1*, where *OR_11_* represents combined high genetic risks, *OR_10_* represents high dementia PRS only, and OR_01_ represents high BP PRS only (35–38). For this analysis, both BP and dementia PRSs were binarised at the 75th percentile, with RERIL>L0 indicating positive additive interaction (combined effects exceeding the sum of individual contributions). Third, recognising that dementia often manifests in late life and that mixed pathology increases with age, we performed age-stratified analyses, dividing participants into <65 and ≥65 years at baseline. Models were repeated within each age stratum to assess potential age-dependent differences in main effects and interactions. Fourth, given established differences in BP trajectories and dementia risk between males and females, we repeated interaction analyses stratified by sex to investigate sex-specific effects. Finally, we examined potential confounding by hormone replacement therapy (HRT) in females. Although our use of BP PRSs (reflecting genetic predisposition) reduces the likelihood of confounding by HRT (since HRT use cannot influence inherited genetic variants), we conducted a female-only analysis adjusting for HRT status (ever vs. never used) for completeness.

## Results

### Participant Characteristics

The study comprised of 334,759 UK Biobank participants (Figure 1). We identified 3,001 cases of Alzheimer’s disease (AD), 1,442 cases of vascular dementia (VaD), and 6,717 cases of all-cause dementia. Participants with dementia were, on average, substantially older at recruitment than those without (mean age 64.7 years vs 56.8 years, difference = 7.9 years) and had a slightly higher proportion of females (53.8% vs 51.7%). Among participants with available BP measurements (n = 312,741, 93.4% of the total sample), 160,214 (47.9%) met criteria for hypertension (SBP ≥140 mmHg or DBP ≥90 mmHg).

### Validation of the polygenic risk scores

PRSs for AD, VaD, and WMH were each associated with dementia risk, with the strongest effects observed for AD PRS (Table 1). A 1 SD increase in the AD PRS was associated with a two-fold increase in AD risk (OR = 2.16, 95%CI: 2.09–2.22), and an increase in risk for all-cause dementia (OR = 1.82, 95%CI, 1.78–1.86). The VaD PRS was most strongly associated with VaD (OR = 1.54, 95%CI: 1.47–1.60), though effects extended to other dementia subtypes. WMH PRS demonstrated modest, directionally consistent associations, with the largest effect for VaD (OR = 1.10, 95%CI, 1.05–1.16). The SBP PRS was significantly associated with higher SBP (β = 2.19, 95%CI: 2.13–2.25, P<0.001), similarly the DBP PRS was strongly associated with higher DBP (β =1.32, 95%CI: 1.28–1.35, P<0.001).

**Table 1:** Associations between polygenic risk scores and demographic variables, with dementia subtypes.

| Risk Factor | AD OR (95%CI) | VaD OR (95%CI) | All-cause dementia OR (95%CI) |
| --- | --- | --- | --- |
| AD PRS (per SD) | 2.16 (2.09–2.22) *** | 1.66 (1.59–1.73) *** | 1.82 (1.78-1.86) *** |
| VaD PRS (per SD) | 1.90 (1.84-1.96) *** | 1.54 (1.47-1.60) *** | 1.67 (1.63-1.70) *** |
| WMH PRS (per SD) | 1.04 (1.00-1.08) * | 1.10 (1.05-1.16) *** | 1.05 (1.02-1.07) *** |
| SBP PRS (per SD) | 1.04 (1.01-1.08) * | 1.06 (1.00-1.11) * | 1.05 (1.03-1.08) *** |
| DBP PRS (per SD) | 0.99 (0.95-1.03) | 1.05 (0.99-1.10) | 1.00 (0.98-1.03) |
| Age (per year) | 1.24 (1.23-1.25) *** | 1.24 (1.22-1.25) *** | 1.20 (1.20-1.21) *** |
| Male (vs. female) | 0.97 (0.90-1.05) | 1.51 (1.36-1.68) *** | 1.21 (1.15-1.27) *** |

Each model includes the specified risk factor (e.g., PRS), age at recruitment, sex, and 10 genetic principal components as covariates. PRSs were analysed separately in individual models. Models examining SBP or DBP were additionally adjusted for the BMI PRS. Odds ratios (ORs) represent the effect per 1 standard deviation increase in PRS. \*p < 0.05, \*\*p < 0.01, \*\*\*p < 0.001.

### Main effects of the SBP and DBP PRSs on dementia risk

Figure 2 shows the main effects of the SBP and DBP PRSs on dementia diagnosis outcomes. The SBP PRS was consistently associated with increased dementia risk (Table 1). Each SD increase in SBP PRS resulted in 4% higher odds of AD (95%CI: 1.01–1.08), 6% for VaD (95%CI: 1.00–1.11), and 5% for all-cause dementia (95%CI: 1.03–1.08). In contrast, the DBP PRS showed little evidence of associations with AD or all-cause dementia, and only weak evidence for an association with VaD

**Figure 2:**
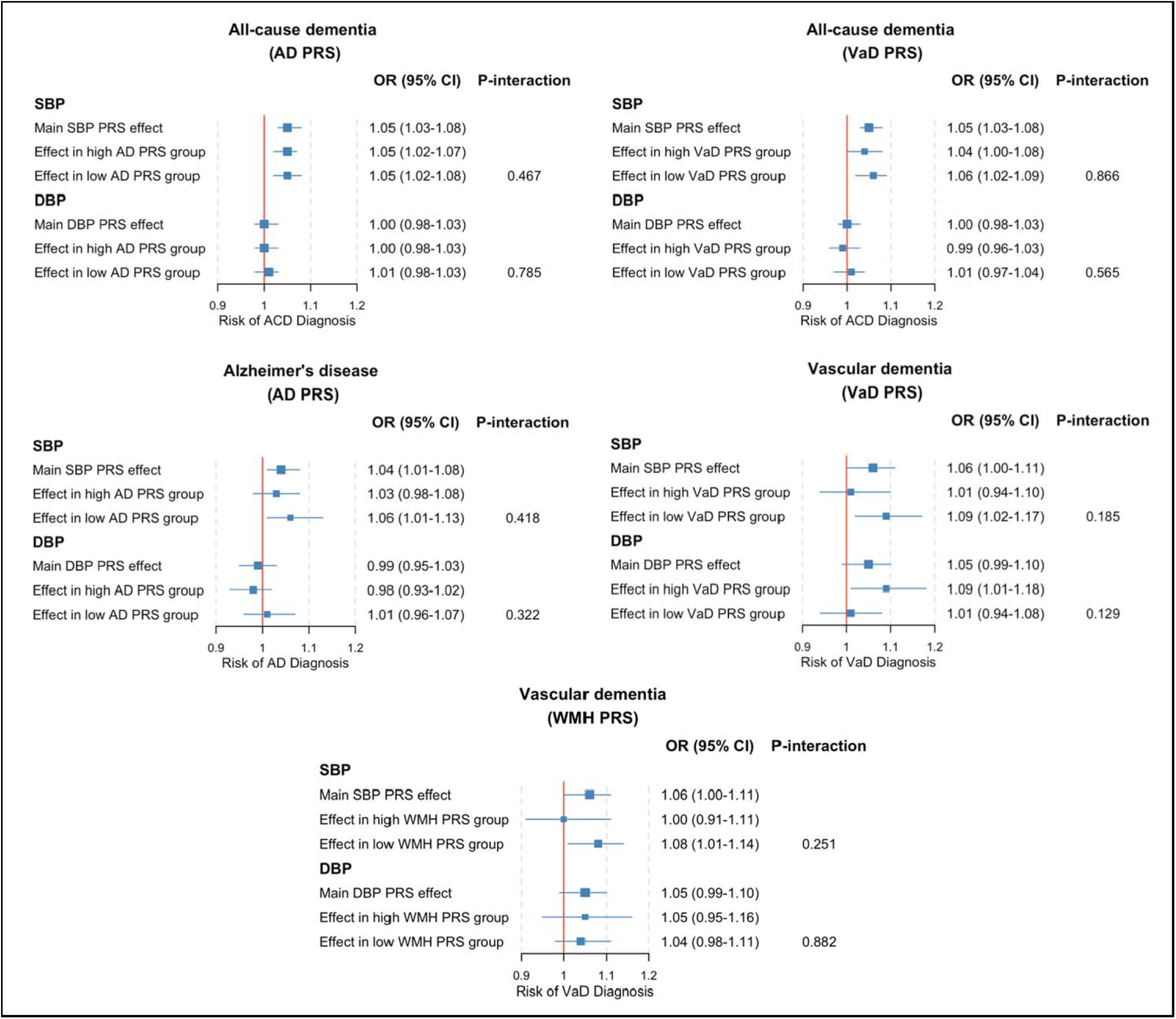
Main effects SBP/DBP (continuous) on each dementia diagnosis subtype, and interaction effects stratified by high vs low dementia PRS.

### Interaction Analyses

#### Systolic blood pressure x genetic liability to dementia

Interaction analyses with SBP are shown in Figure 2. Primary analyses examined multiplicative interactions between the continuous SBP and DBP PRS and the binary dementia-specific PRS on dementia diagnosis outcomes. Overall, these analyses did not provide strong evidence for interaction effects (Supplementary Tables 1–5).

For Alzheimer’s disease, the SBP PRS was positively associated with risk of AD diagnosis among individuals with lower AD genetic risk (OR = 1.06, 95% CI: 1.01–1.13), whereas little evidence for association was observed in those with higher AD genetic risk (OR = 1.03, 95% CI: 0.98–1.08), though the difference in effect estimates across genetic risk groups was small (P*_interaction_*= 0.418). A similar pattern was observed for VaD: the SBP PRS was positively associated with risk of VaD diagnosis among individuals with lower VaD genetic risk (OR = 1.09, 95% CI: 1.02–1.17), but showed limited evidence of association in those with higher VaD genetic risk (OR = 1.01, 95% CI: 0.94–1.10; P*_interaction_* = 0.185). Likewise, for WMH genetic risk, the SBP PRS was associated with risk of VaD diagnosis in individuals with lower WMH genetic risk (OR = 1.08, 95% CI: 1.01–1.14), with little evidence of association in those with higher risk (OR = 1.00, 95% CI: 0.91–1.11; P*_interaction_* = 0.251). For all-cause dementia, the SBP PRS showed similar positive associations across both high and low AD genetic risk groups (high AD PRS: OR = 1.05, 95% CI: 1.02–1.07; low AD PRS: OR = 1.05, 95% CI: 1.02–1.08; P*_interaction_* = 0.467). Results were also consistent across high and low VaD genetic risk groups (high VaD PRS: OR = 1.04, 95% CI: 1.00–1.08; low VaD PRS: OR = 1.06, 95% CI: 1.02–1.09; P*_interaction_*= 0.866).

#### Diastolic blood pressure x genetic liability to dementia

Interaction analyses with DBP are shown in Figure 2. There was little evidence for multiplicative interactions between the continuous DBP PRS and the binary dementia-specific PRS on dementia diagnosis outcomes (Supplementary Tables 1–5). For Alzheimer’s disease, the DBP PRS showed little evidence of association across both high and low AD genetic risk groups (high AD PRS: OR = 0.98, 95% CI: 0.93–1.02; low AD PRS: OR = 1.01, 95% CI: 0.96–1.07), with little evidence for a difference between effect estimates (P*_interaction_* = 0.322). For VaD, a different pattern was observed: the DBP PRS showed positive associations with risk of VaD diagnosis in individuals with high VaD genetic risk (OR = 1.09, 95% CI: 1.01–1.18), but not in those with low VaD genetic risk (OR = 1.01, 95% CI: 0.94– 1.08), although evidence for a difference in effect estimates across genetic risk groups was modest (P*_interaction_* = 0.129). In contrast, the DBP PRS showed no clear associations with risk of VaD diagnosis in either high or low WMH genetic risk groups (high WMH PRS: OR = 1.05, 95% CI: 0.95–1.16; low WMH PRS: OR = 1.04, 95% CI: 0.98–1.11; P*_interaction_* = 0.882). For all-cause dementia, the DBP PRS showed no clear associations across both high and low AD genetic risk groups (high AD PRS: OR = 0.99, 95% CI: 0.96–1.03; low AD PRS: OR = 1.01, 95% CI: 0.98–1.05; P*_interaction_* = 0.398), nor across high and low VaD genetic risk groups (high VaD PRS: OR = 0.99, 95% CI: 0.96–1.03; low VaD PRS: OR = 1.01, 95% CI: 0.97–1.04; P*_interaction_* = 0.565).

### Sensitivity analyses

#### Continuous dementia PRSs

Analyses treating PRSs as continuous predictors showed no strong evidence for multiplicative interactions across dementia outcomes (Supplementary Table 6). For AD, interaction terms were close to the null (P_interaction_>L0.670). For VaD, there was weak evidence for interactions with the SBP PRS (P_interaction_=L0.105) and DBP PRS (P_interaction_=L0.092). For all-cause dementia, interaction P-values ranged from 0.467 to 0.891.

#### Additive interaction models (RERI)

On the additive scale, most interactions were negligible (Supplementary Tables 7–9). However, AD PRS × SBP PRS showed evidence of supra-additive effects on all-cause dementia (RERI = 0.31, 95%LCI: 0.02–0.60; P_interaction_=L0.036). Other additive interactions (e.g. VaD × DBP PRS) showed little evidence of deviation from additivity.

#### Age-stratified analyses

The SBP PRS showed age-dependent main effects. It was associated with AD only in younger participants (OR = 1.09, 95%LCI: 1.03–1.15), not older (OR = 1.02, 95%LCI: 0.97–1.07; Supplementary Table 10). For VaD, the pattern reversed: associated only in older participants (OR = 1.08, 95%LCI: 1.01–1.15; P = 0.018), not younger (OR = 1.02, 95%LCI: 0.93–1.11; Supplementary Table 11). For all-cause dementia, the SBP PRS showed consistent associations in both age groups (OR = 1.05, 95%LCI: ∼1.01–1.09; P ≤ 0.007). The DBP PRS showed very little evidence of main effects on AD, VaD, or all-cause dementia in either age group. Interaction analyses within age strata showed little evidence overall, except for the DBP PRS in younger participants, where it was associated with VaD only in those with high VaD genetic risk (P_interaction_=L0.008); this finding should be interpreted cautiously given multiple testing.

#### Sex-stratified analyses

The SBP PRS was associated with AD in females (OR = 1.06, 95%LCI: 1.00–1.11; P = 0.035) but not males (OR = 1.03, 95%LCI: 0.98–1.09; P = 0.216; Supplementary Table 15). For VaD, it was associated in males (OR = 1.07, 95%LCI: 1.00–1.15; P = 0.047) but not females (OR = 1.04, 95%LCI: 0.96–1.13; P = 0.361; Supplementary Table 16). For all-cause dementia, the SBP PRS showed consistent associations in both sexes (OR = 1.05, 95%LCI: ∼1.01–1.09; P ≤ 0.008). The DBP PRS showed little evidence of main effects in either sex (Supplementary Tables 15–19). Overall, there was very little evidence of interactions by sex for both SBP and DBP.

#### HRT adjustment in females

Adjustment for HRT made little difference to our results. The only changes were that, after HRT adjustment, the SBP PRS was associated with all-cause dementia only in low AD genetic risk (OR = 1.07, 95%LCI: 1.02–1.13; P_interaction_=L0.008; Supplementary Table 23), and the DBP PRS was associated with VaD only in high VaD genetic risk (OR = 1.15, 95%LCI: 1.02–1.30; P_interaction_=L0.023; Supplementary Table 21).

## Discussion

Our findings provide strong evidence to support an overall causal effect of SBP on all dementia subtypes, resulting in 4-6% increased odds of diagnosis, on average, across dementia outcomes per SD increase in the SBP PRS. There was very little evidence to support an overall causal effect of DBP on risk of any dementia diagnosis.

Given the conflicting findings from existing observational causal inference studies (e.g. MR and RCT evidence) on the effects of BP on dementia risk, we hypothesised that the effect of BP on dementia risk may depend on the underlying neuropathological context, and that BP may interact with pre-existing or underlying neurodegenerative or cerebrovascular vulnerability to accelerate pathological processes and ultimately lead to greater cognitive decline and a higher risk of dementia diagnosis. However, findings from our interaction analyses do not support this hypothesis; overall, we found very little evidence to suggest that BP interacts with underlying genetic predisposition to either AD or VaD to influence dementia risk. Instead, our results suggest that elevated SBP increases dementia risk in a relatively uniform way, without strong modification by underlying genetic risk for AD or VaD.

Our use of PRSs, which capture lifelong genetic predisposition to higher BP, avoids confounding from age-dependent changes and measurement timing that complicate observational studies. While midlife hypertension is consistently associated with increased dementia risk, BP measured in late life often shows inconsistent or even paradoxical patterns due to reverse causation and frailty (39). By leveraging genetic instruments, our approach provides a more stable estimate of the long-term impact of elevated BP on dementia risk across subtypes.

Our findings align with emerging evidence suggesting that cerebrovascular dysregulation may represent one of the earliest pathological events in late-onset AD development. Using multifactorial analysis of over 7,700 brain images, Iturria-Medina et al. (2016) demonstrated that cerebral blood flow abnormalities emerge before amyloid deposition, metabolic changes, or structural atrophy (40). Their temporal ordering model places vascular changes as the primary event, with amyloid pathology developing secondarily within an already compromised vascular environment. This sequence could explain why BP operates independently of dementia genetic risk in our analyses: if vascular dysfunction establishes the foundational substrate for disease, subsequent neurodegeneration influenced by AD-specific genetic factors occurs against this pre-existing vascular backdrop. This reinforces the case for population-wide interventions to control SBP to reduce dementia burden, targeting the earliest detectable disease stage, rather than intervening only in high-risk groups.

Very few studies have previously examined an interaction between genetic risk for dementia and BP. One study by Littlejohns et al. (2022) examined whether a PRS for dementia containing 39 dementia associated genetic variants from the Ebenau et al. (2021) GWAS (41), plus *APOE* ε4 status, modified the association between hypertension and all-cause dementia risk in 198,000 UK Biobank participants. Like our study, they reported very little evidence of an interaction (P*_interaction_* = 0.2). Our analysis builds on this work by incorporating 75 AD-associated variants from the most recent large-scale GWAS by Bellenguez et al. (2022) (27), and also considered dementia subtypes. Beyond the methodological enhancement, our study differs in several important aspects. We examined both SBP and DBP across the whole spectrum rather than a diagnosis of hypertension; examined genetic risk of both AD and cerebrovascular disease separately (using three PRSs for AD, VaD and WMH); and examined dementia subtypes rather than just all-cause dementia. Littlejohns et al., also examined associations using observed hypertension (as opposed to a genetic proxy for SBP/DBP as in our study), which could potentially result in residual confounding and reverse causation bias. In contrast, a second study in participants from the University of Pittsburgh Adult Health and Behavior project (AHAB) examined whether the combined presence of the *APOE* ε4 allele and elevated BP was associated with lower cognitive performance in cognitively healthy middle-aged adults (aged 30 to 54). That study concluded that the joint presence of *APOE* ε4 and elevated SBP, even at prehypertensive levels, was associated with lower cognitive performance, suggesting they may synergistically compromise memory function long before the appearance of clinically significant impairments. However, it is worth noting that these participants were, on average, relatively young and were not followed-up long enough to establish whether the observed difference in cognitive function in mid-life ultimately translated into a difference in dementia diagnosis rates across genetic risk groups in late life. The AHAB study also only used *APOE* ε4 as a marker of genetic risk for dementia, and did not consider cerebrovascular pathology.

Our sex-stratified analyses provided tentative evidence that SBP is only associated with AD in females, with VaD only in males, with all-cause dementia in both sexes. Importantly, adjustment for hormone replacement therapy in females did not substantially alter these relationships, suggesting that observed sex differences reflect deeper biological mechanisms rather than simply hormonal confounding. These findings align with established sex differences in cardiovascular aging, where females experience distinct BP trajectories following menopause that may influence dementia pathogenesis differently than the more linear patterns observed in males (42,43). Importantly, our HRT adjusted analysis addresses a gap in the current literature, as previous studies examining sex differences in cardiovascular-cognitive relationships have typically not controlled for hormone therapy use, despite it being an important confounder, given that HRT affects both vascular function and cognitive outcomes in postmenopausal women (40,41). Overall, findings to-date suggest that an interaction between genetic risk for dementia and BP is unlikely to fully explain the heterogeneity we observe in the literature on this risk factor-disease association. Instead, these relationships appear to involve complex, potentially sex-dependent mechanisms, that operate largely independently across strata for genetic liability to dementia.

### Strengths and Limitations

Our study has several strengths. Subgroup analyses can typically be underpowered, however, we utilised a large population-based cohort and confidence intervals within subgroups were relatively precise, suggesting we were well powered to detect any interaction effects. We also used a well-validated genetic proxy for both SBP and DBP, avoiding the potential for reverse causation and confounding when using measured SBP and DBP. We also examined genetic risk using the most comprehensive dementia genetic architecture available, incorporating 75 AD-associated variants from the latest large-scale GWAS; nearly double the genetic coverage of previous interaction studies. Our study encompassed both Alzheimer’s and cerebrovascular pathology, and examined diagnoses by dementia subtype. Lastly, we examined interactions on both the multiplicative and additive scale, which most previous studies have not done and is important for providing a complete understanding of how two risk factors jointly influence disease risk. Additionally, our sex-stratified analyses addresses important biological and methodological considerations, aligning with current best practices for inclusive research design and, if replicated in independent representative studies, potentially shedding light on some sex-specific patterns.

There are also some limitations to our study. Firstly, restricting our analysis to individuals of white British ancestry was necessary to control for potential genetic confounding due to population structure, but it also limits generalizability of the findings to other ethnic groups. Genetic risk effects for dementia may differ across populations, and thus our results may not be applicable to non-European populations. Secondly, the GWASs used to create our SBP and DBP PRSs adjusted for BMI. Adjusting for covariates downstream of genes in a GWAS can potentially introduce collider bias and distort SNP-trait estimates, particularly in the case of having weak instruments (which we do not have for SBP and DBP in our study). To mitigate this risk, we adjusted for a BMI PRS in all analyses, and results were very similar with and without this adjustment. Thirdly, UK Biobank dementia diagnoses are algorithmically defined from both administrative records and self-report, and accurately diagnosing dementia subtype pre-mortem is notoriously challenging, so there is likely to be some misclassification. Lastly, the UK Biobank cohort is middle-aged at baseline and healthier than the general population, and the prevalence of dementia cases in this cohort is relatively low given the follow-up period, which may limit generalisability to older or more diverse populations.

## Conclusion

In conclusion, our study supports elevated SBP as an independent risk factor for dementia across subtypes, with little evidence that this effect is modified by genetic predisposition to Alzheimer’s disease or vascular dementia. These findings reinforce the importance of population-wide strategies to lower SBP as a means of reducing dementia risk at the population level.

## Data Availability

The genetic and phenotypic data underlying this research were accessed from the UK Biobank under application number 123335. UK Biobank data are available to researchers through application to the UK Biobank Access Management System (https://www.ukbiobank.ac.uk/enable-your-research/apply-for-access). Information on summary statistics from the GWAS used to construct the polygenic risk score can be found within Methods. Detailed analysis code and supplementary materials are available upon reasonable request to the authors.

## Author’s contributions

ELA and YBS conceived the idea for the study. PB an RM conducted all statistical analyses under the supervision of ELA. VTB conducted the GWAS of VaD for a previous study, which enabled generation of the VaD PRS for this study. PB and ELA wrote the first draft of the manuscript, with critical comments from YBS, VTB and PGK.

## Acknowledgements

This research was conducted using data from the UK Biobank. We are grateful to the UK Biobank participants and the team for their contributions and for maintaining this important resource. UK Biobank is an open access resource available to verified researchers upon application (http://www.ukbiobank.ac.uk/). We are also grateful to the MEGAVCID consortium for provided us with GWAS summary statistics for vascular dementia.

## Supplementary Tables

### Primary Interaction Analyses

**Supplementary Table 1:** Blood pressure PRS effects on Alzheimer’s disease by AD genetic risk level.

| Variable | High AD PRS | Low AD PRS | P for interaction |
| --- | --- | --- | --- |
| SBP PRS | 1.03 (0.98-1.08) | 1.06 (1.01-1.13) * | 0.418 |
| DBP PRS | 0.98 (0.93-1.02) | 1.01 (0.96-1.07) | 0.322 |

All models adjusted for age, sex, BMI PRS, and 10 genetic principal components. Values are odds ratios (OR) (95% confidence intervals), that represent the effect per 1 standard deviation increase in SBP or DBP PRS. High genetic risk defined as top quartile (>75th percentile); low genetic risk defined as bottom three quartiles (≤75th percentile). \*p < 0.05, \*\*p < 0.01, \*\*\*p < 0.001.

**Supplementary Table 2:**
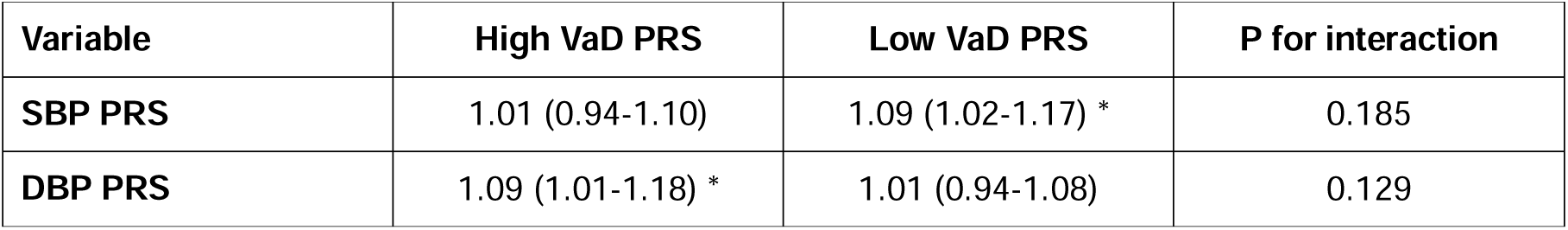
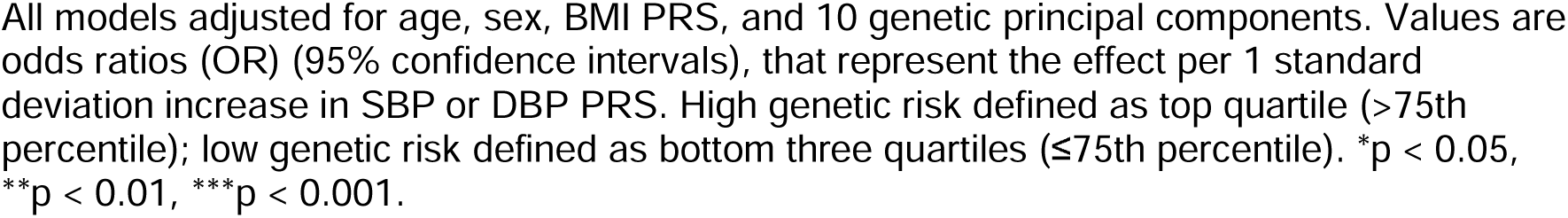
Blood pressure PRS effects on vascular dementia by VaD genetic risk level.

| Variable | High VaD PRS | Low VaD PRS | P for interaction |
| --- | --- | --- | --- |
| SBP PRS | 1.01 (0.94-1.10) | 1.09 (1.02-1.17) * | 0.185 |
| DBP PRS | 1.09 (1.01-1.18) * | 1.01 (0.94-1.08) | 0.129 |

All models adjusted for age, sex, BMI PRS, and 10 genetic principal components. Values are odds ratios (OR) (95% confidence intervals), that represent the effect per 1 standard deviation increase in SBP or DBP PRS. High genetic risk defined as top quartile (>75th percentile); low genetic risk defined as bottom three quartiles ( ≤ 75th percentile). \* $p < 0.05$ , \*\* $p < 0.01$ , \*\*\* $p < 0.001$ .

**Supplementary Table 3:** Blood pressure PRS effects on vascular dementia by WMH genetic risk level.

| Variable | High WMH PRS | Low WMH PRS | P for interaction |
| --- | --- | --- | --- |
| **SBP PRS** | 1.00 (0.91-1.11) | 1.08 (1.01-1.14) * | 0.251 |
| **DBP PRS** | 1.05 (0.95-1.16) | 1.04 (0.98-1.11) | 0.882 |

All models adjusted for age, sex, BMI PRS, and 10 genetic principal components. Values are odds ratios (OR) (95% confidence intervals), that represent the effect per 1 standard deviation increase in SBP or DBP PRS. High genetic risk defined as top quartile (>75th percentile); low genetic risk defined as bottom three quartiles ( ≤ 75th percentile). \* $p < 0.05$ , \*\* $p < 0.01$ , \*\*\* $p < 0.001$ .

**Supplementary Table 4:** Blood pressure PRS effects on all-cause dementia by AD genetic risk level.

| Variable | High AD PRS | Low AD PRS | P for interaction |
| --- | --- | --- | --- |
| **SBP PRS** | 1.05 (1.02-1.07) ** | 1.05 (1.02-1.08) ** | 0.467 |
| **DBP PRS** | 1.00 (0.98-1.03) | 1.01 (0.98-1.03) | 0.785 |

All models adjusted for age, sex, BMI PRS, and 10 genetic principal components. Values are odds ratios (OR) (95% confidence intervals), that represent the effect per 1 standard deviation increase in SBP or DBP PRS. High genetic risk defined as top quartile (>75th percentile); low genetic risk defined as bottom three quartiles ( ≤ 75th percentile). \* $p < 0.05$ , \*\* $p < 0.01$ , \*\*\* $p < 0.001$ .

**Supplementary Table 5:**
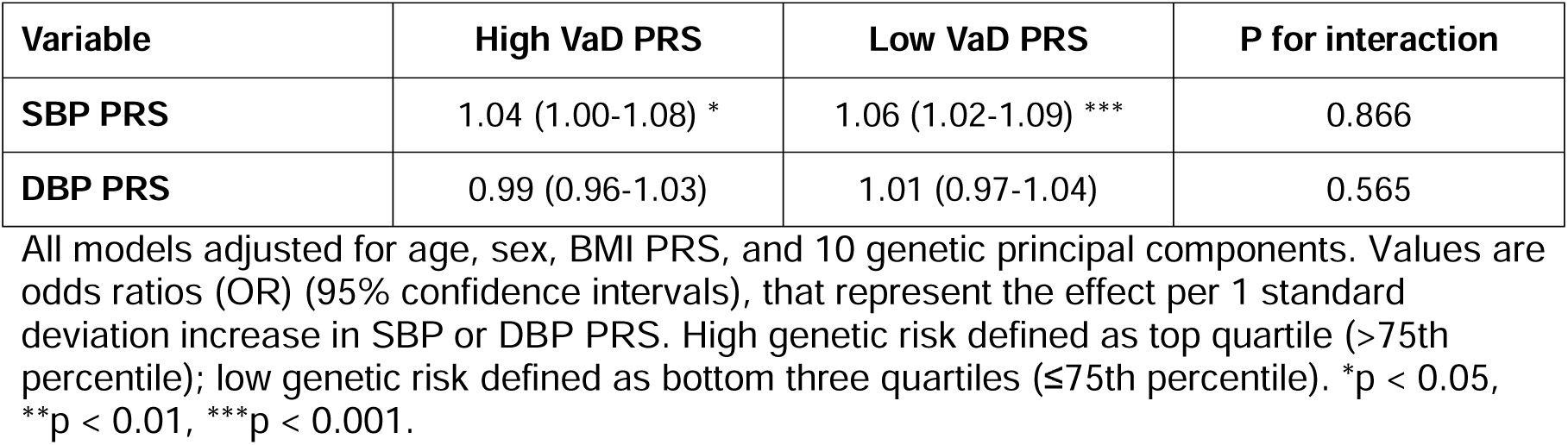
Blood pressure PRS effects on all-cause dementia by VaD genetic risk level.

| Variable | High VaD PRS | Low VaD PRS | P for interaction |
| --- | --- | --- | --- |
| **SBP PRS** | 1.04 (1.00-1.08) * | 1.06 (1.02-1.09) *** | 0.866 |
| **DBP PRS** | 0.99 (0.96-1.03) | 1.01 (0.97-1.04) | 0.565 |

All models adjusted for age, sex, BMI PRS, and 10 genetic principal components. Values are odds ratios (OR) (95% confidence intervals), that represent the effect per 1 standard deviation increase in SBP or DBP PRS. High genetic risk defined as top quartile (>75th percentile); low genetic risk defined as bottom three quartiles ( ≤ 75th percentile). \* $p < 0.05$ , \*\* $p < 0.01$ , \*\*\* $p < 0.001$ .

### Sensitivity analyses

#### Continuous Dementia PRS Interaction Analysis

**Supplementary Table 6.** Multiplicative interactions between continuous dementia and blood pressure polygenic risk scores across dementia outcomes.

| Variable | Alzheimer's disease<br>OR (95%CI) | Vascular dementia<br>OR (95%CI) | All-cause dementia<br>OR (95%CI) |
| --- | --- | --- | --- |
| AD PRS x SBP PRS | 0.99 (0.97–1.01),<br>P = 0.256 | - | 0.99 (0.97–1.01),<br>P = 0.467 |
| AD PRS x DBP PRS | 1.00 (0.98–1.02),<br>P = 0.678 | - | 1.00 (0.98–1.02),<br>P = 0.891 |
| VaD PRS x SBP PRS | - | 0.96 (0.92–1.01),<br>P = 0.105 | 1.00 (0.98–1.02),<br>P = 0.866 |
| VaD PRS x DBP PRS | - | 1.04 (0.99–1.08),<br>P = 0.092 | 0.99 (0.97–1.01),<br>P = 0.565 |
| WMH PRS x SBP PRS | - | 0.96 (0.91–1.01),<br>P = 0.089 | - |
| WMH PRS v DBP PRS | - | 1.00 (0.94–1.05),<br>P = 0.840 | - |

All models adjusted for age, sex, BMI PRS, and 10 genetic principal components. Values are odds ratios (OR) (95% confidence intervals), that represent the effect per 1 standard deviation increase in PRS. Dementia-specific PRS and blood pressure PRS are both continuous.

#### Additive Interaction Analyses on Risk-Stratified PRS

**Supplementary Table 7:** Risk-stratified additive interactions for Alzheimer’s disease.

| Variable | OR (95%CI) | P value | RERI (95%CI) | P value |
| --- | --- | --- | --- | --- |
| AD PRS | 5.18 (4.78–5.62) | <0.001 | - | - |
| SBP PRS | 1.06 (0.92–1.21) | 0.283 | - | - |
| DBP PRS | 0.98 (0.82–1.17) | 0.787 | - | - |
| AD PRS x SBP PRS | 1.09 (0.88–1.34) | 0.761 | 0.40 (-0.16, 0.96) | 0.157 |
| AD PRS x DBP PRS | 0.95 (0.85–1.13) | 0.557 | -0.18 (-0.71, 0.36) | 0.522 |

All models adjusted for age, sex, BMI PRS, and 10 genetic principal components. Values are odds ratios (OR) (95% confidence intervals), that represent the effect per 1 standard deviation increase in PRS. Dementia-specific PRS and blood pressure PRS are both binarised in high (top 25%) vs low risk. RERI = relative excess risk due to interactions.

**Supplementary Table 8:**
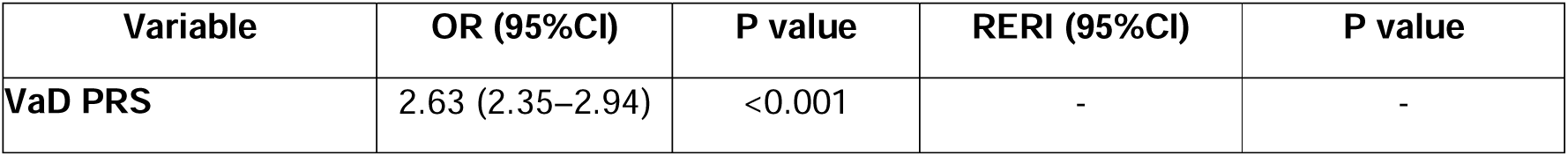
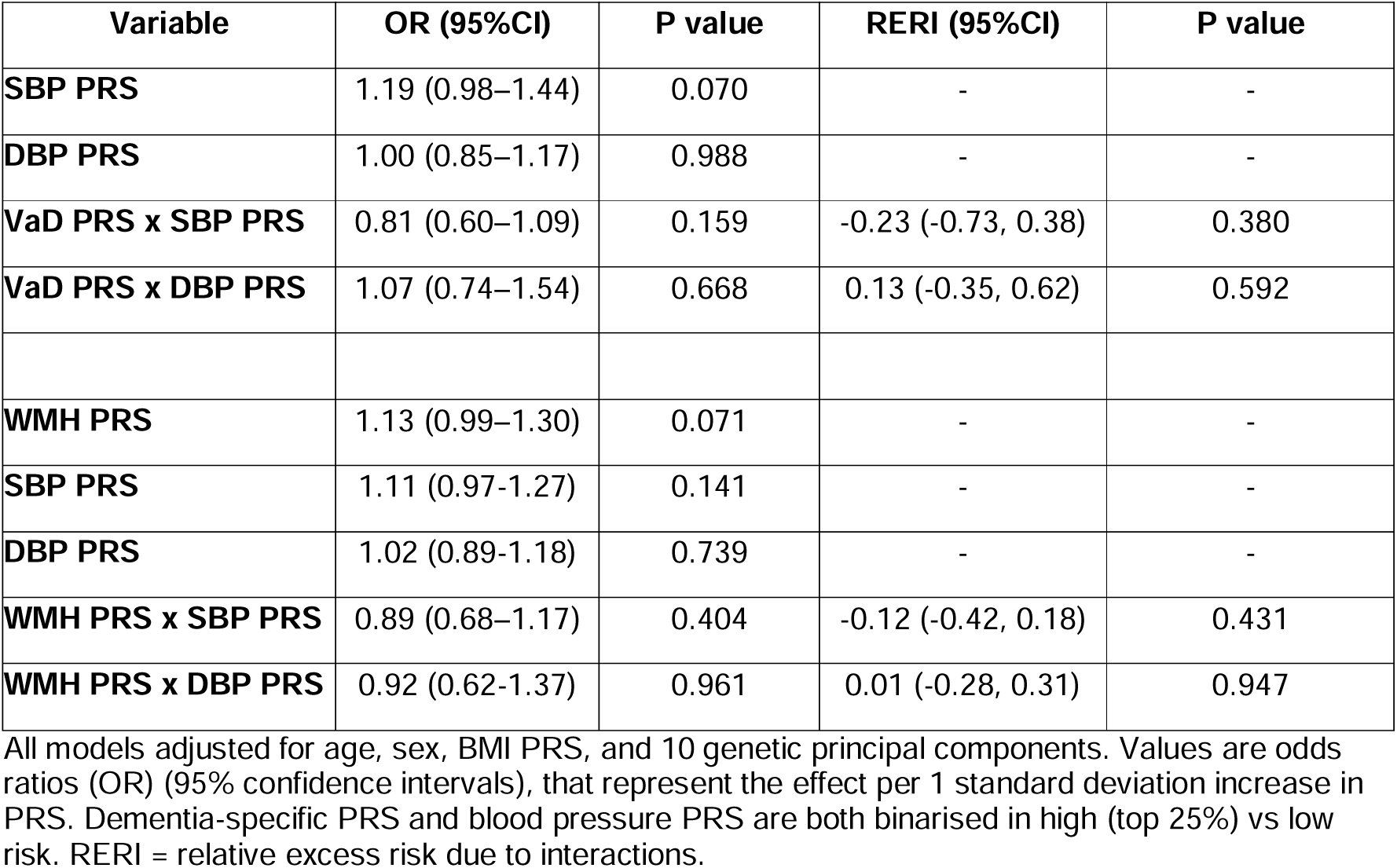
Risk-stratified additive interactions for vascular dementia.

| Variable | OR (95%CI) | P value | RERI (95%CI) | P value |
| --- | --- | --- | --- | --- |
| VaD PRS | 2.63 (2.35–2.94) | <0.001 | - | - |
| **SBP PRS** | 1.19 (0.98–1.44) | 0.070 | - | - |
| **DBP PRS** | 1.00 (0.85–1.17) | 0.988 | - | - |
| **VaD PRS x SBP PRS** | 0.81 (0.60–1.09) | 0.159 | -0.23 (-0.73, 0.38) | 0.380 |
| **VaD PRS x DBP PRS** | 1.07 (0.74–1.54) | 0.668 | 0.13 (-0.35, 0.62) | 0.592 |
| **WMH PRS** | 1.13 (0.99–1.30) | 0.071 | - | - |
| **SBP PRS** | 1.11 (0.97-1.27) | 0.141 | - | - |
| **DBP PRS** | 1.02 (0.89-1.18) | 0.739 | - | - |
| **WMH PRS x SBP PRS** | 0.89 (0.68–1.17) | 0.404 | -0.12 (-0.42, 0.18) | 0.431 |
| **WMH PRS x DBP PRS** | 0.92 (0.62-1.37) | 0.961 | 0.01 (-0.28, 0.31) | 0.947 |

All models adjusted for age, sex, BMI PRS, and 10 genetic principal components. Values are odds ratios (OR) (95% confidence intervals), that represent the effect per 1 standard deviation increase in PRS. Dementia-specific PRS and blood pressure PRS are both binarised in high (top 25%) vs low risk. RERI = relative excess risk due to interactions.

**Supplementary Table 9:** Risk-stratified additive interactions for all-cause dementia.

| Variable | OR (95%CI) | P value | RERI (95%CI) | P value |
| --- | --- | --- | --- | --- |
| **AD PRS** | 3.24 (3.06–3.43) | <0.001 | - | - |
| **SBP PRS** | 1.08 (0.99–1.18) | 0.056 | - | - |
| **DBP PRS** | 1.00 (0.89–1.11) | 0.443 | - | - |
| **AD PRS x SBP PRS** | 1.09 (0.94–1.27) | 0.496 | 0.31 (0.02-0.60) | 0.036 |
| **AD PRS x DBP PRS** | 0.98 (0.88–1.10) | 0.762 | -0.12 (-0.40, 1.15) | 0.369 |
| **VaD PRS** | 2.99 (2.83–3.15) | <0.001 | - | - |
| **SBP PRS** | 1.09 (1.01–1.18) | 0.025 | - | - |
| **DBP PRS** | 0.98 (0.86–1.11) | 0.509 | - | - |
| **VaD PRS x SBP PRS** | 1.01 (0.90–1.13) | 0.917 | 0.19 (-0.08, 0.46) | 0.158 |
| **VaD PRS x DBP PRS** | 0.96 (0.86–1.08) | 0.529 | -0.16 (-0.41, 0.10) | 0.227 |

All models adjusted for age, sex, BMI PRS, and 10 genetic principal components. Values are odds ratios (OR) (95% confidence intervals), that represent the effect per 1 standard deviation increase in PRS. Dementia-specific PRS and blood pressure PRS are both binarised in high (top 25%) vs low risk. RERI = relative excess risk due to interactions.

#### Age-Stratified Analysis

**Supplementary Table 10:**
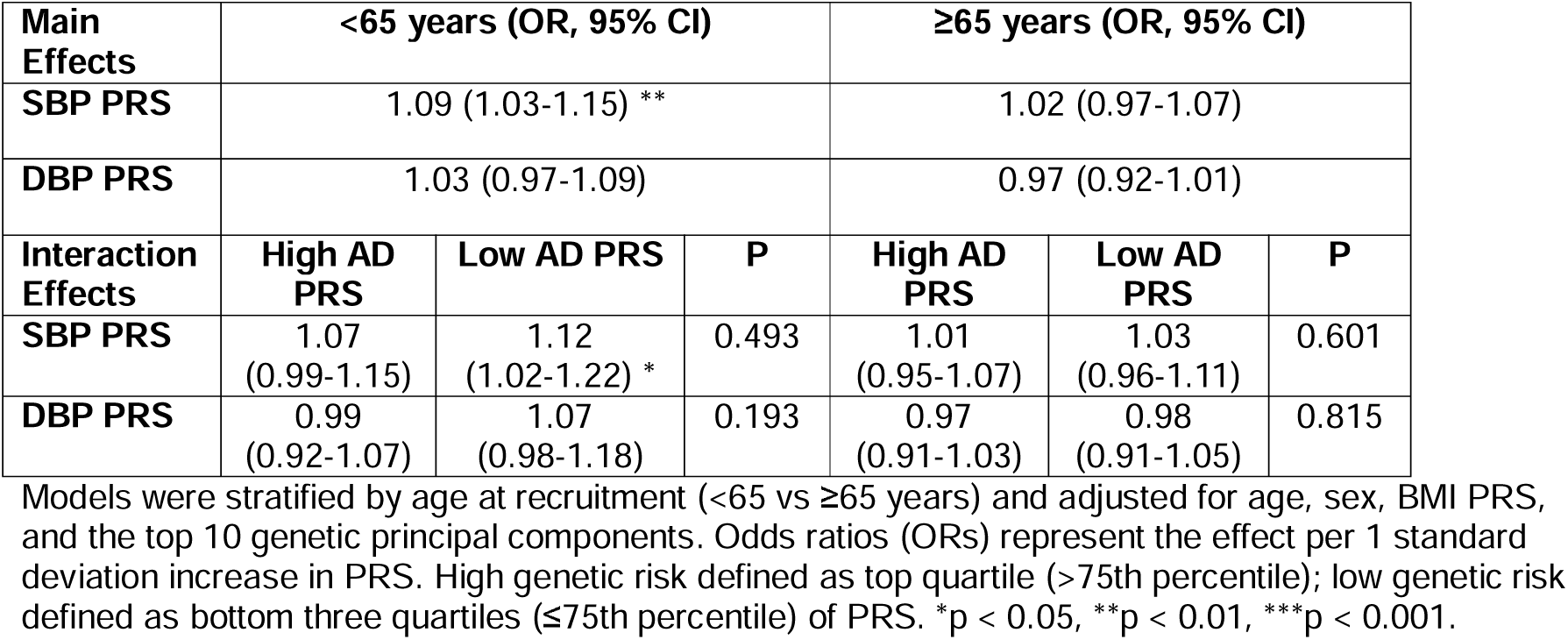
Age-stratified blood pressure PRS effects on Alzheimer’s disease by AD genetic risk level.

| Main Effects | <65 years (OR, 95% CI) |  |  | ≥65 years (OR, 95% CI) |  |  |
| --- | --- | --- | --- | --- | --- | --- |
| SBP PRS | 1.09 (1.03-1.15) ** |  |  | 1.02 (0.97-1.07) |  |  |
| DBP PRS | 1.03 (0.97-1.09) |  |  | 0.97 (0.92-1.01) |  |  |
| Interaction Effects | High AD PRS | Low AD PRS | P | High AD PRS | Low AD PRS | P |
| SBP PRS | 1.07<br>(0.99-1.15) | 1.12<br>(1.02-1.22) * | 0.493 | 1.01<br>(0.95-1.07) | 1.03<br>(0.96-1.11) | 0.601 |
| DBP PRS | 0.99<br>(0.92-1.07) | 1.07<br>(0.98-1.18) | 0.193 | 0.97<br>(0.91-1.03) | 0.98<br>(0.91-1.05) | 0.815 |

Models were stratified by age at recruitment (<65 vs ≥65 years) and adjusted for age, sex, BMI PRS, and the top 10 genetic principal components. Odds ratios (ORs) represent the effect per 1 standard deviation increase in PRS. High genetic risk defined as top quartile (>75th percentile); low genetic risk defined as bottom three quartiles (≤75th percentile) of PRS. \*p < 0.05, \*\*p < 0.01, \*\*\*p < 0.001.

**Supplementary Table 11:** Age-stratified blood pressure PRS effects on vascular dementia by VaD genetic risk level.

| Main Effects | <65 years (OR, 95% CI) |  |  | ≥65 years (OR, 95% CI) |  |  |
| --- | --- | --- | --- | --- | --- | --- |
| SBP PRS | 1.02 (0.93-1.11) |  |  | 1.08 (1.01-1.15) * |  |  |
| DBP PRS | 1.06 (0.97-1.15) |  |  | 1.04 (0.97-1.11) |  |  |
| Interaction Effects | High VaD PRS | Low VaD PRS | P | High VaD PRS | Low VaD PRS | P |
| SBP PRS | 0.96<br>(0.84-1.09) | 1.06<br>(0.94-1.18) | 0.270 | 1.05<br>(0.95-1.16) | 1.11<br>(1.01-1.21) * | 0.405 |
| DBP PRS | 1.21<br>(1.06-1.38) ** | 0.95<br>(0.85-1.07) | 0.008 | 1.04<br>(0.94-1.14) | 1.04<br>(0.95-1.14) | 0.951 |

Models were stratified by age at recruitment (<65 vs ≥65 years) and adjusted for sex, BMI PRS, and the top 10 genetic principal components. Odds ratios (ORs) represent the effect per 1 standard deviation increase in PRS. High genetic risk defined as top quartile (>75th percentile); low genetic risk defined as bottom three quartiles (≤75th percentile) of PRS. \*p < 0.05, \*\*p < 0.01, \*\*\*p < 0.001.

**Supplementary Table 12:**
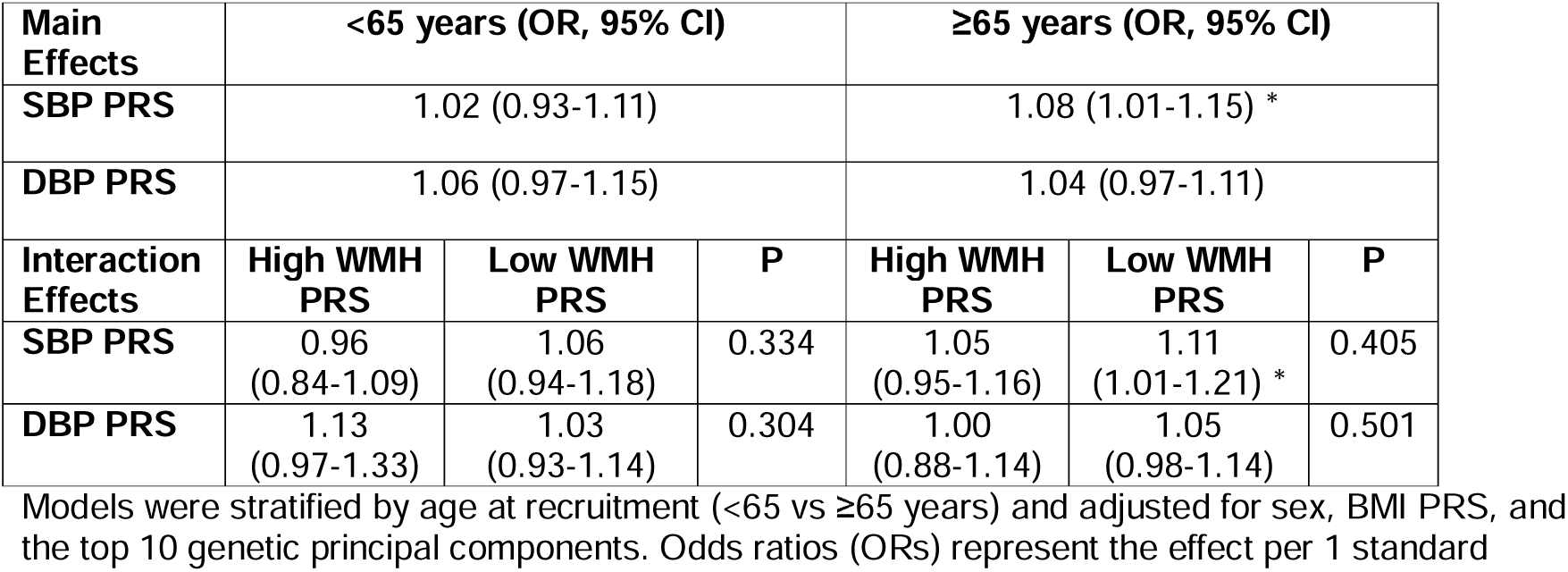
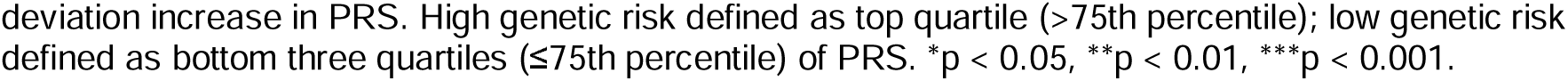
Age-stratified blood pressure PRS effects on vascular dementia by WMH genetic risk level.

| Main Effects | <65 years (OR, 95% CI) |  |  | ≥65 years (OR, 95% CI) |  |  |
| --- | --- | --- | --- | --- | --- | --- |
| SBP PRS | 1.02 (0.93-1.11) |  |  | 1.08 (1.01-1.15) * |  |  |
| DBP PRS | 1.06 (0.97-1.15) |  |  | 1.04 (0.97-1.11) |  |  |
| Interaction Effects | High WMH PRS | Low WMH PRS | P | High WMH PRS | Low WMH PRS | P |
| SBP PRS | 0.96<br>(0.84-1.09) | 1.06<br>(0.94-1.18) | 0.334 | 1.05<br>(0.95-1.16) | 1.11<br>(1.01-1.21) * | 0.405 |
| DBP PRS | 1.13<br>(0.97-1.33) | 1.03<br>(0.93-1.14) | 0.304 | 1.00<br>(0.88-1.14) | 1.05<br>(0.98-1.14) | 0.501 |

Models were stratified by age at recruitment (<65 vs ≥65 years) and adjusted for sex, BMI PRS, and the top 10 genetic principal components. Odds ratios (ORs) represent the effect per 1 standard
deviation increase in PRS. High genetic risk defined as top quartile (>75th percentile); low genetic risk defined as bottom three quartiles ( ≤ 75th percentile) of PRS. \* $p < 0.05$ , \*\* $p < 0.01$ , \*\*\* $p < 0.001$ .

**Supplementary Table 13:** Age-stratified blood pressure PRS effects on all-cause dementia by AD genetic risk level.

| Main Effects | <65 years (OR, 95% CI) | | | $\geq 65$ years (OR, 95% CI) | | |
| --- | --- | --- | --- | --- | --- | --- |
| SBP PRS | 1.05 (1.01-1.09) ** |  |  | 1.05 (1.02-1.08) ** |  |  |
| DBP PRS | 1.00 (0.96-1.04) |  |  | 1.00 (0.97-1.03) |  |  |
| Interaction Effects | High AD PRS | Low AD PRS | P | High AD PRS | Low AD PRS | P |
| SBP PRS | 1.05<br>(0.99-1.11) | 1.06<br>(1.00-1.11) * | 0.878 | 1.05<br>(1.00-1.10) | 1.05<br>(1.01-1.10) * | 0.859 |
| DBP PRS | 0.99<br>(0.94-1.05) | 1.00<br>(0.95-1.06) | 0.820 | 0.99<br>(0.94-1.04) | 1.01<br>(0.97-1.06) | 0.514 |

Models were stratified by age at recruitment (<65 vs $\geq 65$ years) and adjusted for sex, BMI PRS, and the top 10 genetic principal components. Odds ratios (ORs) represent the effect per 1 standard deviation increase in PRS. High genetic risk defined as top quartile (>75th percentile); low genetic risk defined as bottom three quartiles ( ≤ 75th percentile) of PRS. \* $p < 0.05$ , \*\* $p < 0.01$ , \*\*\* $p < 0.001$ .

**Supplementary Table 14:** Age-stratified blood pressure PRS effects on all-cause dementia by VaD genetic risk level.

| Main Effects | <65 years (OR, 95% CI) | | | $\geq 65$ years (OR, 95% CI) | | |
| --- | --- | --- | --- | --- | --- | --- |
| SBP PRS | 1.05 (1.01-1.09) ** |  |  | 1.05 (1.02-1.08) ** |  |  |
| DBP PRS | 1.00 (0.96-1.04) |  |  | 1.00 (0.97-1.03) |  |  |
| Interaction Effects | High VaD PRS | Low VaD PRS | P | High VaD PRS | Low VaD PRS | P |
| SBP PRS | 1.06<br>(1.00-1.12) | 1.05<br>(1.00-1.10) | 0.824 | 1.03<br>(0.98-1.08) | 1.07<br>(1.02-1.12) ** | 0.237 |
| DBP PRS | 1.00<br>(0.94-1.06) | 1.00<br>(0.95-1.05) | 0.990 | 1.00<br>(0.94-1.04) | 1.01<br>(0.97-1.06) | 0.514 |

Models were stratified by age at recruitment (<65 vs $\geq 65$ years) and adjusted for sex, BMI PRS, and the top 10 genetic principal components. Odds ratios (ORs) represent the effect per 1 standard deviation increase in PRS. High genetic risk defined as top quartile (>75th percentile); low genetic risk defined as bottom three quartiles ( ≤ 75th percentile) of PRS. \* $p < 0.05$ , \*\* $p < 0.01$ , \*\*\* $p < 0.001$ .

**Supplementary Table 15:** Sex-stratified blood pressure PRS effects on Alzheimer’s disease by AD genetic risk level.

| Main Effects | Female (OR, 95% CI) |  |  | Male (OR, 95% CI) |  |  |
| --- | --- | --- | --- | --- | --- | --- |
| SBP PRS | 1.06 (1.00-1.11) * |  |  | 1.03 (0.98-1.09) |  |  |
| DBP PRS | 1.01 (0.96-1.06) |  |  | 0.97 (0.92-1.03) |  |  |
| Interaction Effects | High AD PRS | Low AD PRS | P | High AD PRS | Low AD PRS | P |
| SBP PRS | 1.06<br>(0.99-1.13) | 1.06<br>(0.98-1.15) | 0.900 | 1.01<br>(0.94-1.08) | 1.07<br>(0.99-1.15) | 0.291 |
| DBP PRS | 1.02<br>(0.95-1.09) | 1.00<br>(0.92-1.08) | 0.700 | 0.93<br>(0.87-1.00) | 1.03<br>(0.95-1.11) | 0.061 |

Models were stratified by sex and adjusted for BMI PRS, and the top 10 genetic principal components. Odds ratios (ORs) represent the effect per 1 standard deviation increase in PRS. High genetic risk defined as top quartile (>75th percentile); low genetic risk defined as bottom three quartiles ( ≤ 75th percentile) of PRS. \*p < 0.05, \*\*p < 0.01, \*\*\*p < 0.001.

#### Sex-Stratified Analysis

**Supplementary Table 16:**
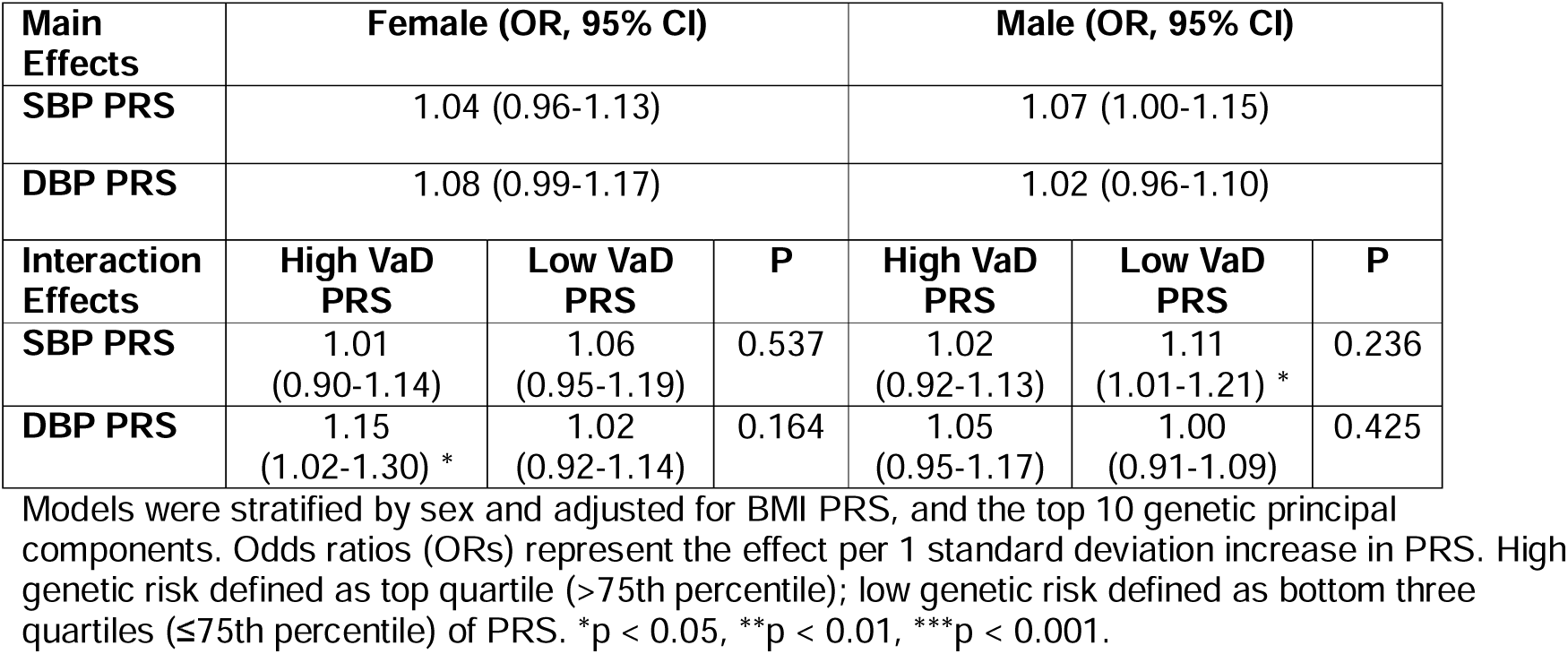
Sex-stratified blood pressure PRS effects on vascular dementia by VaD genetic risk level.

| Main Effects | Female (OR, 95% CI) |  |  | Male (OR, 95% CI) |  |  |
| --- | --- | --- | --- | --- | --- | --- |
| SBP PRS | 1.04 (0.96-1.13) |  |  | 1.07 (1.00-1.15) |  |  |
| DBP PRS | 1.08 (0.99-1.17) |  |  | 1.02 (0.96-1.10) |  |  |
| Interaction Effects | High VaD PRS | Low VaD PRS | P | High VaD PRS | Low VaD PRS | P |
| SBP PRS | 1.01<br>(0.90-1.14) | 1.06<br>(0.95-1.19) | 0.537 | 1.02<br>(0.92-1.13) | 1.11<br>(1.01-1.21) * | 0.236 |
| DBP PRS | 1.15<br>(1.02-1.30) * | 1.02<br>(0.92-1.14) | 0.164 | 1.05<br>(0.95-1.17) | 1.00<br>(0.91-1.09) | 0.425 |

Models were stratified by sex and adjusted for BMI PRS, and the top 10 genetic principal components. Odds ratios (ORs) represent the effect per 1 standard deviation increase in PRS. High genetic risk defined as top quartile (>75th percentile); low genetic risk defined as bottom three quartiles ( ≤ 75th percentile) of PRS. \*p < 0.05, \*\*p < 0.01, \*\*\*p < 0.001.

**Supplementary Table 17:** Sex-stratified blood pressure PRS effects on vascular dementia by WMH genetic risk level.

| Main Effects | Female (OR, 95% CI) |  |  | Male (OR, 95% CI) |  |  |
| --- | --- | --- | --- | --- | --- | --- |
| SBP PRS | 1.04 (0.96-1.13) |  |  | 1.07 (1.00-1.15) |  |  |
| DBP PRS | 1.08 (0.99-1.17) |  |  | 1.02 (0.96-1.10) |  |  |
| Interaction Effects | High WMH PRS | Low WMH PRS | P | High WMH PRS | Low WMH PRS | P |
| **SBP PRS** | 0.97<br>(0.83-1.14) | 1.06<br>(0.97-1.17) | 0.351 | 1.03<br>(0.90-1.17) | 1.09<br>(1.00-1.18) * | 0.466 |
| **DBP PRS** | 1.13<br>(0.96-1.33) | 1.06<br>(0.97-1.17) | 0.516 | 1.01<br>(0.89-1.14) | 1.03<br>(0.95-1.12) | 0.762 |

Models were stratified by sex and adjusted for BMI PRS, and the top 10 genetic principal components. Odds ratios (ORs) represent the effect per 1 standard deviation increase in PRS. High genetic risk defined as top quartile (>75th percentile); low genetic risk defined as bottom three quartiles ( ≤ 75th percentile) of PRS. \*p < 0.05, \*\*p < 0.01, \*\*\*p < 0.001.

**Supplementary Table 18:** Sex-stratified blood pressure PRS effects on all-cause dementia by AD genetic risk level.

| Main Effects | Female (OR, 95% CI) |  |  | Male (OR, 95% CI) |  |  |
| --- | --- | --- | --- | --- | --- | --- |
| **SBP PRS** | 1.05 (1.02-1.09) ** |  |  | 1.05 (1.01-1.08) ** |  |  |
| **DBP PRS** | 1.01 (0.98-1.05) |  |  | 0.99 (0.96-1.02) |  |  |
| Interaction Effects | High AD PRS | Low AD PRS | P | High AD PRS | Low AD PRS | P |
| **SBP PRS** | 1.05<br>(1.00-1.10) | 1.07<br>(1.02-1.13) * | 0.566 | 1.05<br>(1.00-1.10) | 1.04<br>(1.00-1.09) | 0.846 |
| **DBP PRS** | 1.03<br>(0.98-1.08) | 1.00<br>(0.95-1.06) | 0.540 | 0.96<br>(0.91-1.01) | 1.02<br>(0.97-1.07) | 0.066 |

Models were stratified by sex and adjusted for BMI PRS, and the top 10 genetic principal components. Odds ratios (ORs) represent the effect per 1 standard deviation increase in PRS. High genetic risk defined as top quartile (>75th percentile); low genetic risk defined as bottom three quartiles ( ≤ 75th percentile) of PRS. \*p < 0.05, \*\*p < 0.01, \*\*\*p < 0.001.

**Supplementary Table 19:**
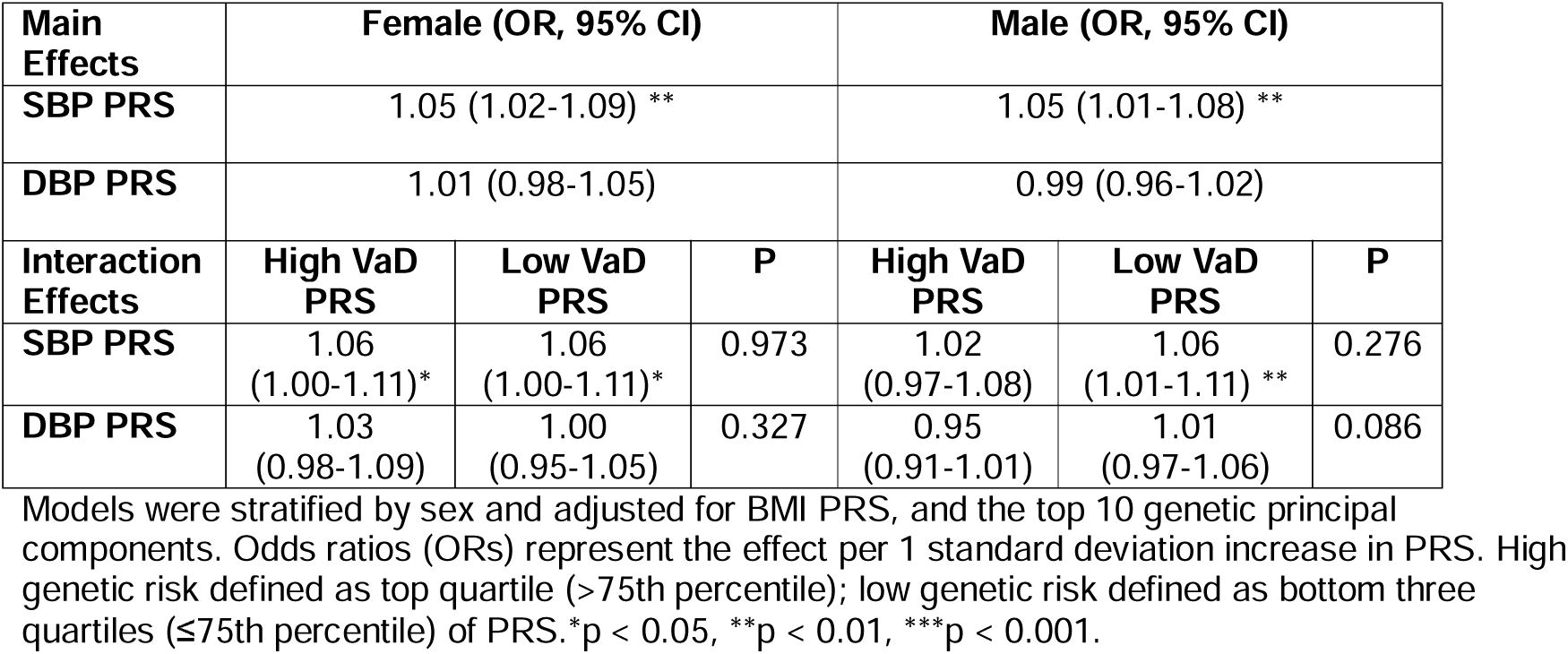
Sex-stratified blood pressure PRS effects on all-cause dementia by VaD genetic risk level.

### Sex-Stratified: Females only, additionally adjusting for HRT use

**Supplementary Table 20:** Blood pressure PRS effects on Alzheimer’s disease by AD genetic risk level.

| Main Effects | Female (OR, 95% CI) |  |  |
| --- | --- | --- | --- |
| SBP | 1.05 (1.00-1.11) * |  |  |
| DBP | 1.01 (0.96-1.06) |  |  |
| Variable | High AD PRS | Low AD PRS | P for interaction |
| SBP PRS | 1.05 (0.99-1.12) | 1.07 (0.98-1.16) | 0.811 |
| DBP PRS | 1.02 (0.95-1.09) | 1.00 (0.92-1.09) | 0.750 |

All models adjusted for age, sex, hormone-replacement therapy (HRT), BMI PRS, and 10 genetic principal components. Values are odds ratios (OR) (95% confidence intervals), that represent the effect per 1 standard deviation increase in SBP or DBP PRS. High genetic risk defined as top quartile (>75th percentile); low genetic risk defined as bottom three quartiles (≤75th percentile). \*p < 0.05, \*\*p < 0.01, \*\*\*p < 0.001.

**Supplementary Table 21:** Blood pressure PRS effects on vascular dementia by VaD genetic risk level.

| Main Effects | Female (OR, 95% CI) |  |  |
| --- | --- | --- | --- |
| SBP | 1.04 (0.96-1.13) |  |  |
| DBP | 1.08 (1.00-1.17) |  |  |
| Variable | High VaD PRS | Low VaD PRS | P for interaction |
| SBP PRS | 1.01 (0.89-1.14) | 1.07 (0.96-1.19) | 0.482 |
| DBP PRS | 1.15 (1.02-1.30) * | 1.03 (0.92-1.15) | 0.176 |

All models adjusted for age, sex, hormone-replacement therapy (HRT), BMI PRS, and 10 genetic principal components. Values are odds ratios (OR) (95% confidence intervals), that represent the effect per 1 standard deviation increase in SBP or DBP PRS. High genetic risk defined as top quartile (>75th percentile); low genetic risk defined as bottom three quartiles (≤75th percentile). \*p < 0.05, \*\*p < 0.01, \*\*\*p < 0.001.

**Supplementary Table 22:** Blood pressure PRS effects on vascular dementia by WMH genetic risk level.

| Main Effects | Female (OR, 95% CI) |  |  |
| --- | --- | --- | --- |
| SBP | 1.04 (0.96-1.13) |  |  |
| DBP | 1.08 (1.00-1.17) |  |  |
| Variable | High WMH PRS | Low WMH PRS | P for interaction |
| **SBP PRS** | 0.97 (0.83-1.14) | 1.07 (0.97-1.17) | 0.316 |
| **DBP PRS** | 1.13 (0.96-1.32) | 1.06 (0.97-1.17) | 0.537 |

All models adjusted for age, sex, hormone-replacement therapy (HRT), BMI PRS, and 10 genetic principal components. Values are odds ratios (OR) (95% confidence intervals), that represent the effect per 1 standard deviation increase in SBP or DBP PRS. High genetic risk defined as top quartile (>75th percentile); low genetic risk defined as bottom three quartiles ( ≤ 75th percentile). \*p < 0.05, \*\*p < 0.01, \*\*\*p < 0.001.

**Supplementary Table 23:** Blood pressure PRS effects on all-cause dementia by AD genetic risk level.

| Main Effects | Female (OR, 95% CI) |  |  |
| --- | --- | --- | --- |
| **SBP** | 1.05 (1.02-1.09) ** |  |  |
| **DBP** | 1.01 (0.98-1.05) |  |  |
| Variable | High AD PRS | Low AD PRS | P for interaction |
| **SBP PRS** | 1.04 (0.99-1.10) | 1.07 (1.02-1.13) ** | 0.460 |
| **DBP PRS** | 1.03 (0.98-1.08) | 1.01 (0.96-1.06) | 0.546 |

All models adjusted for age, sex, hormone-replacement therapy (HRT), BMI PRS, and 10 genetic principal components. Values are odds ratios (OR) (95% confidence intervals), that represent the effect per 1 standard deviation increase in SBP or DBP PRS. High genetic risk defined as top quartile (>75th percentile); low genetic risk defined as bottom three quartiles ( ≤ 75th percentile). \*p < 0.05, \*\*p < 0.01, \*\*\*p < 0.001.

**Supplementary Table 24:** Blood pressure PRS effects on all-cause dementia by VaD genetic risk level.

| Main Effects | Female (OR, 95% CI) |  |  |
| --- | --- | --- | --- |
| **SBP** | 1.05 (1.02-1.09) ** |  |  |
| **DBP** | 1.01 (0.98-1.05) |  |  |
| Variable | High VaD PRS | Low VaD PRS | P for interaction |
| **SBP PRS** | 1.05 (1.00-1.11) | 1.06 (1.01-1.11) | 0.931 |
| **DBP PRS** | 1.03 (0.98-1.09) | 1.00 (0.95-1.05) | 0.355 |

All models adjusted for age, sex, hormone-replacement therapy (HRT), BMI PRS, and 10 genetic principal components. Values are odds ratios (OR) (95% confidence intervals), that represent the effect per 1 standard deviation increase in SBP or DBP PRS. High genetic risk defined as top quartile (>75th percentile); low genetic risk defined as bottom three quartiles ( ≤ 75th percentile). \*p < 0.05, \*\*p < 0.01, \*\*\*p < 0.001.

**Supplementary Table 25:**
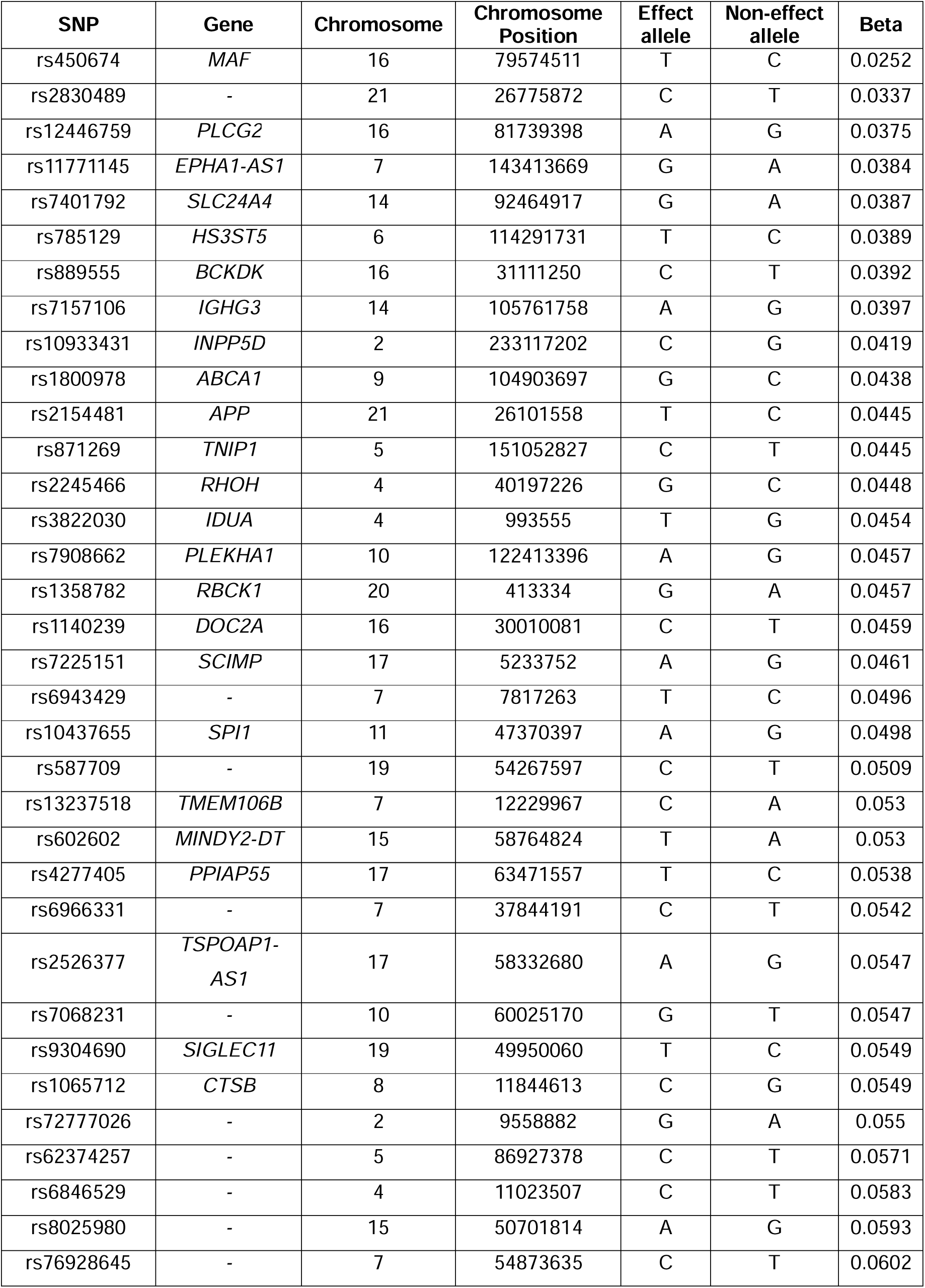
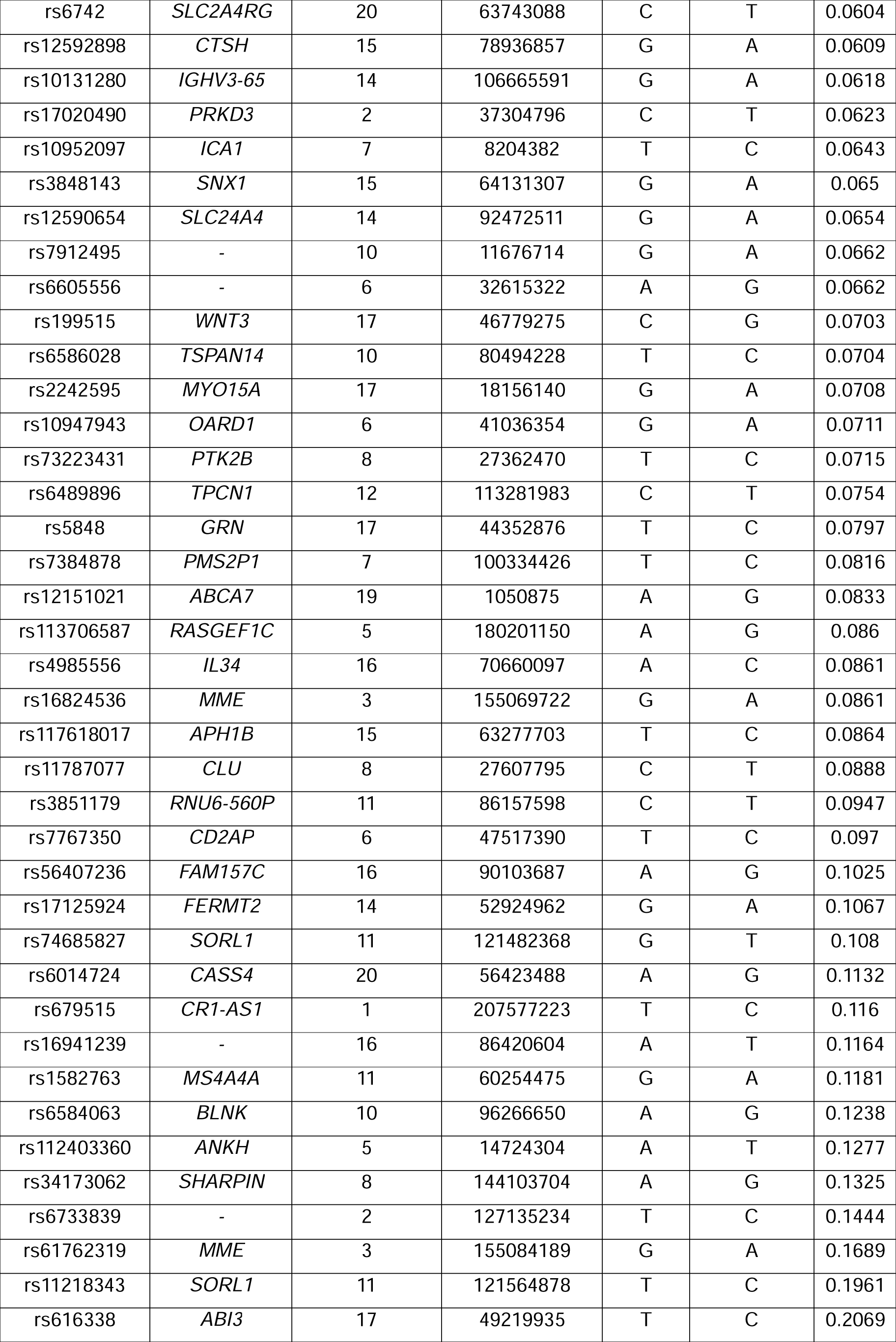
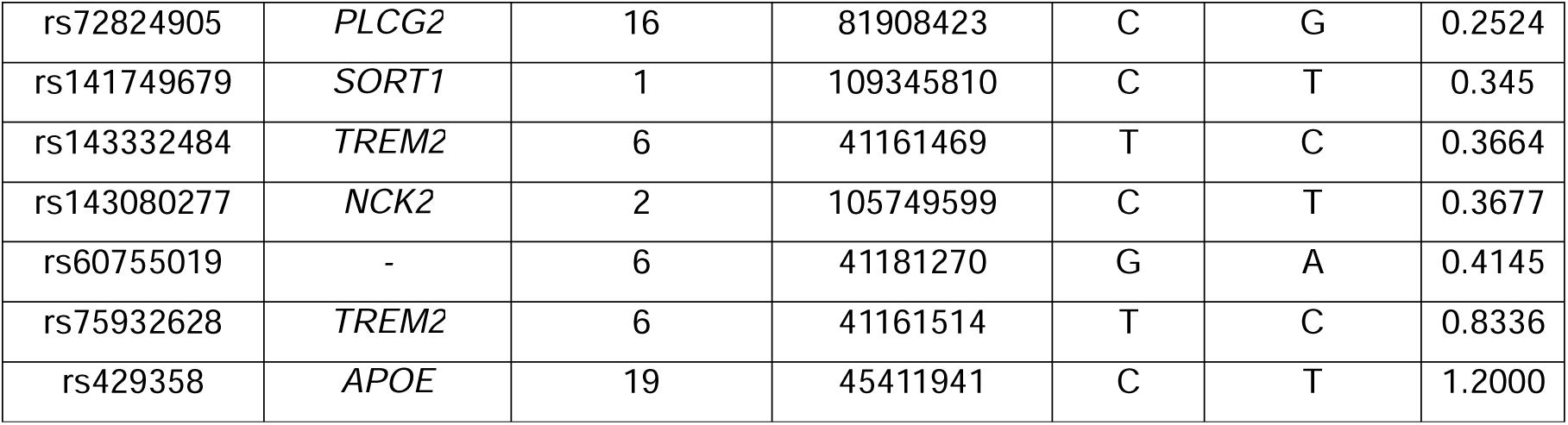
Genetic variants and weights used to construct Alzheimer’s disease PRS.

**Supplementary Table 26:**
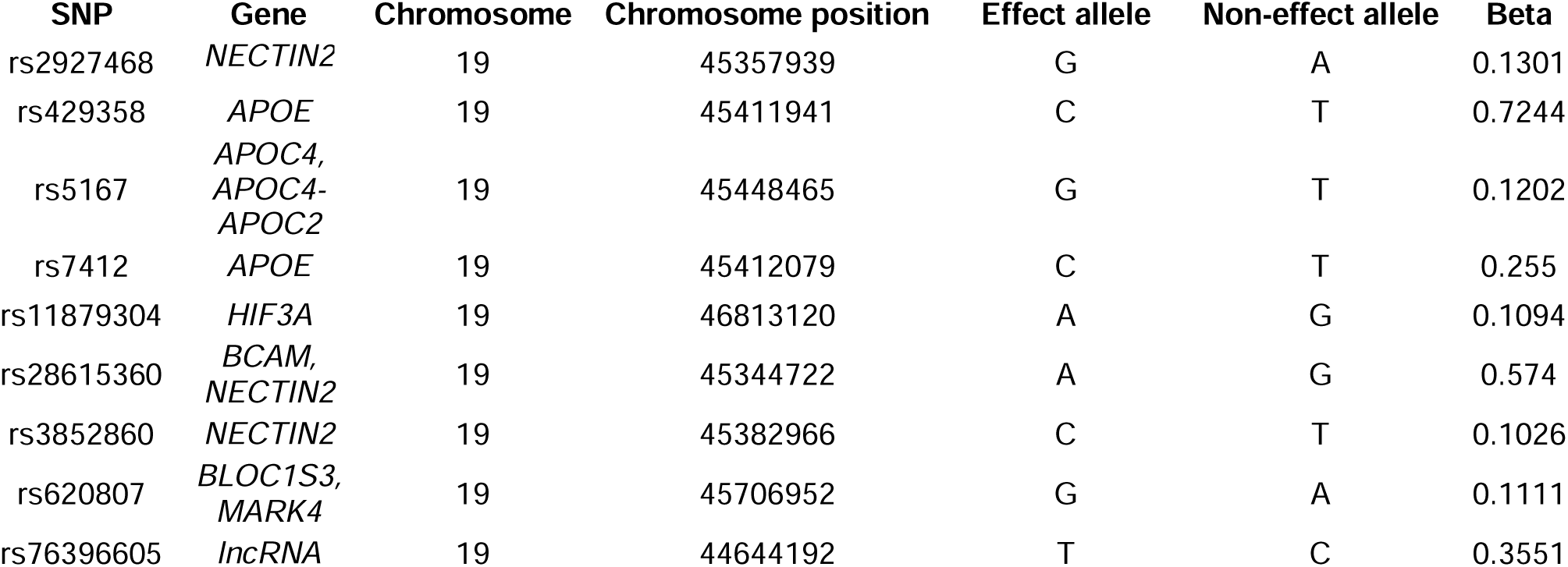
Genetic variants and weights used to construct vascular dementia PRS.

**Supplementary Table 27.** Genetic variants and weights used to construct white matter hyperintensities PRS.

| SNP | Gene | Chromosome | Chromosome position | Effect allele | Non-effect allele | Beta |
| --- | --- | --- | --- | --- | --- | --- |
| rs6503417 | *NMT1* | 17 | 43144218 | C | T | 0.052 |
| rs12921170 | *C16orf95* | 16 | 87227397 | A | G | 0.05 |
| rs6797002 | *KLHL24* | 3 | 18336326 | C | T | 0.049 |
| rs17205972 | *VCAN-*<br>*AS1* | 5 | 82859065 | T | G | 0.049 |
| rs62172472 | *CALCRL-*<br>*AS1* | 2 | 18802831 | G | A | 0.047 |
| rs2303655 | *ZNF475* | 5 | 12151837 | T | C | 0.048 |
| rs1948948 | - | 16 | 51442679 | C | T | 0.037 |
| rs1285847 | *CCDC88*<br>*C-DT* | 14 | 91884655 | T | C | 0.036 |
| rs73184312 | *PRAG1* | 8 | 8179639 | G | A | 0.038 |
| rs71471298 | *SH3PXD*<br>*2A-AS1* | 10 | 10550714 | T | C | 0.053 |
| rs5762197 | - | 22 | 27887471 | C | A | 0.039 |
| rs12443113 | *RASL12* | 15 | 65355468 | G | A | 0.031 |
| rs11249945 | *TNKS* | 8 | 9628753 | A | G | 0.034 |
| rs72680374 | *LINC02319* | 14 | 52604843 | T | A | 0.032 |
| rs7004825 | *XKR6* | 8 | 11031472 | T | C | 0.031 |
| rs786921 | *PKN2* | 1 | 89286673 | A | G | 0.033 |
| rs34974290 | *TRIM65* | 17 | 73888354 | A | G | 0.104 |
| rs7596872 | *EFEMP1* | 2 | 56128091 | A | C | 0.097 |
| rs6940540 | *PLEKHG1* | 6 | 15101890 | G | T | 0.045 |
| rs73923006 | - | 2 | 43132224 | G | C | 0.055 |
| rs4630220 | - | 10 | 10545911 | G | A | 0.048 |
| rs7603972 | *CARF* | 2 | 20378051 | A | G | 0.07 |
| rs10786772 | *SH3PXD2A* | 10 | 10561032 | G | A | 0.042 |
| rs55940034 | *COL4A2* | 13 | 11104330 | G | A | 0.037 |
| rs7157599 | - | 14 | 10062590 | C | T | 0.041 |

**Supplementary Table 28.**
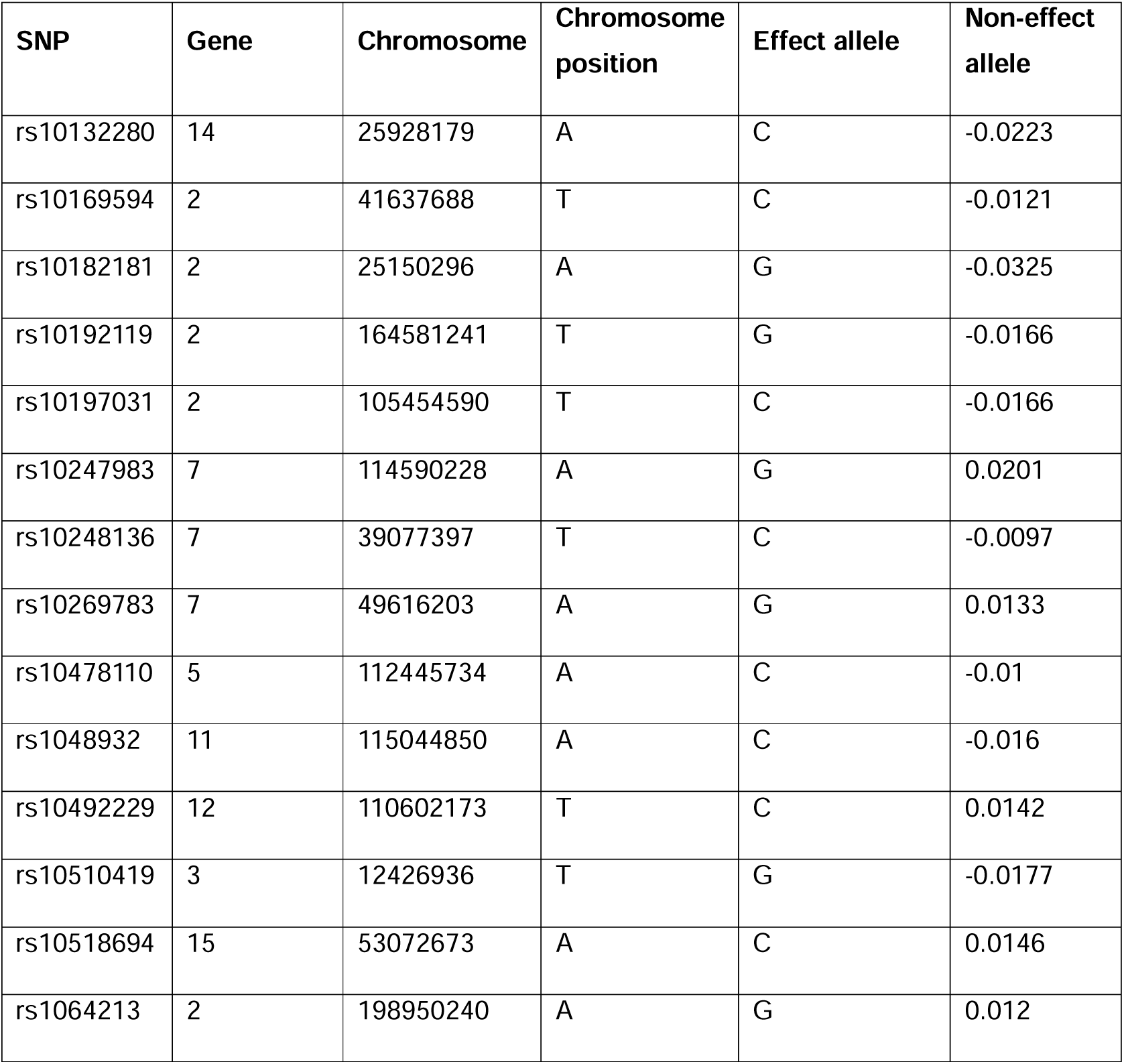
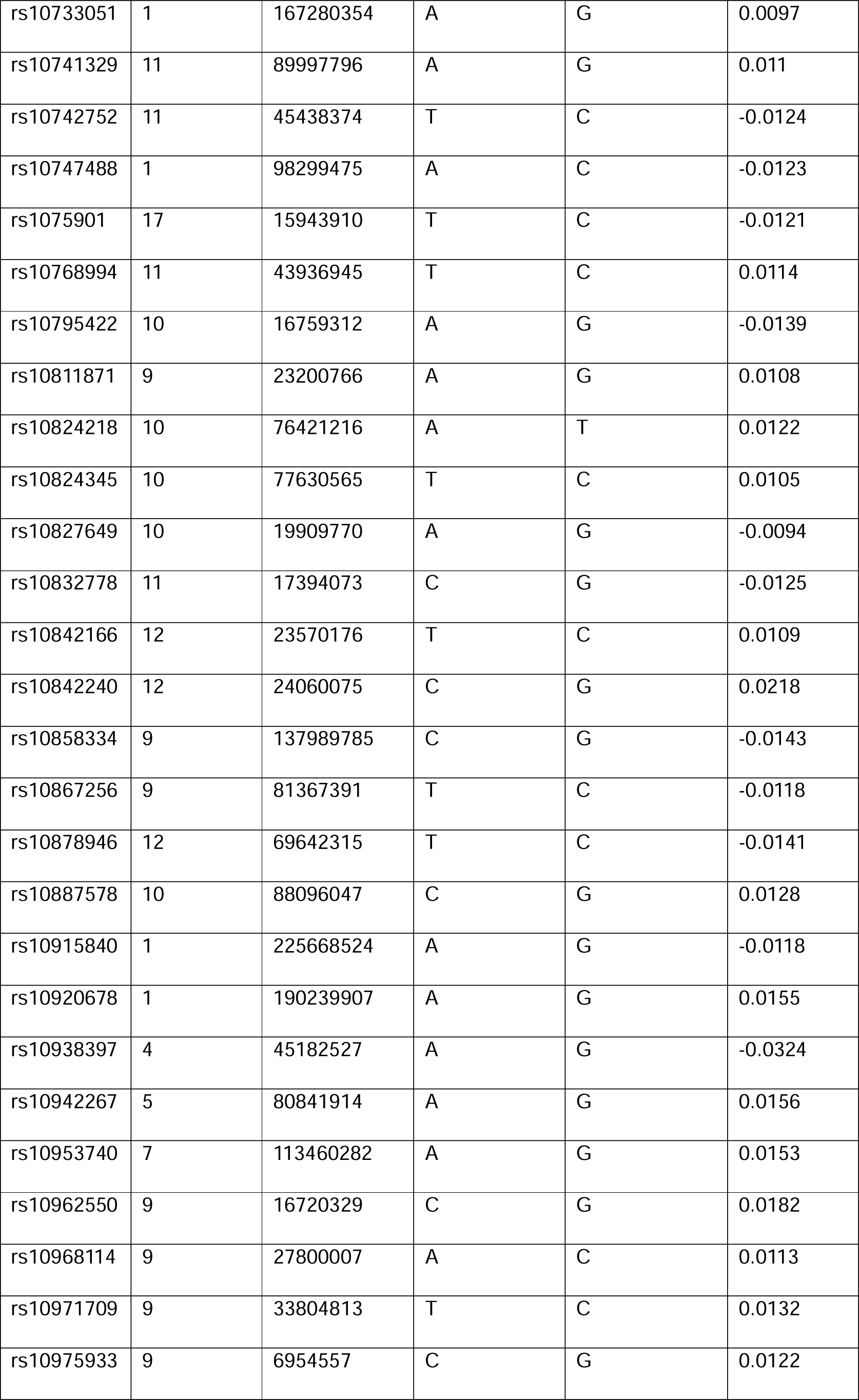
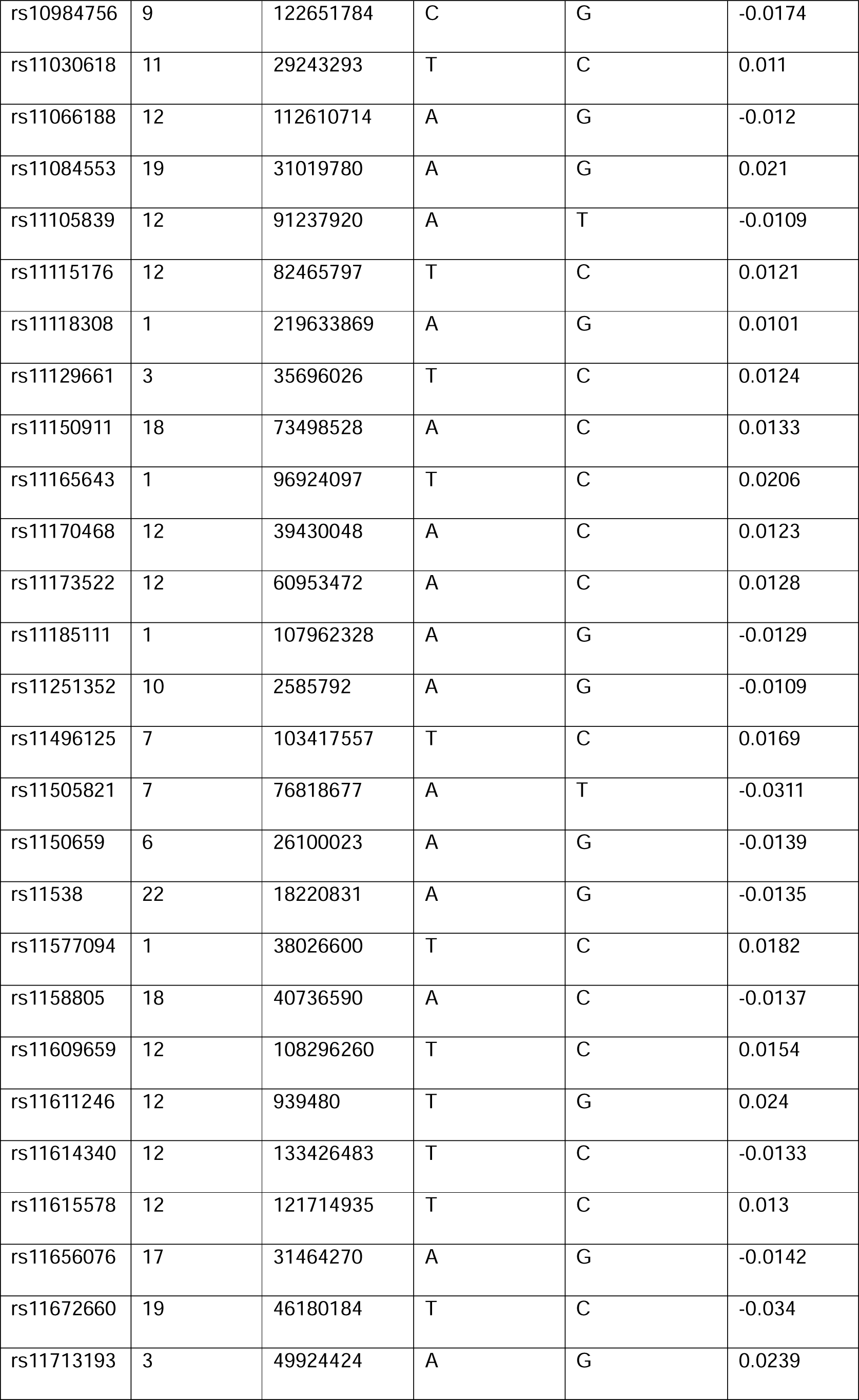
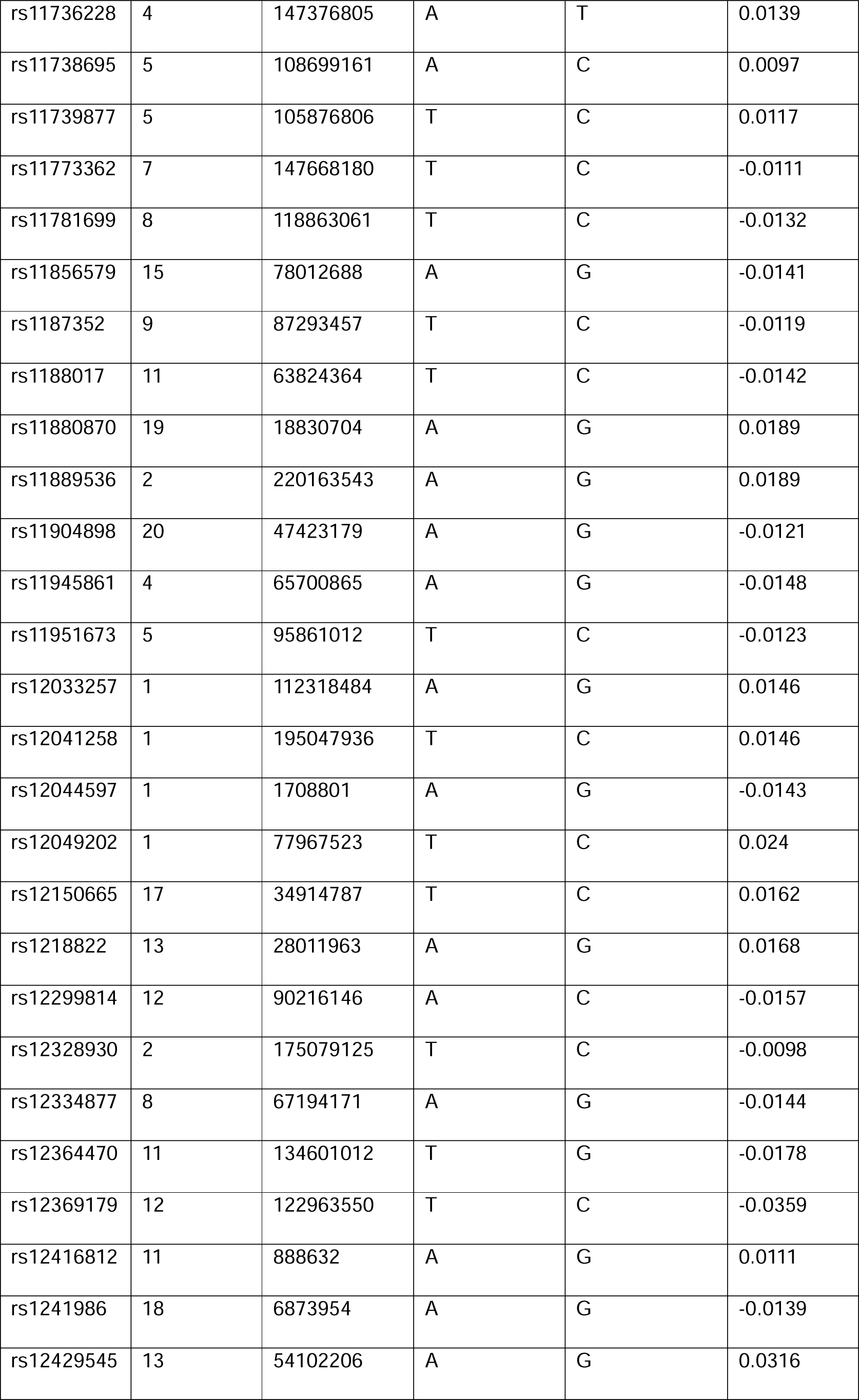
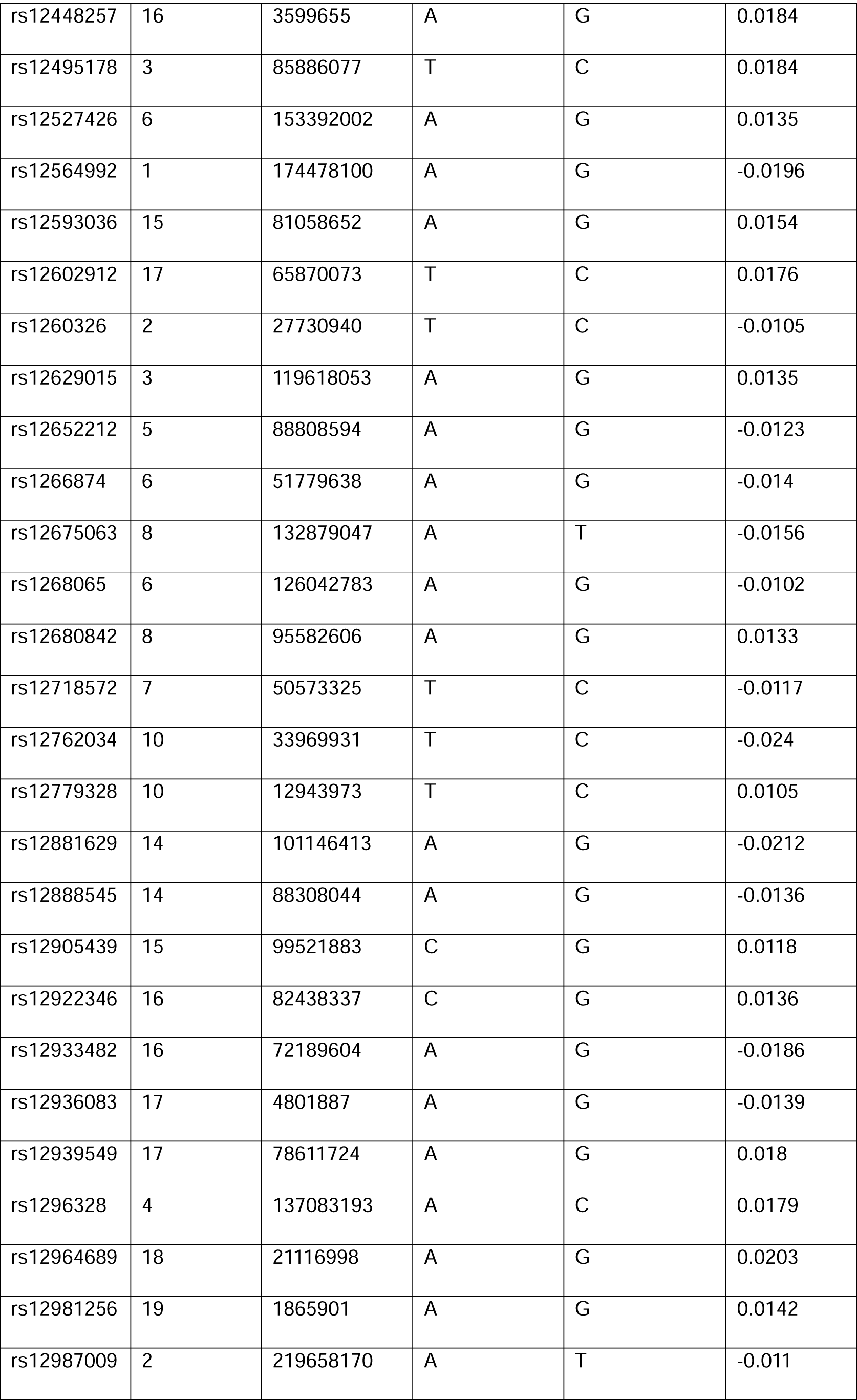
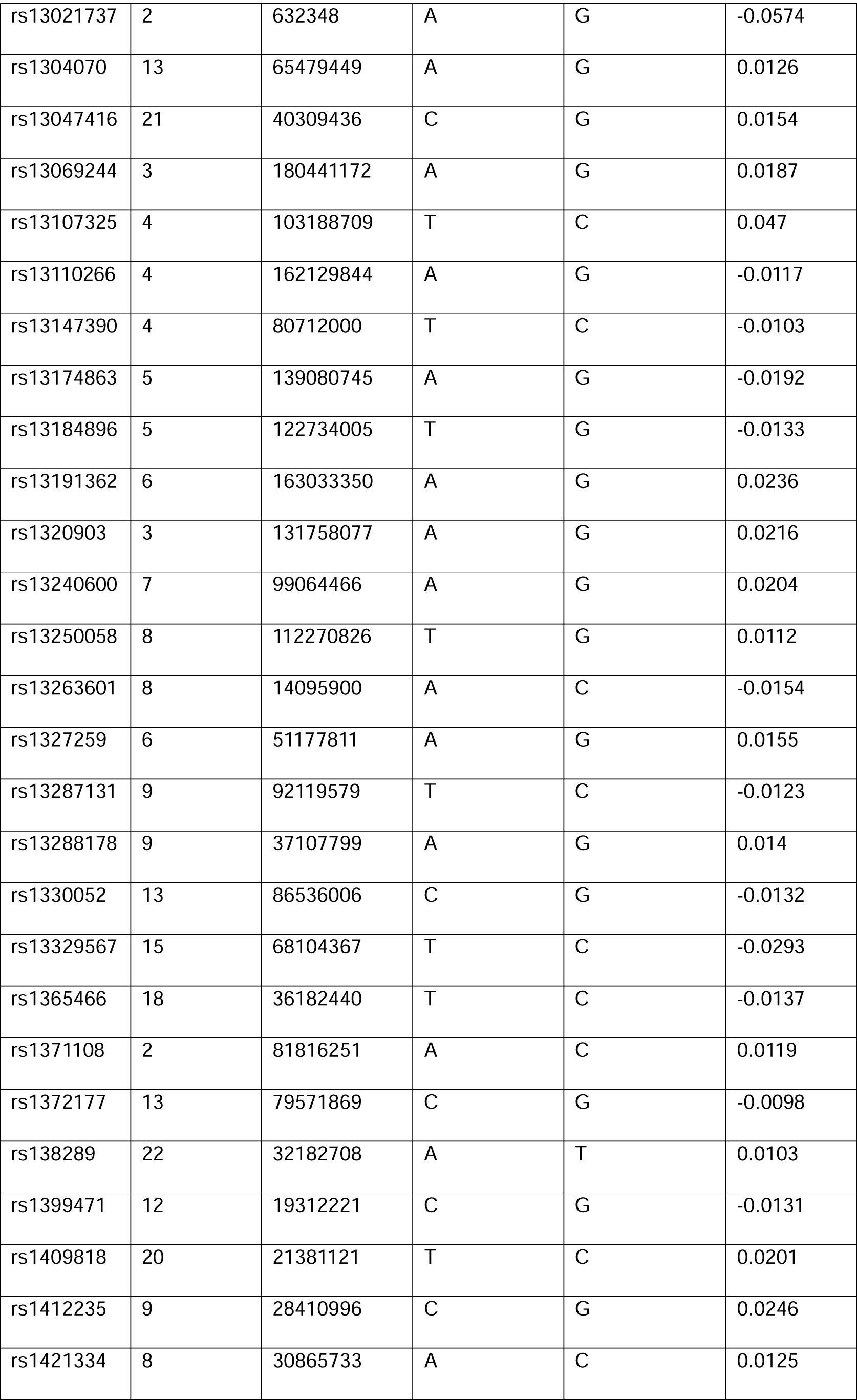
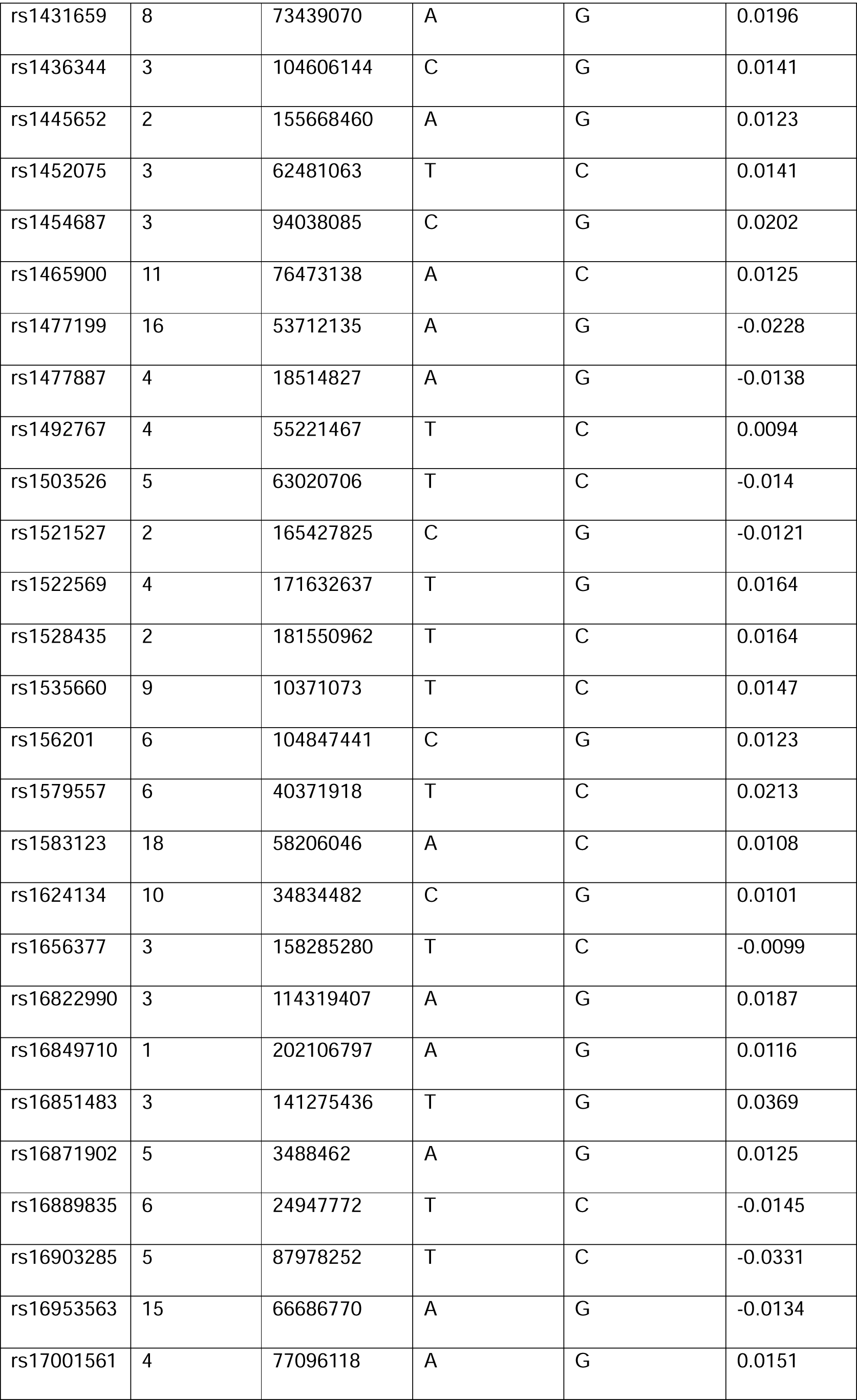
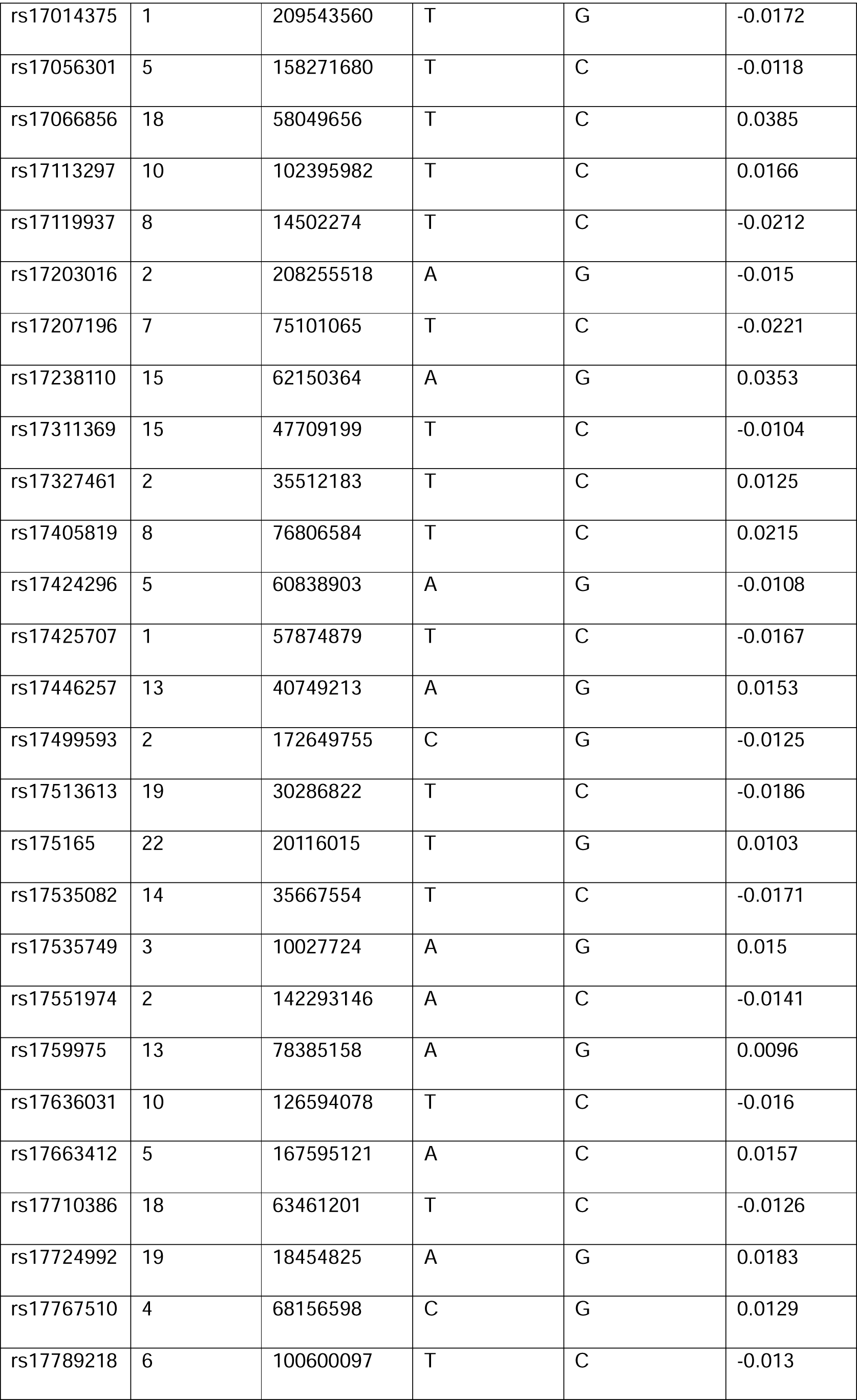
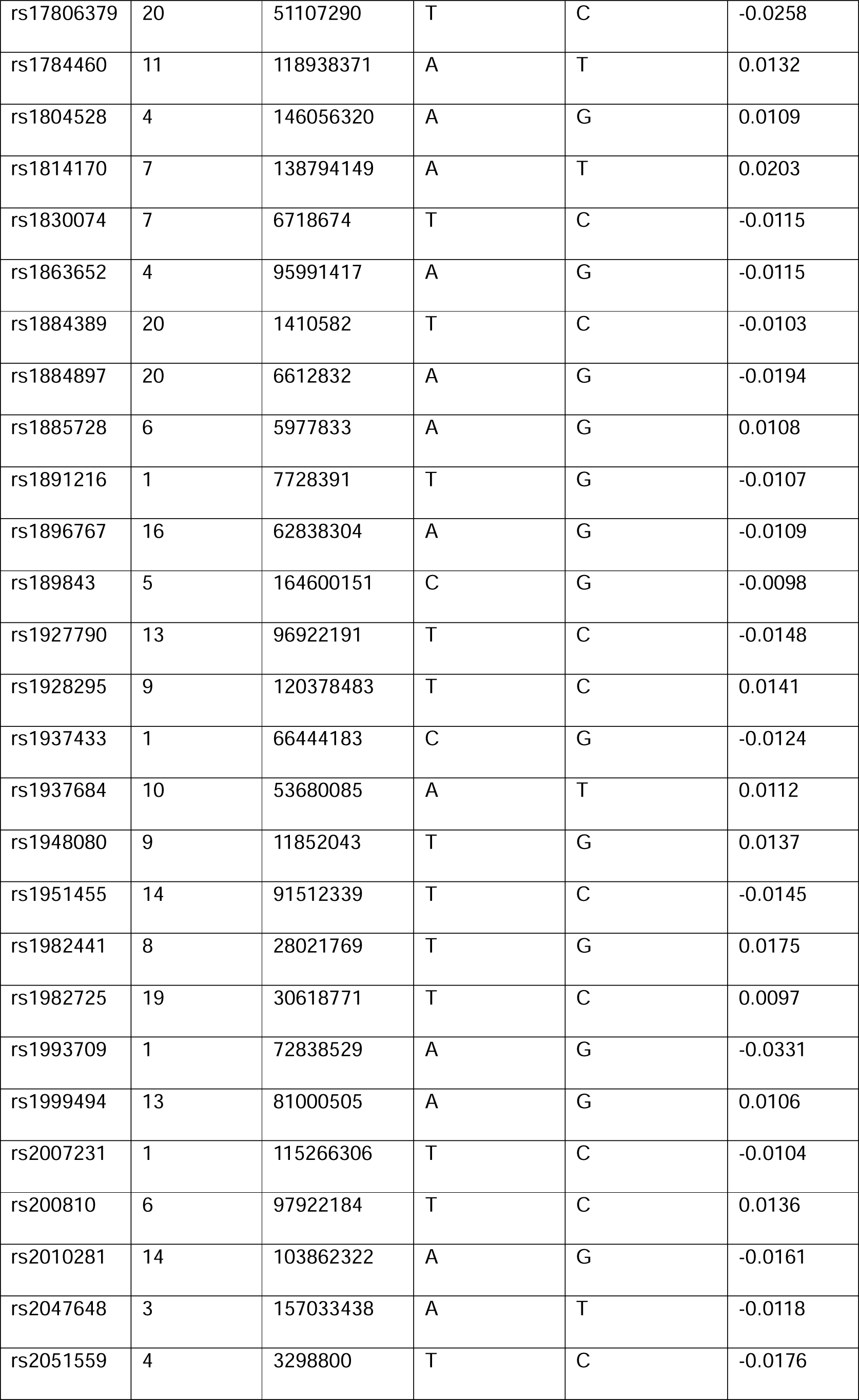
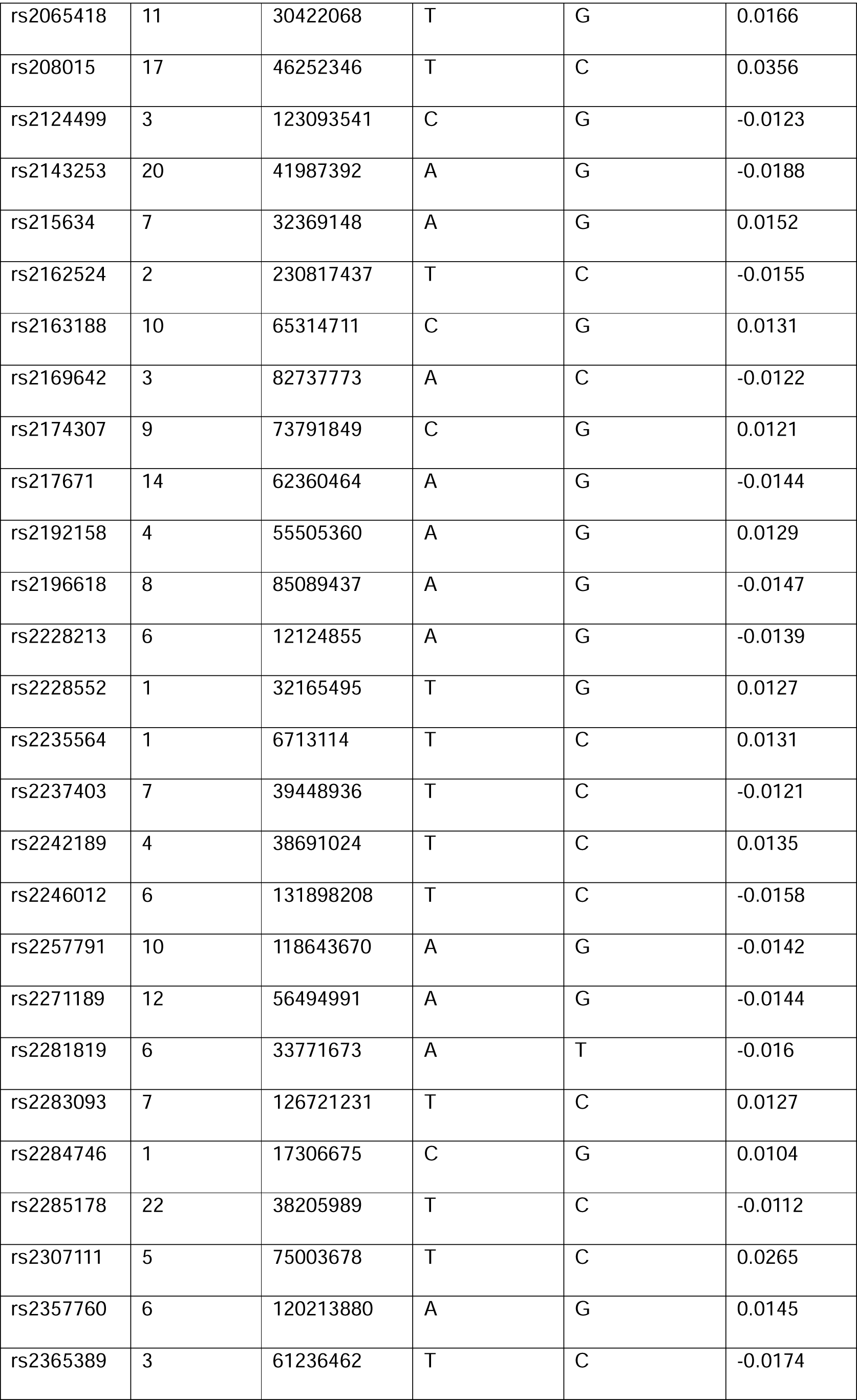
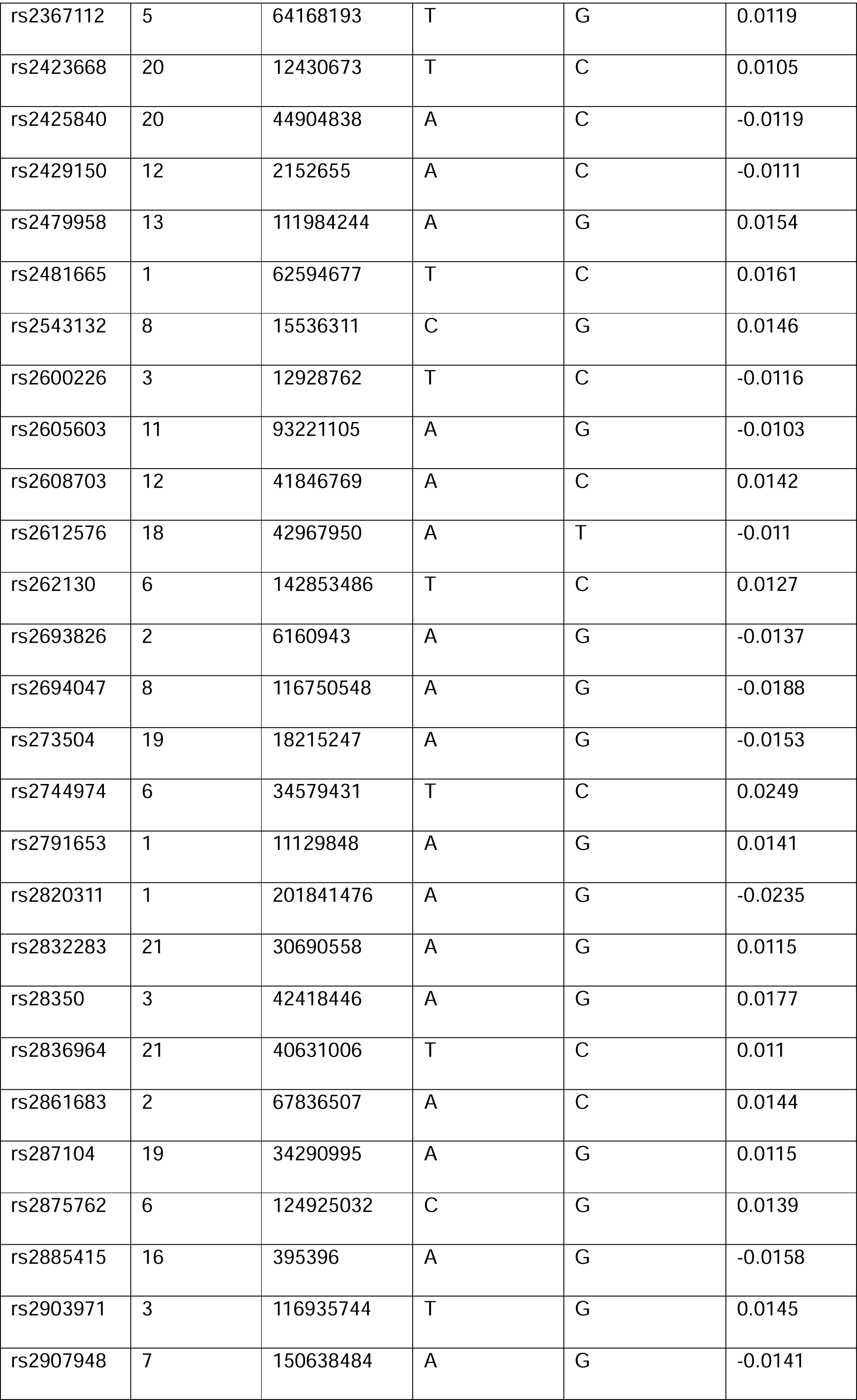
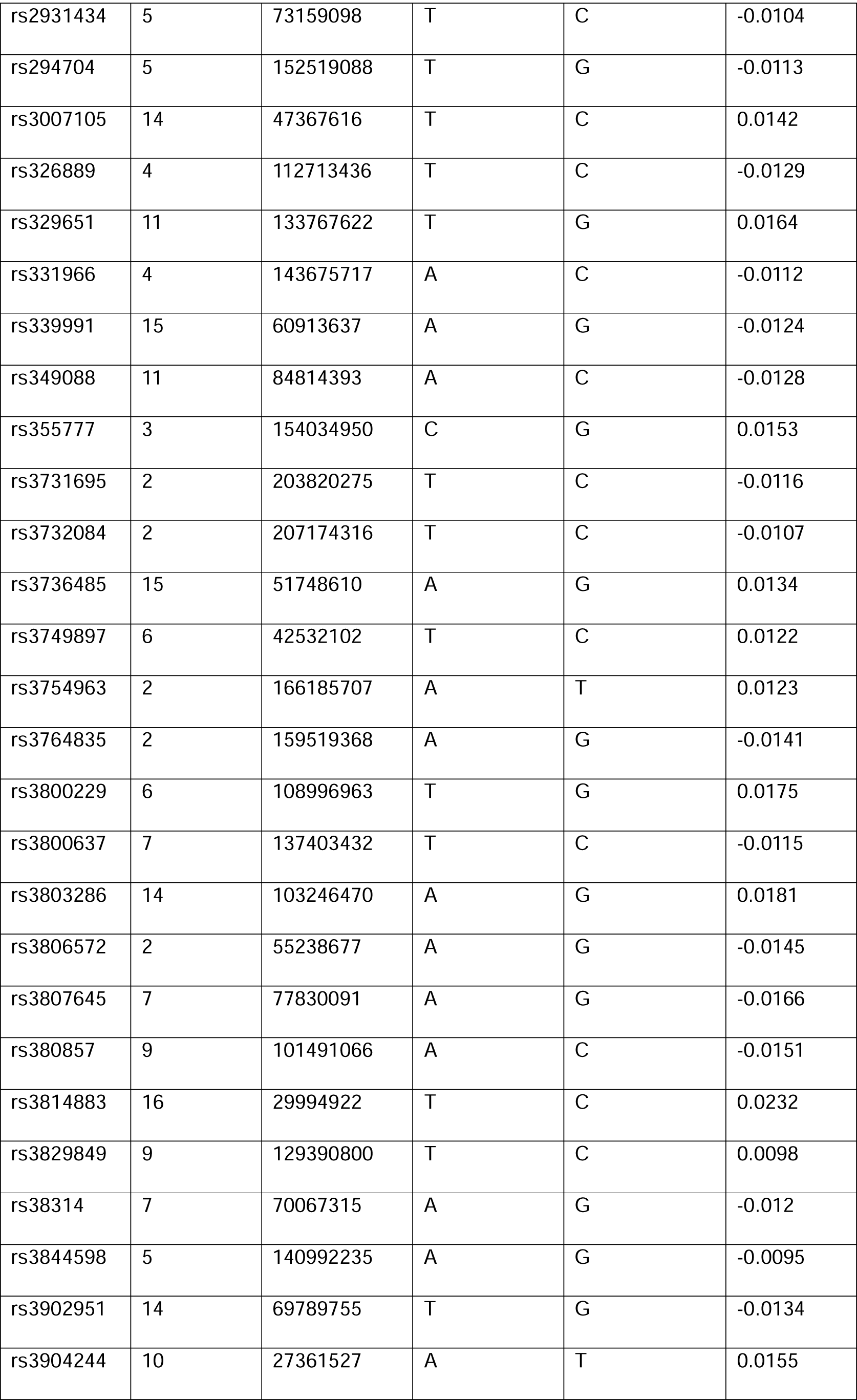
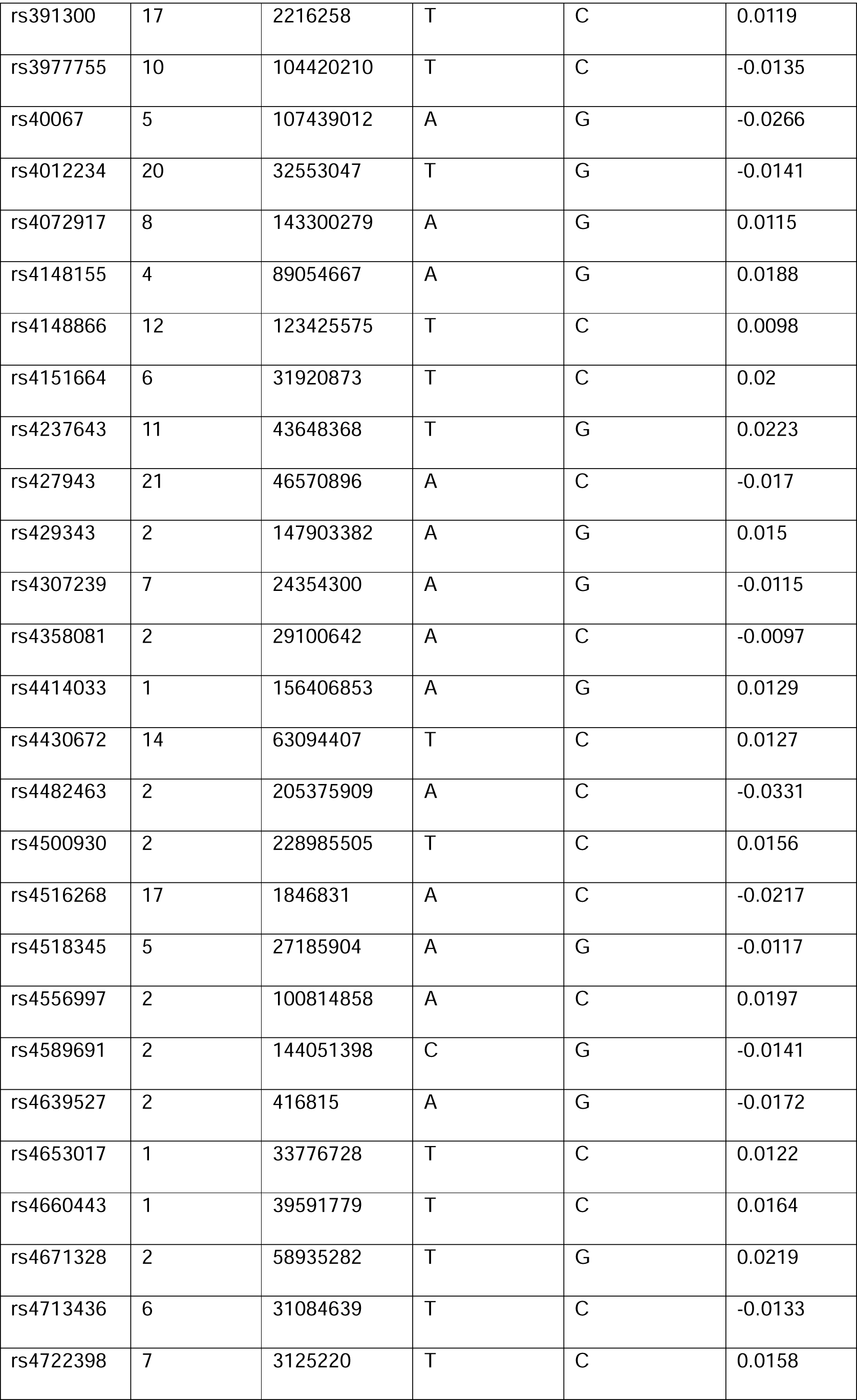
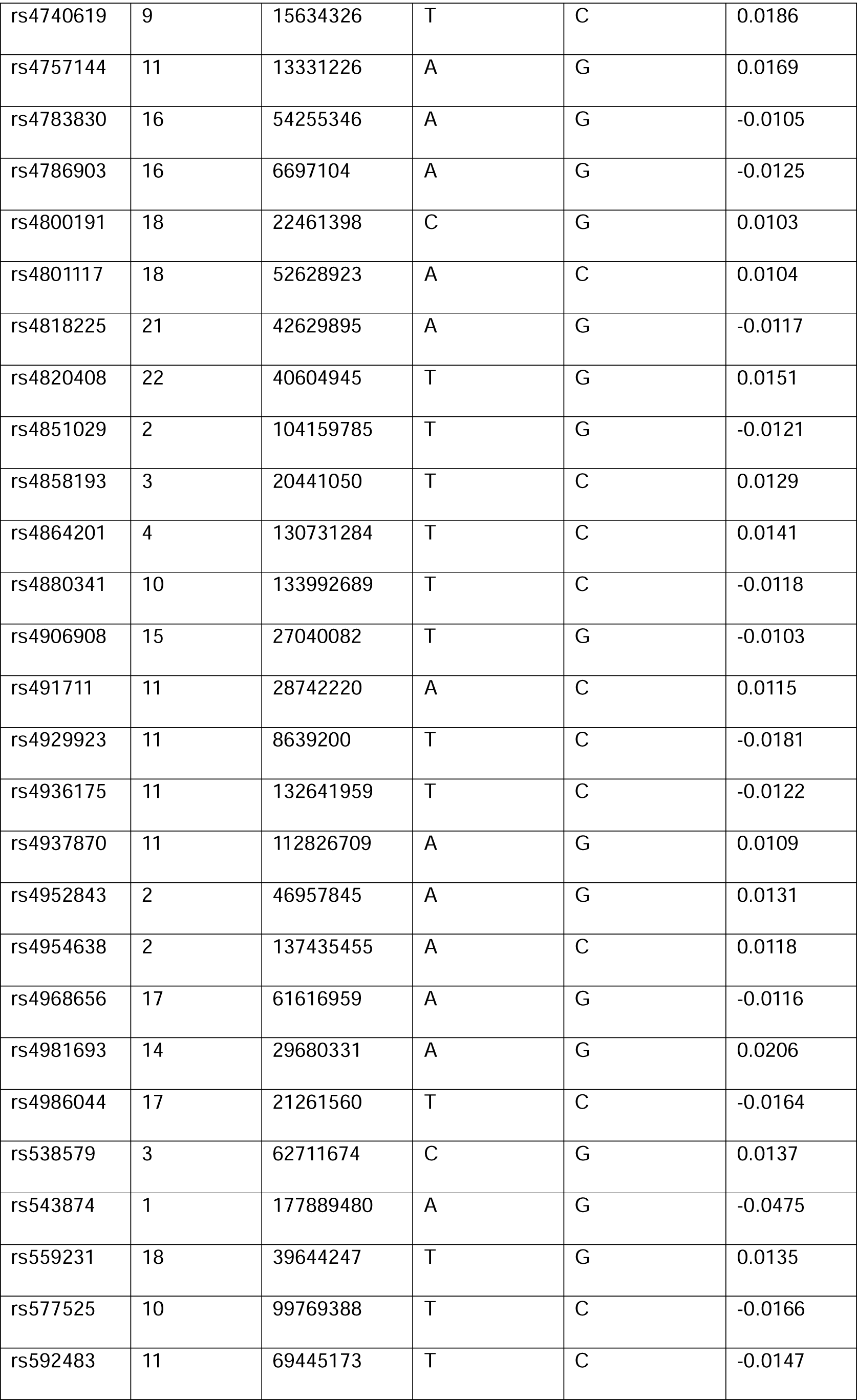
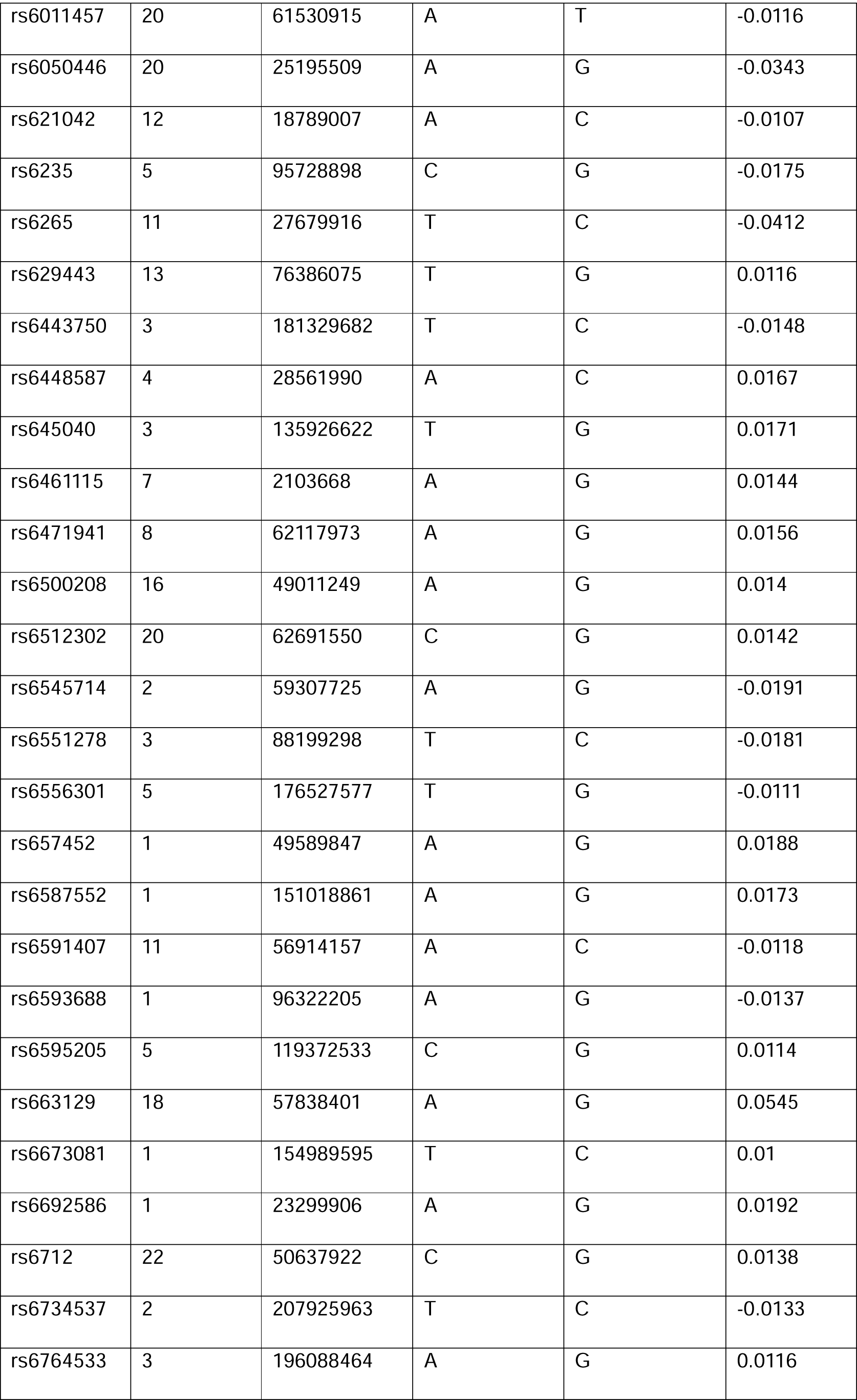
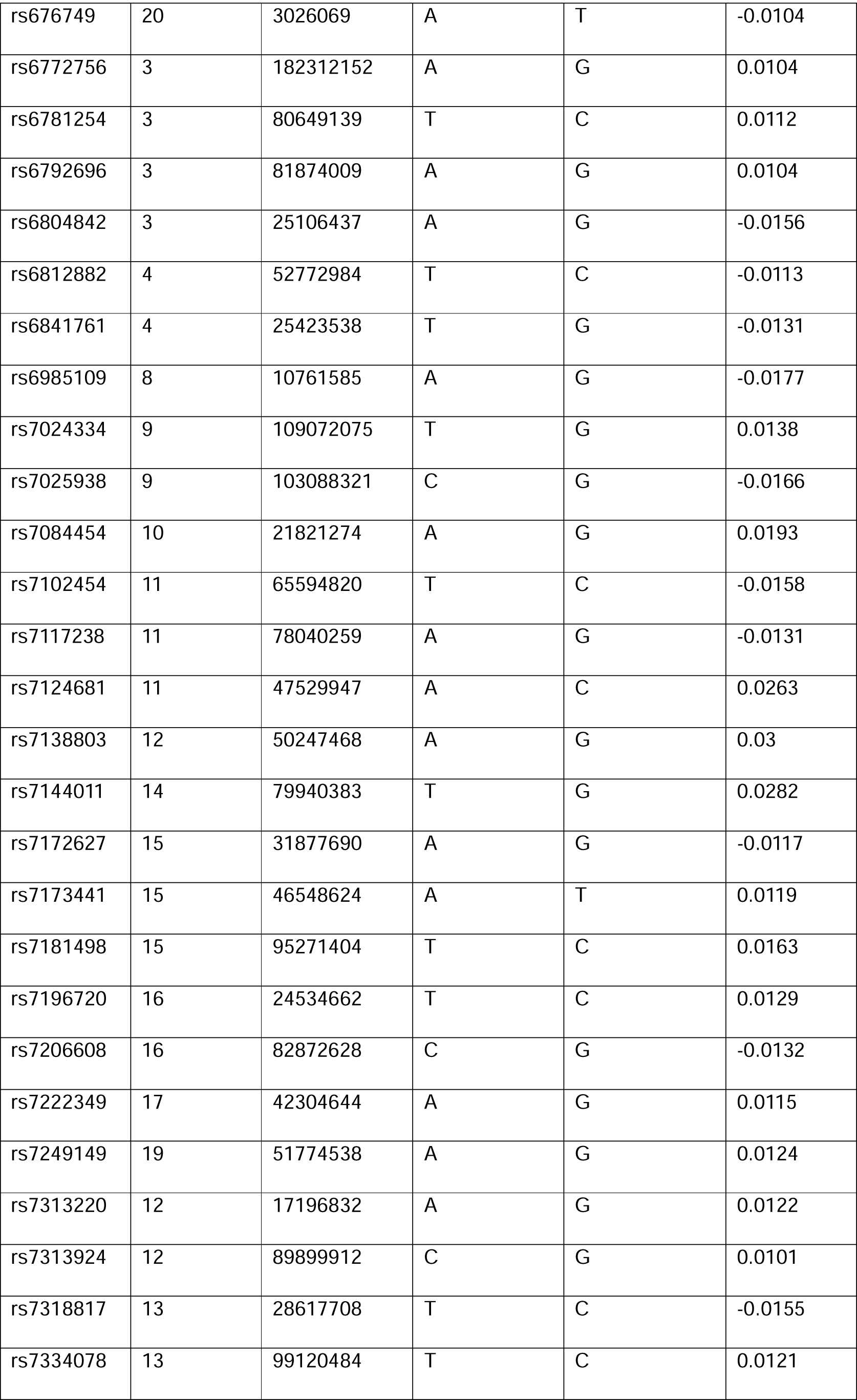
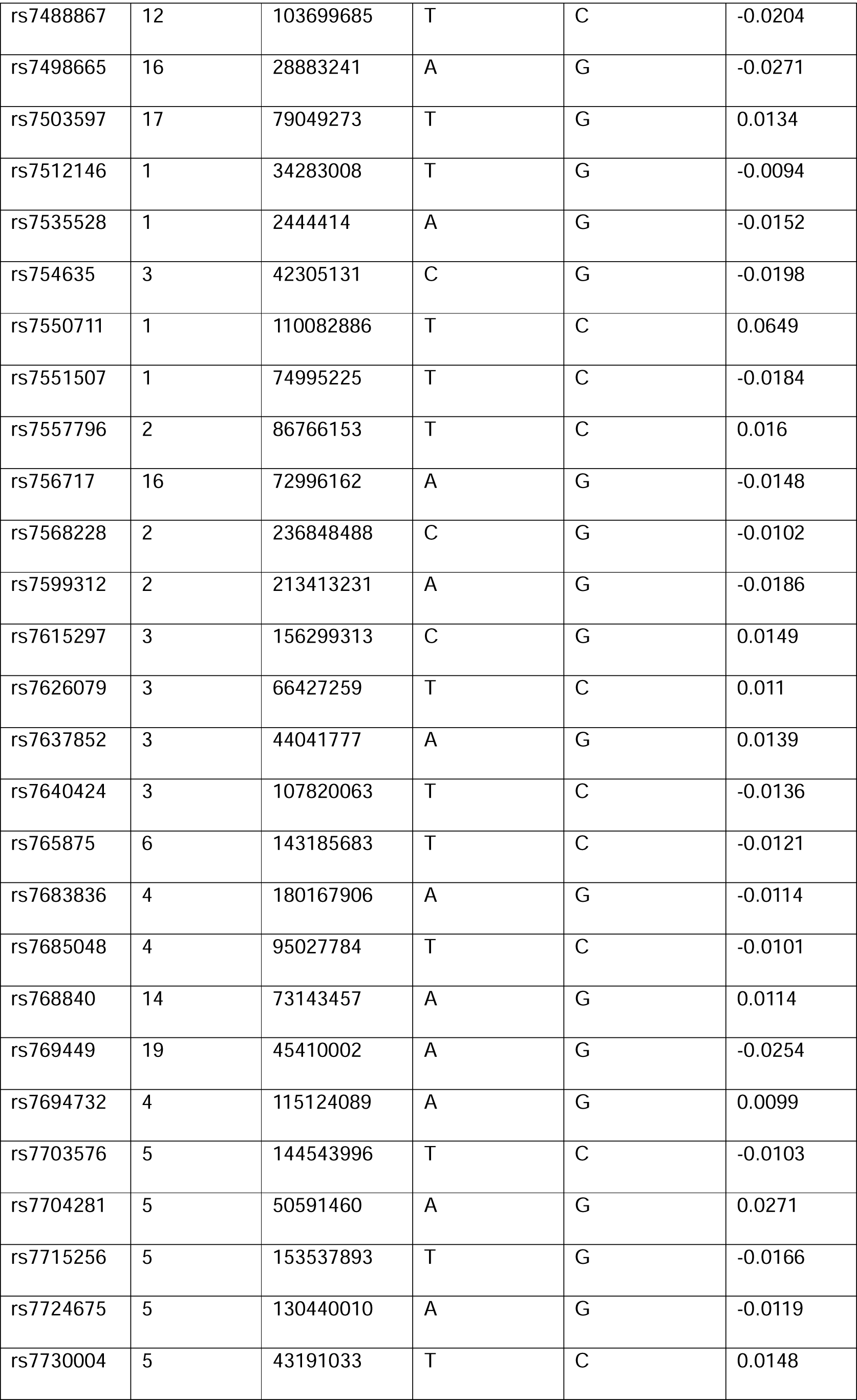
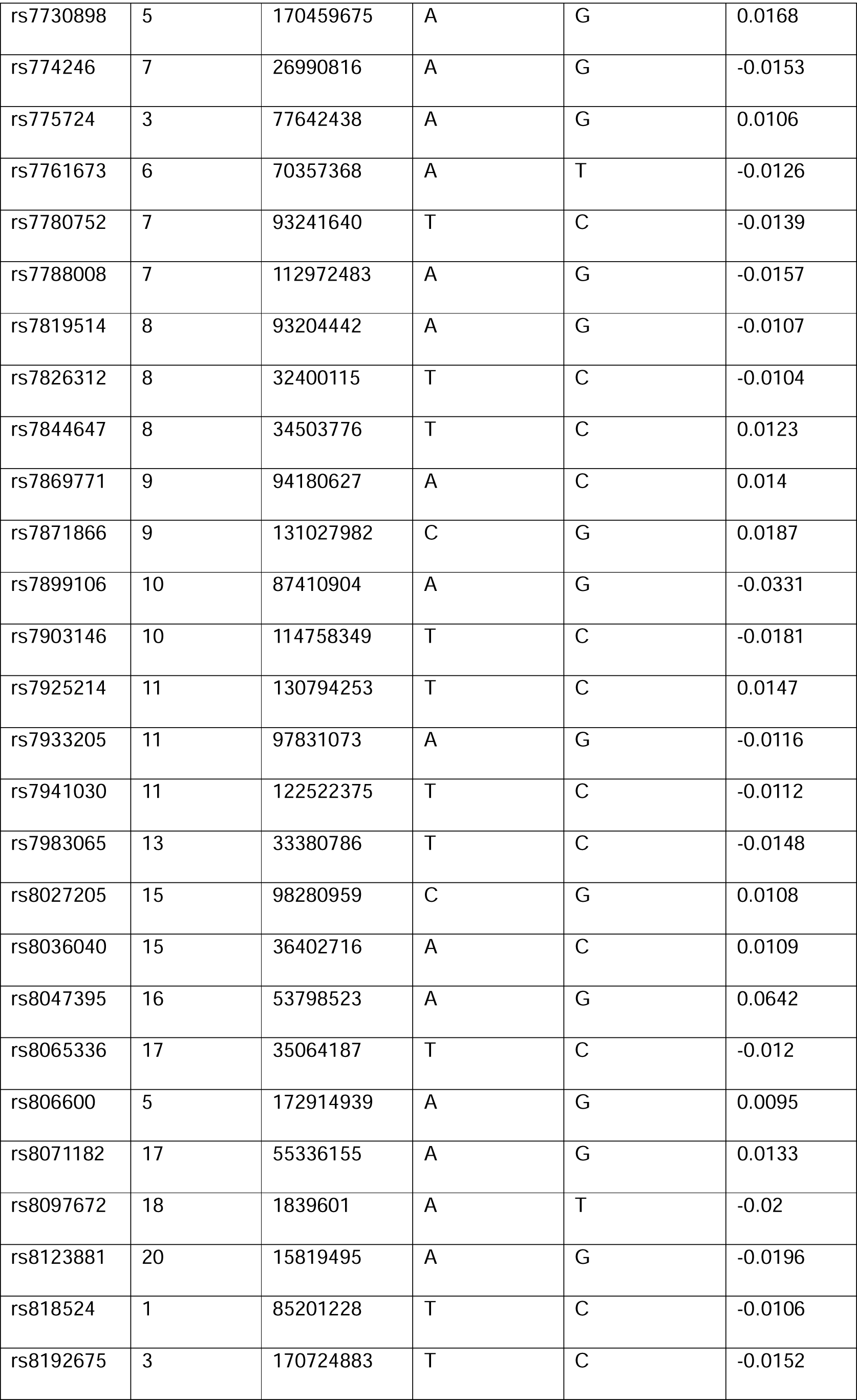
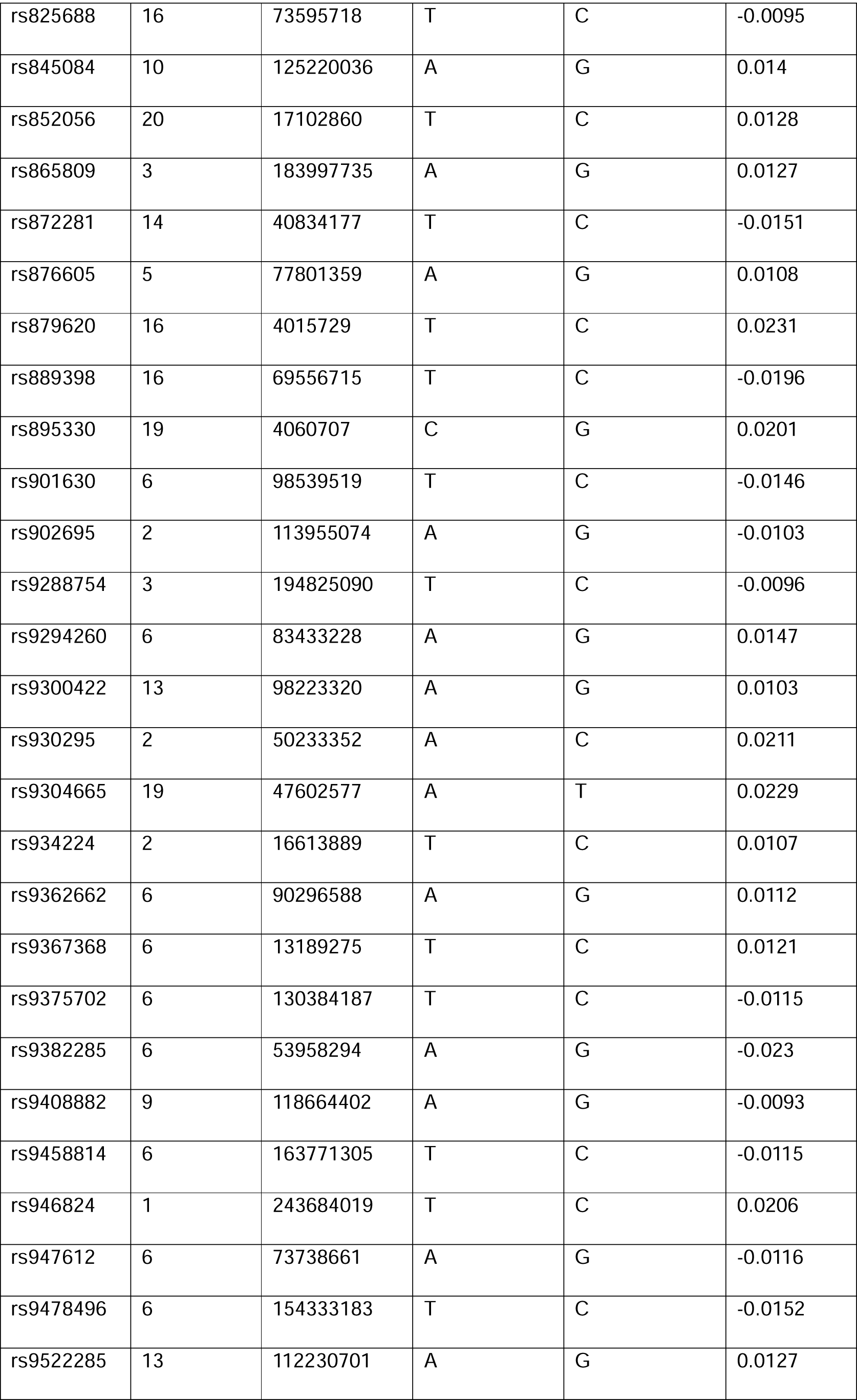
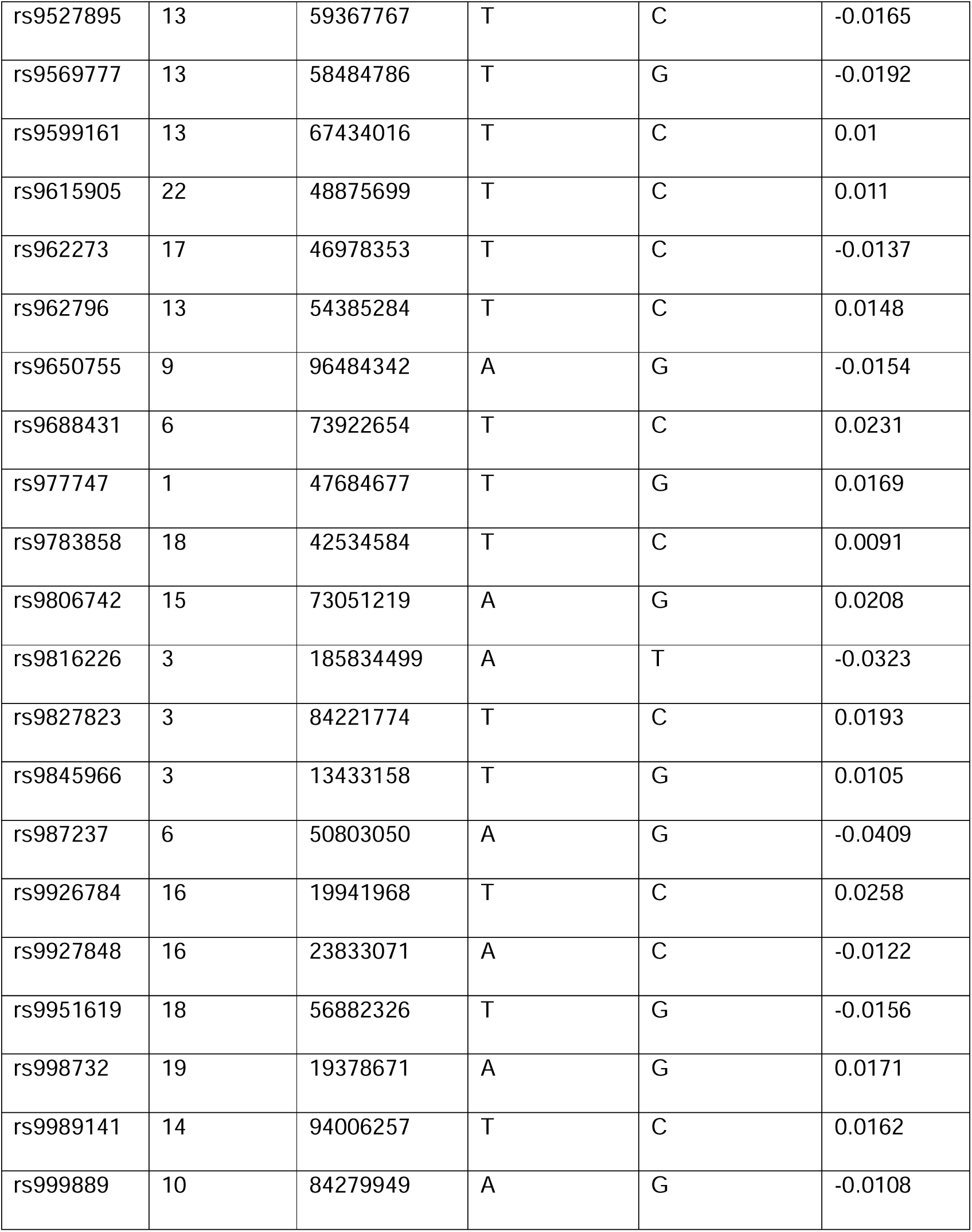
521 genome-wide significant genetic variants and weights used to construct BMI PRS (p < 5×10^-8^, R^2^ < 0.001, >10,000kb)

**Supplementary Table 29.** Genetic variants and weights used to construct systolic blood pressure PRS.

| SNP | Chromosome | Chromosome position | Effect allele | Non-effect allele | Beta |
| --- | --- | --- | --- | --- | --- |
| rs6667998 | 1 | 1159540 | A | G | -0.335 |
| rs260508 | 1 | 2187085 | T | G | 0.1485 |
| rs11122123 | 1 | 6791386 | A | T | -0.1365 |
| rs12724366 | 1 | 20798620 | A | G | 0.182 |
| rs2807337 | 1 | 22577371 | T | C | 0.1753 |
| rs79598313 | 1 | 27284913 | T | C | 0.5926 |
| rs3737801 | 1 | 27960832 | C | G | 0.2984 |
| rs34393348 | 1 | 39328203 | A | C | 0.1154 |
| rs11210029 | 1 | 41865293 | A | G | -0.1484 |
| rs839755 | 1 | 43856410 | A | C | -0.2274 |
| rs11579440 | 1 | 49052423 | T | C | 0.2117 |
| rs12063422 | 1 | 54585415 | T | C | -0.1213 |
| rs4439379 | 1 | 61782375 | A | C | 0.15 |
| rs10747396 | 1 | 82954952 | T | C | 0.1357 |
| rs10923038 | 1 | 88651771 | A | C | 0.1625 |
| rs7514579 | 1 | 94051350 | A | C | 0.1969 |
| rs17396055 | 1 | 94730954 | A | G | -0.1727 |
| rs4604740 | 1 | 95789603 | T | C | -0.108 |
| rs2797536 | 1 | 111193044 | A | G | -0.1261 |
| rs2786989 | 1 | 112148362 | T | G | 0.1926 |
| rs1707111 | 1 | 117870085 | C | G | 0.122 |
| rs76719272 | 1 | 156129796 | T | C | -0.2394 |
| rs4573493 | 1 | 166023209 | A | G | -0.1114 |
| rs1043069 | 1 | 180859368 | T | G | 0.1995 |
| rs6684423 | 1 | 181560367 | T | G | 0.191 |
| rs4651224 | 1 | 184585182 | T | C | 0.1944 |
| rs1007460 | 1 | 193510827 | T | C | -0.1287 |
| rs12042924 | 1 | 197297417 | T | C | -0.1615 |
| rs33996239 | 1 | 203109801 | T | C | -0.3538 |
| rs7555285 | 1 | 209970355 | C | G | 0.1961 |
| rs504130 | 1 | 244166416 | A | G | 0.0978 |
| rs4926499 | 1 | 249155909 | C | G | 0.2954 |
| rs67720684 | 2 | 18975439 | A | C | 0.2039 |
| rs35590893 | 2 | 43716933 | A | G | -0.2152 |
| rs4586678 | 2 | 44219937 | A | T | -0.1482 |
| rs6545155 | 2 | 50429861 | T | C | 0.1691 |
| rs10189186 | 2 | 53025757 | A | G | 0.1428 |
| rs2920899 | 2 | 55279681 | T | G | 0.1851 |
| rs72816333 | 2 | 60096560 | A | T | 0.2225 |
| rs2300481 | 2 | 66782467 | T | C | 0.1949 |
| rs13383272 | 2 | 69639911 | A | G | -0.1248 |
| rs72847885 | 2 | 86326717 | A | G | 0.2183 |
| rs6753060 | 2 | 105210762 | T | C | 0.128 |
| rs13006904 | 2 | 107550073 | A | C | 0.1275 |
| rs28377357 | 2 | 112769721 | A | G | -0.1862 |
| rs6723509 | 2 | 122000745 | T | C | 0.2348 |
| rs4456714 | 2 | 124885328 | T | C | -0.139 |
| rs72844590 | 2 | 138421227 | T | G | 0.2083 |
| rs62172389 | 2 | 140378190 | A | C | 0.1417 |
| rs10928206 | 2 | 144965236 | C | G | -0.1309 |
| rs79523138 | 2 | 161368213 | A | G | -0.2629 |
| rs55732192 | 2 | 162278233 | T | G | -0.2399 |
| rs6712203 | 2 | 165557318 | T | C | -0.1561 |
| rs11893300 | 2 | 170526696 | T | G | -0.1274 |
| rs11694601 | 2 | 174949358 | A | G | -0.1545 |
| rs1837164 | 2 | 178716601 | A | T | 0.1497 |
| rs6739913 | 2 | 185033065 | A | G | 0.1675 |
| rs28558491 | 2 | 187816321 | T | C | -0.1874 |
| rs10183297 | 2 | 197342834 | C | G | -0.1894 |
| rs296797 | 2 | 201102905 | T | C | 0.1647 |
| rs1047891 | 2 | 211540507 | A | C | -0.1959 |
| rs12694277 | 2 | 213188795 | T | C | -0.1765 |
| rs1044822 | 2 | 230629138 | T | C | -0.2275 |
| rs3754944 | 2 | 231279616 | A | C | 0.1342 |
| rs13427703 | 2 | 234247508 | C | G | 0.1889 |
| rs2059752 | 2 | 235793752 | T | C | -0.2511 |
| rs139354822 | 2 | 242344695 | T | C | 0.5473 |
| rs6442906 | 3 | 4824012 | T | C | -0.1341 |
| rs3729931 | 3 | 12626516 | A | G | -0.1569 |
| rs189267552 | 3 | 20073193 | A | T | -0.783 |
| rs3911499 | 3 | 21670219 | A | G | -0.1017 |
| rs111464338 | 3 | 26922146 | T | C | -0.6111 |
| rs12638085 | 3 | 30405936 | A | T | 0.2132 |
| rs6788984 | 3 | 41107173 | A | G | 0.2558 |
| rs9311344 | 3 | 43940061 | A | G | 0.1833 |
| rs6774721 | 3 | 49381898 | C | G | 0.2457 |
| rs4499560 | 3 | 70920485 | A | T | -0.2009 |
| rs9857362 | 3 | 74710462 | A | C | 0.151 |
| rs1027829 | 3 | 78661129 | T | G | 0.1275 |
| rs1375564 | 3 | 85656311 | T | C | 0.2343 |
| rs1882289 | 3 | 114461208 | A | G | -0.2159 |
| rs6805393 | 3 | 117492152 | A | G | -0.1134 |
| rs9842387 | 3 | 121616995 | T | C | 0.1379 |
| rs6438857 | 3 | 124557643 | T | C | 0.2142 |
| rs6802742 | 3 | 126282060 | T | C | 0.4007 |
| rs9875380 | 3 | 132780356 | T | C | -0.1651 |
| rs863930 | 3 | 135949737 | A | G | 0.1629 |
| rs893982 | 3 | 148616274 | C | G | -0.1377 |
| rs62271373 | 3 | 150066540 | A | T | 0.4226 |
| rs78151625 | 3 | 158316726 | T | C | -0.2108 |
| rs262986 | 3 | 183435713 | A | G | -0.1856 |
| rs112813049 | 3 | 183974480 | T | C | 0.3183 |
| rs9869437 | 3 | 196228360 | A | C | -0.1522 |
| rs231708 | 4 | 2694773 | C | G | -0.2281 |
| rs7698832 | 4 | 4834691 | A | T | 0.1073 |
| rs2610990 | 4 | 18008232 | A | G | -0.2573 |
| rs12511987 | 4 | 46595623 | T | G | -0.2153 |
| rs10008637 | 4 | 77414144 | T | C | 0.1879 |
| rs13149209 | 4 | 89750668 | T | C | 0.2509 |
| rs1347345 | 4 | 95938386 | A | G | -0.1792 |
| rs3097937 | 4 | 124794644 | A | T | 0.1587 |
| rs7439567 | 4 | 138464842 | T | C | 0.2355 |
| rs72719160 | 4 | 144051276 | A | T | -0.2335 |
| rs6823767 | 4 | 151295085 | T | C | -0.1731 |
| rs7665985 | 4 | 153006312 | T | C | 0.1402 |
| rs62328661 | 4 | 157148635 | A | G | 0.261 |
| rs17035181 | 4 | 157678511 | T | G | 0.2949 |
| rs78231605 | 4 | 160180569 | T | C | -0.3843 |
| rs4957026 | 5 | 361148 | A | G | 0.1446 |
| rs10069690 | 5 | 1279790 | T | C | 0.2735 |
| rs74774746 | 5 | 33411769 | C | G | -0.1705 |
| rs112197024 | 5 | 35269275 | C | G | 0.5596 |
| rs1694068 | 5 | 53283630 | A | T | 0.2339 |
| rs13179413 | 5 | 55868097 | T | C | 0.2045 |
| rs6875372 | 5 | 64079015 | A | T | 0.1344 |
| rs3121685 | 5 | 65662133 | T | C | -0.1475 |
| rs246973 | 5 | 68007803 | T | C | 0.2244 |
| rs6884092 | 5 | 88109590 | A | C | 0.1832 |
| rs709668 | 5 | 96174186 | A | G | -0.2534 |
| rs1871190 | 5 | 97953719 | T | G | 0.1404 |
| rs2112453 | 5 | 112368522 | A | G | -0.1452 |
| rs62373688 | 5 | 127352807 | A | T | 0.2607 |
| rs3923060 | 5 | 138775301 | T | C | 0.1487 |
| rs13166235 | 5 | 139396275 | A | G | 0.2837 |
| rs702395 | 5 | 140086677 | T | C | 0.191 |
| rs1650911 | 5 | 141740620 | C | G | 0.1787 |
| rs17057329 | 5 | 159433524 | A | G | -0.2383 |
| rs6872902 | 5 | 166662779 | A | G | 0.1648 |
| rs12153395 | 5 | 179411477 | A | G | -0.2528 |
| rs2745599 | 6 | 1613686 | A | G | 0.1908 |
| rs11759779 | 6 | 13475669 | T | C | 0.1294 |
| rs179972 | 6 | 16362692 | T | C | 0.1478 |
| rs9368222 | 6 | 20686996 | A | C | 0.2301 |
| rs12661604 | 6 | 23181119 | A | G | 0.1376 |
| rs2395622 | 6 | 35388758 | T | C | -0.1752 |
| rs12055796 | 6 | 46306547 | T | C | 0.113 |
| rs9449350 | 6 | 82281417 | T | C | -0.1348 |
| rs11757455 | 6 | 112522852 | A | G | -0.2745 |
| rs2498586 | 6 | 118026126 | T | C | -0.1916 |
| rs9401090 | 6 | 119113317 | T | C | 0.1637 |
| rs10782230 | 6 | 126228512 | A | G | 0.1995 |
| rs9885632 | 6 | 131311909 | T | C | 0.2102 |
| rs1015112 | 6 | 137726614 | C | G | -0.1402 |
| rs7763294 | 6 | 140383733 | T | G | -0.1804 |
| rs197456 | 6 | 143088071 | A | G | -0.1955 |
| rs7765526 | 6 | 147713764 | A | G | 0.1444 |
| rs6959688 | 7 | 1966831 | A | G | -0.2002 |
| rs3807925 | 7 | 18543250 | A | G | -0.1512 |
| rs12979 | 7 | 24738164 | C | G | 0.2025 |
| rs1011390 | 7 | 26420092 | T | C | 0.1442 |
| rs10274928 | 7 | 28142088 | A | G | 0.1592 |
| rs10233127 | 7 | 30933453 | A | T | 0.3025 |
| rs6593297 | 7 | 56122058 | A | T | 0.1581 |
| rs10950289 | 7 | 71429308 | A | G | 0.1667 |
| rs6963105 | 7 | 75097488 | A | G | -0.1779 |
| rs848445 | 7 | 77572461 | T | C | -0.2004 |
| rs4732535 | 7 | 83669343 | A | G | -0.1755 |
| rs62470665 | 7 | 114622864 | A | G | 0.0936 |
| rs6466878 | 7 | 123272278 | T | C | 0.1507 |
| rs6953231 | 7 | 127681555 | T | C | -0.1531 |
| rs34072724 | 7 | 130432469 | A | G | -0.2181 |
| rs12707230 | 7 | 135104691 | A | C | -0.1207 |
| rs12703989 | 7 | 140238048 | A | G | 0.1521 |
| rs11771693 | 7 | 150050111 | A | G | 0.1673 |
| rs1870735 | 7 | 155744303 | C | G | 0.1701 |
| rs10093740 | 8 | 617341 | A | G | -0.1403 |
| rs4875958 | 8 | 1721090 | A | G | 0.1762 |
| rs61040371 | 8 | 8503700 | T | C | 0.1493 |
| rs62491354 | 8 | 9730663 | A | G | 0.2565 |
| rs1986971 | 8 | 10268736 | A | G | 0.2377 |
| rs118102099 | 8 | 10836373 | A | T | 0.1885 |
| rs74772683 | 8 | 15060597 | T | C | -0.2227 |
| rs71510683 | 8 | 20006331 | A | T | -0.1803 |
| rs2979470 | 8 | 30288272 | T | C | 0.1618 |
| rs1906672 | 8 | 38130025 | A | G | 0.2715 |
| rs4873492 | 8 | 51947549 | T | C | 0.3012 |
| rs6996733 | 8 | 60535824 | T | C | 0.1882 |
| rs2354862 | 8 | 64501744 | A | C | 0.1921 |
| rs13253358 | 8 | 68920135 | T | C | 0.1732 |
| rs1405349 | 8 | 77192089 | T | C | 0.1732 |
| rs72688070 | 8 | 81393697 | T | C | -0.241 |
| rs62526122 | 8 | 92769569 | A | G | 0.1476 |
| rs142449193 | 8 | 102750597 | T | C | -0.3877 |
| rs79127267 | 8 | 106504609 | A | G | -0.3261 |
| rs10955942 | 8 | 120939911 | T | C | 0.1319 |
| rs62523863 | 8 | 126520544 | A | G | 0.2323 |
| rs4598218 | 8 | 129483956 | T | C | 0.1797 |
| rs4129585 | 8 | 143312933 | A | C | 0.1749 |
| rs2978398 | 8 | 146130326 | A | G | -0.1381 |
| rs520015 | 9 | 211762 | C | G | 0.1203 |
| rs60191654 | 9 | 753648 | A | G | -0.1731 |
| rs28558845 | 9 | 4334791 | C | G | -0.218 |
| rs1332813 | 9 | 9350706 | T | C | 0.1965 |
| rs7874646 | 9 | 13899209 | T | C | -0.1712 |
| rs10757204 | 9 | 21233057 | A | G | -0.1697 |
| rs9886665 | 9 | 22942770 | T | C | 0.1509 |
| rs41296342 | 9 | 71627364 | T | G | -0.1773 |
| rs10746963 | 9 | 77238558 | A | G | -0.1727 |
| rs13290449 | 9 | 81747339 | T | G | -0.1397 |
| rs10820855 | 9 | 94201341 | T | C | -0.1492 |
| rs7045409 | 9 | 95201540 | A | T | -0.1823 |
| rs10820747 | 9 | 107686823 | A | G | 0.1247 |
| rs818692 | 9 | 116165978 | A | G | -0.1429 |
| rs7023828 | 9 | 128498594 | T | C | -0.2436 |
| rs1891730 | 9 | 130309028 | T | C | -0.1785 |
| rs184457 | 9 | 131940019 | A | G | -0.1659 |
| rs3861882 | 9 | 132465304 | T | C | 0.1347 |
| rs11252324 | 10 | 4124568 | T | G | -0.3023 |
| rs56352451 | 10 | 5804865 | T | C | 0.1992 |
| rs11257593 | 10 | 12241815 | T | C | 0.1359 |
| rs7100317 | 10 | 21764469 | A | C | 0.1299 |
| rs3858217 | 10 | 24690604 | C | G | 0.1668 |
| rs3802517 | 10 | 28233469 | A | T | 0.1895 |
| rs11011062 | 10 | 37484723 | A | C | -0.2271 |
| rs34130368 | 10 | 48411796 | T | G | -0.2715 |
| rs72799495 | 10 | 53654149 | A | G | -0.184 |
| rs10762120 | 10 | 68668627 | T | C | -0.1643 |
| rs12572586 | 10 | 74751579 | T | C | -0.3646 |
| rs77413490 | 10 | 89681688 | T | G | 0.473 |
| rs11187142 | 10 | 94468685 | T | C | 0.2378 |
| rs56085433 | 10 | 97326325 | A | G | -0.2098 |
| rs57807455 | 10 | 98558501 | A | T | -0.1664 |
| rs11197208 | 10 | 117187965 | A | T | 0.1232 |
| rs11197813 | 10 | 118523933 | A | G | -0.1801 |
| rs11592107 | 10 | 122968964 | A | G | 0.2306 |
| rs72834453 | 10 | 124235226 | T | G | -0.276 |
| rs7912283 | 10 | 133773019 | A | G | 0.1943 |
| rs1133400 | 10 | 134459388 | A | G | -0.2607 |
| rs10743086 | 11 | 8774923 | A | G | -0.1832 |
| rs10766533 | 11 | 19224677 | A | T | 0.1665 |
| rs871004 | 11 | 28512458 | A | G | 0.1999 |
| rs11031051 | 11 | 30355707 | A | C | -0.1899 |
| rs190194639 | 11 | 34068037 | T | C | 0.2746 |
| rs1585453 | 11 | 46884713 | A | T | -0.3892 |
| rs4385883 | 11 | 51539339 | T | G | -0.2331 |
| rs75905900 | 11 | 55113534 | A | C | 0.3883 |
| rs4980515 | 11 | 63744609 | T | C | 0.2086 |
| rs574381 | 11 | 64757504 | C | G | -0.1169 |
| rs67976715 | 11 | 68023742 | C | G | 0.1971 |
| rs7110305 | 11 | 76716483 | T | G | -0.1263 |
| rs11600236 | 11 | 91864848 | T | C | 0.1112 |
| rs67885470 | 11 | 99998431 | T | C | -0.1672 |
| rs4754196 | 11 | 107096777 | A | G | -0.2884 |
| rs7128707 | 11 | 112855354 | A | T | -0.1648 |
| rs1076485 | 11 | 116772441 | T | C | 0.3126 |
| rs2155572 | 11 | 123067655 | T | G | 0.1341 |
| rs78998485 | 12 | 434755 | C | G | -0.2036 |
| rs11571376 | 12 | 1059556 | C | G | -0.1884 |
| rs2024385 | 12 | 12888438 | A | T | -0.2326 |
| rs28621435 | 12 | 13860990 | A | G | -0.1932 |
| rs7976167 | 12 | 24210599 | T | C | 0.1638 |
| rs10437954 | 12 | 58003922 | A | G | -0.3471 |
| rs4143175 | 12 | 67782397 | T | C | 0.1802 |
| rs2083857 | 12 | 73931720 | T | C | -0.1431 |
| rs7963801 | 12 | 79685226 | T | C | -0.2027 |
| rs10858966 | 12 | 90567026 | C | G | 0.254 |
| rs5742643 | 12 | 102837863 | A | G | -0.2236 |
| rs11112548 | 12 | 105871914 | A | T | 0.4608 |
| rs278121 | 12 | 120152743 | T | C | 0.1264 |
| rs117206641 | 12 | 133086888 | A | G | 0.2157 |
| rs2480171 | 13 | 21559858 | T | C | 0.2047 |
| rs606950 | 13 | 22298923 | A | G | 0.2324 |
| rs1331012 | 13 | 27115424 | T | G | 0.1632 |
| rs9507885 | 13 | 27951090 | T | C | -0.2347 |
| rs7319898 | 13 | 28633433 | A | G | 0.1441 |
| rs9532243 | 13 | 32191408 | A | C | 0.2709 |
| rs1998727 | 13 | 33713419 | A | G | 0.1487 |
| rs2152258 | 13 | 40668694 | T | C | 0.1444 |
| rs4274337 | 13 | 41967193 | A | G | -0.22 |
| rs73187288 | 13 | 42738672 | A | C | -0.2558 |
| rs912434 | 13 | 47189928 | T | G | 0.2151 |
| rs9526707 | 13 | 51489186 | A | G | -0.1939 |
| rs9316725 | 13 | 55077856 | T | G | -0.136 |
| rs75961402 | 13 | 56398286 | A | G | 0.182 |
| rs17245822 | 13 | 73131694 | A | C | -0.1516 |
| rs78474310 | 13 | 73826901 | A | G | -0.4828 |
| rs7988232 | 13 | 79808655 | A | G | 0.148 |
| rs7491529 | 13 | 97020634 | C | G | 0.1208 |
| rs7331680 | 13 | 115000650 | T | G | 0.3864 |
| rs17115145 | 14 | 30122409 | T | C | 0.1569 |
| rs34983854 | 14 | 39858442 | A | G | -0.1624 |
| rs72683923 | 14 | 50735947 | T | C | 0.9769 |
| rs72682483 | 14 | 52798795 | A | G | 0.3238 |
| rs8021944 | 14 | 64679298 | T | G | -0.1564 |
| rs11623535 | 14 | 72462381 | A | G | 0.2022 |
| rs11159091 | 14 | 75074316 | A | G | 0.1793 |
| rs8007818 | 14 | 91584108 | A | G | -0.1511 |
| rs8014182 | 14 | 103859962 | T | C | -0.3397 |
| rs12439246 | 15 | 37279531 | C | G | -0.1273 |
| rs11629850 | 15 | 40317075 | A | G | 0.1995 |
| rs12902197 | 15 | 51431658 | A | G | 0.1336 |
| rs17730281 | 15 | 53907948 | A | G | -0.1366 |
| rs4775376 | 15 | 61517218 | T | C | 0.138 |
| rs11634028 | 15 | 76276150 | A | T | 0.2258 |
| rs3743157 | 15 | 85680532 | A | C | 0.2602 |
| rs11632436 | 15 | 86295286 | C | G | 0.2056 |
| rs12595031 | 15 | 96024055 | T | C | 0.138 |
| rs12901664 | 15 | 98338524 | T | C | 0.1515 |
| rs4965529 | 15 | 100145224 | T | C | -0.2497 |
| rs2379829 | 16 | 3538873 | C | G | -0.2024 |
| rs35450617 | 16 | 6889675 | T | G | -0.1609 |
| rs7198817 | 16 | 11945778 | A | C | -0.15 |
| rs34941092 | 16 | 50550137 | A | G | -0.2303 |
| rs12922061 | 16 | 52635000 | T | C | 0.1488 |
| rs2060664 | 16 | 60652439 | T | C | 0.1688 |
| rs75200440 | 16 | 67425646 | T | C | -0.2224 |
| rs1012089 | 16 | 74171973 | C | G | -0.1312 |
| rs62048897 | 16 | 76997174 | T | C | 0.1308 |
| rs7187540 | 16 | 85318302 | A | C | -0.1766 |
| rs3851018 | 16 | 86437811 | C | G | -0.1175 |
| rs6540125 | 16 | 87993889 | T | G | 0.164 |
| rs71355297 | 17 | 568515 | T | C | 0.1351 |
| rs4480845 | 17 | 1958609 | T | C | 0.2595 |
| rs113821951 | 17 | 2550415 | A | G | 0.1968 |
| rs4925159 | 17 | 18185510 | A | G | 0.2281 |
| rs7218708 | 17 | 19926836 | A | G | -0.1436 |
| rs704 | 17 | 26694861 | A | G | -0.1245 |
| rs1551355 | 17 | 30032420 | T | C | 0.1932 |
| rs9899540 | 17 | 30777924 | A | G | -0.1458 |
| rs6503732 | 17 | 36879668 | T | C | -0.1385 |
| rs34430710 | 17 | 56876627 | A | T | -0.2069 |
| rs1036902 | 17 | 58950791 | T | C | -0.2273 |
| rs6504213 | 17 | 62381714 | T | C | -0.2441 |
| rs112260610 | 17 | 64252393 | T | C | 0.2216 |
| rs62086903 | 17 | 66016006 | T | C | 0.1454 |
| rs9302885 | 17 | 76799898 | A | G | 0.1945 |
| rs72847097 | 17 | 78248369 | T | C | -0.0997 |
| rs112280096 | 17 | 79367409 | A | C | -0.1706 |
| rs34413141 | 18 | 777282 | A | T | -0.3084 |
| rs11664106 | 18 | 2846812 | A | T | 0.1339 |
| rs62082230 | 18 | 22676071 | A | T | -0.1646 |
| rs1154214 | 18 | 24546824 | T | G | -0.1476 |
| rs12963523 | 18 | 26687028 | A | G | -0.1215 |
| rs11876341 | 18 | 48799991 | A | G | -0.2225 |
| rs10048404 | 18 | 54578482 | T | C | -0.2277 |
| rs6567160 | 18 | 57829135 | T | C | 0.1542 |
| rs12454712 | 18 | 60845884 | T | C | 0.2038 |
| rs10460108 | 18 | 73034151 | A | G | 0.2118 |
| rs2613765 | 19 | 5066330 | A | G | -0.2336 |
| rs10409243 | 19 | 10332988 | T | C | -0.2415 |
| rs1013424 | 19 | 29693318 | A | G | -0.0921 |
| rs7256564 | 19 | 33889593 | A | G | 0.1533 |
| rs256741 | 19 | 37855839 | A | C | 0.0847 |
| rs3786872 | 19 | 38756089 | C | G | 0.1501 |
| rs3810299 | 19 | 46800228 | T | C | -0.2019 |
| rs1236093 | 19 | 49104308 | T | C | 0.143 |
| rs73046792 | 19 | 49605705 | A | G | -0.282 |
| rs138877676 | 19 | 50935809 | T | G | -0.6556 |
| rs329716 | 19 | 53590507 | A | G | -0.1344 |
| rs2024866 | 20 | 1862223 | A | G | -0.3689 |
| rs1764975 | 20 | 4101290 | T | C | -0.2162 |
| rs6137201 | 20 | 20957669 | A | T | 0.1643 |
| rs6021247 | 20 | 50108980 | A | G | 0.2041 |
| rs2801008 | 20 | 51788718 | T | G | -0.163 |
| rs749209 | 20 | 57088189 | T | C | -0.1292 |
| rs28374392 | 20 | 61189717 | T | C | 0.129 |
| rs2297387 | 20 | 62078360 | A | G | -0.1208 |
| rs1882961 | 21 | 16556367 | T | C | 0.2346 |
| rs9975838 | 21 | 34299793 | A | T | 0.1799 |
| rs9608690 | 22 | 28921347 | A | G | -0.2983 |
| rs143040273 | 22 | 31273453 | T | C | 0.4194 |
| rs28578714 | 22 | 50727921 | T | C | 0.1951 |

**Supplementary Table 19.** Genetic variants and weights used to construct diastolic blood pressure PRS.

| SNP | Chromosome | Chromosome position | Effect allele | Non-effect allele | Beta |
| --- | --- | --- | --- | --- | --- |
| rs4908678 | 1 | 7739250 | C | T | 0.0924 |
| rs2252865 | 1 | 8422676 | T | C | 0.1137 |
| rs3013095 | 1 | 13798362 | T | G | 0.0847 |
| rs55755393 | 1 | 26780724 | A | G | 0.0857 |
| rs1565716 | 1 | 29549216 | A | G | 0.2012 |
| rs1474181 | 1 | 32136610 | G | C | 0.0566 |
| rs17402858 | 1 | 44422622 | C | T | 0.1918 |
| rs56810307 | 1 | 44973546 | A | T | 0.1102 |
| rs12142296 | 1 | 46541679 | G | T | 0.1716 |
| rs4926923 | 1 | 48109225 | T | C | 0.1613 |
| rs241468 | 1 | 49800692 | A | C | 0.1057 |
| rs6681713 | 1 | 51527684 | T | G | 0.4074 |
| rs835352 | 1 | 53048482 | C | T | 0.0422 |
| rs10889711 | 1 | 68143195 | T | C | 0.0892 |
| rs34517439 | 1 | 78450517 | C | A | 0.233 |
| rs2275902 | 1 | 79357360 | G | C | 0.0874 |
| rs2065152 | 1 | 90228519 | T | C | 0.0877 |
| rs11164868 | 1 | 93524944 | C | T | 0.1784 |
| rs892511 | 1 | 110078255 | C | G | 0.2192 |
| rs11466111 | 1 | 115829178 | T | C | 0.5727 |
| rs12078697 | 1 | 117015118 | G | C | 0.1083 |
| rs1886914 | 1 | 119541452 | C | T | 0.1113 |
| rs72704264 | 1 | 145713305 | C | G | 0.1173 |
| rs13796 | 1 | 154245917 | C | T | 0.1376 |
| rs11586348 | 1 | 154819359 | A | C | 0.0779 |
| rs56002643 | 1 | 163181430 | G | A | 0.0836 |
| rs2171690 | 1 | 164740099 | T | C | 0.1048 |
| rs7524019 | 1 | 167367193 | T | C | 0.0989 |
| rs3766694 | 1 | 175143207 | T | C | 0.0931 |
| rs150816167 | 1 | 179571862 | C | T | 0.2641 |
| rs41475048 | 1 | 183058452 | G | A | 0.1024 |
| rs882624 | 1 | 201735913 | C | T | 0.1536 |
| rs12403358 | 1 | 203995994 | C | G | 0.0863 |
| rs2629665 | 1 | 207220800 | C | A | 0.1019 |
| rs35981664 | 1 | 218549354 | T | A | 0.1542 |
| rs9431431 | 1 | 221358796 | G | A | 0.1131 |
| rs10127776 | 1 | 226474479 | G | A | 0.0899 |
| rs1885018 | 1 | 236316575 | A | C | 0.1223 |
| rs56236159 | 2 | 3636478 | C | T | 0.1285 |
| rs112393817 | 2 | 9807226 | C | G | 0.1068 |
| rs139122584 | 2 | 24616446 | T | C | 0.2841 |
| rs1033751 | 2 | 32695759 | T | C | 0.0674 |
| rs1607644 | 2 | 34679626 | G | A | 0.1035 |
| rs13392090 | 2 | 36580097 | C | A | 0.0879 |
| rs2707238 | 2 | 38094149 | C | G | 0.1019 |
| rs11681462 | 2 | 42352567 | C | A | 0.0983 |
| rs6749716 | 2 | 47089862 | G | A | 0.077 |
| rs62140266 | 2 | 54675716 | A | G | 0.0795 |
| rs7608483 | 2 | 61836235 | A | C | 0.1027 |
| rs13014371 | 2 | 64217786 | C | T | 0.1036 |
| rs12052761 | 2 | 69065841 | G | A | 0.0985 |
| rs10193543 | 2 | 72483329 | T | G | 0.1222 |
| rs6730785 | 2 | 73642403 | A | C | 0.0947 |
| rs115528323 | 2 | 95972159 | T | C | 0.2166 |
| rs4851462 | 2 | 98357163 | C | T | 0.1138 |
| rs12622114 | 2 | 106906460 | G | C | 0.1272 |
| rs72948328 | 2 | 111881810 | T | C | 0.1811 |
| rs62158170 | 2 | 114082175 | A | G | 0.1428 |
| rs10864859 | 2 | 121440218 | T | G | 0.1714 |
| rs4849000 | 2 | 126029884 | C | T | 0.0776 |
| rs13001283 | 2 | 127183454 | A | G | 0.1513 |
| rs4954192 | 2 | 135632981 | C | T | 0.1211 |
| rs7606205 | 2 | 144146311 | C | A | 0.1207 |
| rs34570306 | 2 | 146272860 | C | T | 0.1115 |
| rs62169544 | 2 | 146950908 | G | A | 0.0947 |
| rs12990959 | 2 | 148572160 | C | T | 0.1071 |
| rs1220128 | 2 | 158499902 | C | G | 0.1733 |
| rs72936986 | 2 | 159429379 | A | C | 0.0954 |
| rs2390258 | 2 | 166250129 | G | A | 0.1031 |
| rs6758859 | 2 | 173965056 | T | C | 0.1053 |
| rs10184839 | 2 | 181946115 | T | A | 0.0972 |
| rs2053163 | 2 | 190827023 | A | G | 0.0836 |
| rs72926986 | 2 | 203519264 | G | T | 0.0981 |
| rs2162003 | 2 | 205077128 | T | C | 0.1085 |
| rs1263671 | 2 | 207996447 | C | T | 0.1194 |
| rs112905092 | 2 | 214012225 | T | C | 0.3771 |
| rs1996992 | 2 | 219651349 | G | T | 0.2964 |
| rs12474050 | 2 | 220362557 | T | C | 0.0963 |
| rs4507125 | 2 | 239864732 | C | A | 0.1081 |
| rs9865843 | 3 | 7489993 | G | A | 0.0906 |
| rs6442608 | 3 | 16588273 | C | T | 0.1107 |
| rs4634143 | 3 | 23163749 | T | C | 0.1129 |
| rs7628351 | 3 | 25422475 | C | G | 0.1038 |
| rs72851229 | 3 | 29374219 | G | C | 0.1202 |
| rs113134141 | 3 | 46861939 | G | A | 0.1569 |
| rs11721328 | 3 | 47397300 | A | G | 0.1389 |
| rs144483183 | 3 | 48626745 | T | C | 0.2093 |
| rs7631822 | 3 | 55612678 | T | A | 0.0723 |
| rs11130567 | 3 | 57074119 | T | C | 0.0932 |
| rs12486605 | 3 | 57706503 | C | T | 0.1437 |
| rs3774702 | 3 | 63856870 | A | G | 0.1244 |
| rs3911952 | 3 | 73797420 | G | T | 0.0713 |
| rs11923667 | 3 | 101268080 | A | T | 0.1222 |
| rs28675079 | 3 | 111500002 | G | A | 0.1313 |
| rs9834975 | 3 | 122117663 | T | A | 0.0836 |
| rs62264113 | 3 | 127292333 | A | G | 0.1332 |
| rs2369796 | 3 | 132196816 | A | T | 0.0875 |
| rs73158427 | 3 | 153721493 | A | T | 0.1436 |
| rs7611674 | 3 | 179169230 | T | G | 0.1276 |
| rs147501096 | 3 | 186180253 | G | C | 0.1602 |
| rs6776964 | 3 | 189304602 | A | G | 0.0914 |
| rs1706003 | 3 | 194299967 | T | G | 0.1326 |
| rs6777317 | 3 | 197070959 | A | G | 0.1147 |
| rs55829085 | 4 | 2165493 | C | A | 0.2672 |
| rs28667801 | 4 | 26785356 | T | A | 0.1449 |
| rs1878825 | 4 | 36091370 | G | C | 0.1008 |
| rs11945489 | 4 | 56463775 | C | T | 0.1052 |
| rs1718845 | 4 | 57943153 | G | A | 0.0963 |
| rs6551716 | 4 | 63575696 | A | T | 0.1115 |
| rs6829135 | 4 | 71957526 | G | C | 0.0948 |
| rs34587285 | 4 | 73349491 | T | C | 0.0905 |
| rs17484253 | 4 | 81696280 | A | G | 0.3186 |
| rs28682481 | 4 | 87779081 | A | T | 0.0974 |
| rs7694000 | 4 | 95324968 | T | A | 0.0954 |
| rs17248480 | 4 | 102435265 | G | A | 0.5064 |
| rs223361 | 4 | 103769304 | T | C | 0.1522 |
| rs7694643 | 4 | 109017528 | G | A | 0.1209 |
| rs17595265 | 4 | 119455055 | A | T | 0.0937 |
| rs4834735 | 4 | 119958809 | T | C | 0.1265 |
| rs4696248 | 4 | 152254350 | A | C | 0.0841 |
| rs4342155 | 4 | 163273274 | C | T | 0.1248 |
| rs954767 | 5 | 3706050 | C | A | 0.1389 |
| rs42551 | 5 | 14321785 | A | T | 0.1058 |
| rs2921604 | 5 | 14867948 | C | T | 0.0829 |
| rs72732580 | 5 | 36785541 | A | G | 0.0769 |
| rs172906 | 5 | 38616887 | C | A | 0.092 |
| rs863241 | 5 | 40805847 | T | G | 0.0717 |
| rs2940928 | 5 | 42423905 | G | A | 0.1131 |
| rs6875967 | 5 | 50878292 | A | G | 0.1108 |
| rs17735275 | 5 | 56558949 | A | G | 0.2923 |
| rs12515541 | 5 | 57095011 | T | G | 0.1104 |
| rs1848510 | 5 | 57754005 | A | G | 0.1061 |
| rs10062049 | 5 | 61553881 | T | C | 0.2113 |
| rs17396761 | 5 | 62204890 | A | G | 0.1117 |
| rs4286632 | 5 | 66291370 | A | G | 0.127 |
| rs62362531 | 5 | 76722442 | C | T | 0.0996 |
| rs62380354 | 5 | 89484911 | A | C | 0.15 |
| rs7731530 | 5 | 91983354 | A | C | 0.1126 |
| rs7714279 | 5 | 95527383 | T | C | 0.0718 |
| rs529279 | 5 | 111098877 | C | T | 0.0835 |
| rs114707644 | 5 | 113030009 | A | G | 0.2176 |
| rs6892204 | 5 | 118605139 | A | G | 0.0931 |
| rs55747751 | 5 | 132397351 | G | A | 0.2159 |
| rs3776299 | 5 | 142507651 | A | G | 0.1104 |
| rs596206 | 5 | 153890770 | C | T | 0.0907 |
| rs77625383 | 5 | 157231462 | C | T | 0.1307 |
| rs6555948 | 5 | 170804832 | A | G | 0.0828 |
| rs114503346 | 5 | 172192350 | C | T | 0.2415 |
| rs28362590 | 5 | 176731452 | T | G | 0.096 |
| rs6873880 | 5 | 180078903 | T | G | 0.0867 |
| rs142843359 | 6 | 11612120 | G | A | 0.2769 |
| rs2282823 | 6 | 15478336 | T | C | 0.1131 |
| rs61408836 | 6 | 17302857 | C | T | 0.0997 |
| rs2328238 | 6 | 18327303 | C | T | 0.0935 |
| rs115245297 | 6 | 34244132 | C | T | 0.3148 |
| rs4714224 | 6 | 39186743 | G | C | 0.139 |
| rs9472135 | 6 | 43809802 | T | C | 0.1444 |
| rs1359785 | 6 | 51190813 | G | A | 0.0812 |
| rs504691 | 6 | 72206620 | C | A | 0.1017 |
| rs7753695 | 6 | 80818531 | T | C | 0.0903 |
| rs1933718 | 6 | 98341457 | C | T | 0.0952 |
| rs9496614 | 6 | 100613551 | T | C | 0.12 |
| rs72613227 | 6 | 106320771 | T | A | 0.1608 |
| rs9386780 | 6 | 109584410 | A | G | 0.0823 |
| rs9320368 | 6 | 111904737 | C | T | 0.0759 |
| rs3822857 | 6 | 116313931 | G | C | 0.1194 |
| rs2693560 | 6 | 117523671 | G | A | 0.1475 |
| rs995207 | 6 | 136034677 | G | A | 0.0873 |
| rs668459 | 6 | 139835689 | C | T | 0.1219 |
| rs645144 | 6 | 141180780 | T | C | 0.0919 |
| rs76194939 | 6 | 144303140 | C | T | 0.1173 |
| rs9479509 | 6 | 153427265 | G | A | 0.0992 |
| rs598682 | 6 | 154418759 | G | A | 0.1153 |
| rs73013270 | 6 | 157075800 | T | G | 0.1338 |
| rs9456648 | 6 | 161712235 | C | T | 0.0985 |
| rs12665161 | 6 | 163749986 | A | G | 0.1074 |
| rs2144249 | 6 | 170694084 | T | C | 0.1166 |
| rs73033340 | 7 | 1195692 | A | G | 0.4615 |
| rs73049928 | 7 | 4669949 | G | A | 0.1167 |
| rs1468520 | 7 | 7290732 | G | A | 0.14 |
| rs13240040 | 7 | 14375977 | A | G | 0.0754 |
| rs818508 | 7 | 16719917 | C | T | 0.1005 |
| rs10263380 | 7 | 21578343 | C | A | 0.0703 |
| rs4507656 | 7 | 22156538 | G | C | 0.1275 |
| rs7781425 | 7 | 34944387 | G | A | 0.0917 |
| rs342989 | 7 | 35467896 | A | G | 0.1407 |
| rs73105827 | 7 | 45036785 | G | T | 0.1699 |
| rs17454517 | 7 | 50915776 | A | G | 0.1097 |
| rs2222544 | 7 | 69769369 | C | T | 0.0987 |
| rs1178979 | 7 | 72856430 | T | C | 0.1362 |
| rs34324971 | 7 | 74107374 | A | G | 0.1199 |
| rs3807101 | 7 | 80393418 | C | T | 0.1536 |
| rs1947228 | 7 | 96461649 | C | T | 0.1451 |
| rs7788746 | 7 | 99612405 | G | T | 0.1463 |
| rs12536606 | 7 | 107907875 | C | T | 0.0942 |
| rs144867634 | 7 | 111580166 | T | C | 0.249 |
| rs1722886 | 7 | 134215259 | A | T | 0.1166 |
| rs9638084 | 7 | 156311745 | A | G | 0.1107 |
| rs11136373 | 8 | 1212030 | G | C | 0.0757 |
| rs2922895 | 8 | 6379932 | C | G | 0.1128 |
| rs75902664 | 8 | 17427186 | G | A | 0.3174 |
| rs1047030 | 8 | 22428708 | A | G | 0.1291 |
| rs62503324 | 8 | 23400615 | T | C | 0.1931 |
| rs17321041 | 8 | 26445194 | T | C | 0.1599 |
| rs2955067 | 8 | 48674068 | G | A | 0.1411 |
| rs72639047 | 8 | 49415598 | G | A | 0.1279 |
| rs1440786 | 8 | 53386563 | C | T | 0.0759 |
| rs7843475 | 8 | 72475748 | G | C | 0.1073 |
| rs56345595 | 8 | 82814156 | A | G | 0.1157 |
| rs7815598 | 8 | 88316794 | A | G | 0.0656 |
| rs2142141 | 8 | 90940205 | G | C | 0.0889 |
| rs2250070 | 8 | 97519421 | C | A | 0.0696 |
| rs2513877 | 8 | 103883630 | G | A | 0.1015 |
| rs13270541 | 8 | 124794849 | A | G | 0.11 |
| rs2979040 | 8 | 134125029 | A | G | 0.0873 |
| rs10087782 | 8 | 141858620 | T | C | 0.1211 |
| rs56233017 | 8 | 144981488 | G | A | 0.2669 |
| rs12216886 | 9 | 2493751 | T | G | 0.1175 |
| rs35287509 | 9 | 10594635 | C | T | 0.0919 |
| rs13293465 | 9 | 37127678 | C | T | 0.0817 |
| rs11141731 | 9 | 89888472 | C | T | 0.1043 |
| rs9410352 | 9 | 91406863 | C | A | 0.0812 |
| rs35371869 | 9 | 92458311 | G | A | 0.0786 |
| rs1317939 | 9 | 95964564 | A | G | 0.0758 |
| rs7020564 | 9 | 109670016 | T | A | 0.1105 |
| rs7043304 | 9 | 112358150 | T | C | 0.1458 |
| rs1861881 | 9 | 119312256 | T | G | 0.0968 |
| rs4837127 | 9 | 129792141 | T | C | 0.1058 |
| rs4842069 | 9 | 138891645 | G | C | 0.0821 |
| rs11145807 | 9 | 139520789 | A | G | 0.1188 |
| rs10906391 | 10 | 13523937 | T | C | 0.121 |
| rs7095584 | 10 | 14024991 | A | G | 0.0918 |
| rs6602177 | 10 | 17167141 | C | T | 0.1156 |
| rs1265842 | 10 | 28924901 | T | C | 0.0998 |
| rs76164690 | 10 | 32590362 | G | T | 0.1373 |
| rs1487562 | 10 | 45928822 | T | C | 0.0942 |
| rs7904833 | 10 | 52279428 | A | C | 0.0687 |
| rs10761530 | 10 | 62390726 | T | C | 0.0955 |
| rs7090758 | 10 | 65335315 | C | T | 0.135 |
| rs17476364 | 10 | 71094504 | T | C | 0.1255 |
| rs303217 | 10 | 91153699 | C | T | 0.0818 |
| rs603424 | 10 | 102075479 | A | G | 0.1717 |
| rs72847884 | 10 | 103115345 | A | G | 0.219 |
| rs191784289 | 10 | 106894942 | T | C | 0.6852 |
| rs111777102 | 10 | 111965826 | T | C | 0.1843 |
| rs117707765 | 10 | 112608355 | C | T | 0.095 |
| rs7082836 | 10 | 116646237 | A | G | 0.0797 |
| rs72842207 | 10 | 121433675 | C | T | 0.2112 |
| rs4411245 | 10 | 126712781 | A | G | 0.1023 |
| rs7081529 | 10 | 129772048 | C | G | 0.084 |
| rs17224476 | 11 | 4673788 | A | G | 0.1547 |
| rs2929184 | 11 | 6289118 | A | G | 0.1164 |
| rs360153 | 11 | 9762274 | C | T | 0.1917 |
| rs7479680 | 11 | 14154711 | C | A | 0.1591 |
| rs10832300 | 11 | 14788963 | G | A | 0.1016 |
| rs11026586 | 11 | 22515533 | A | G | 0.2606 |
| rs919045 | 11 | 31111810 | T | C | 0.1017 |
| rs61879810 | 11 | 31821467 | A | G | 0.1463 |
| rs875106 | 11 | 70005641 | G | A | 0.1217 |
| rs504217 | 11 | 72006086 | T | C | 0.2069 |
| rs2298807 | 11 | 73068571 | T | C | 0.0945 |
| rs4420291 | 11 | 74374950 | A | G | 0.0937 |
| rs59986178 | 11 | 77359909 | C | G | 0.1572 |
| rs2450128 | 11 | 77940075 | G | A | 0.1225 |
| rs56263826 | 11 | 84308799 | G | A | 0.1175 |
| rs11021221 | 11 | 95308854 | T | A | 0.1772 |
| rs61909958 | 11 | 96151677 | C | G | 0.1127 |
| rs61892344 | 11 | 101100768 | C | T | 0.1162 |
| rs4250067 | 11 | 108295315 | T | C | 0.0784 |
| rs12362593 | 11 | 111586091 | G | C | 0.1106 |
| rs12790943 | 11 | 120058623 | C | T | 0.0873 |
| rs12574332 | 11 | 122521123 | T | C | 0.1747 |
| rs11604175 | 11 | 124619407 | T | C | 0.0984 |
| rs66518071 | 11 | 128762184 | C | G | 0.1163 |
| rs55935819 | 12 | 2521579 | A | G | 0.1088 |
| rs75507123 | 12 | 5417856 | G | T | 0.1396 |
| rs7132012 | 12 | 8832203 | A | G | 0.148 |
| rs7313556 | 12 | 15297359 | A | G | 0.09 |
| rs7136879 | 12 | 17884529 | T | G | 0.0647 |
| rs61912333 | 12 | 19554817 | C | G | 0.1051 |
| rs1098708 | 12 | 27321112 | G | A | 0.0931 |
| rs7965392 | 12 | 42540280 | A | G | 0.0972 |
| rs11168245 | 12 | 48204499 | C | G | 0.1822 |
| rs61926181 | 12 | 50767037 | G | A | 0.4032 |
| rs3782480 | 12 | 52305069 | T | G | 0.1413 |
| rs7137749 | 12 | 57098040 | T | C | 0.132 |
| rs6581833 | 12 | 69169222 | T | A | 0.0823 |
| rs521033 | 12 | 69951428 | T | C | 0.1605 |
| rs11178076 | 12 | 70477510 | C | T | 0.1186 |
| rs61924955 | 12 | 89507623 | C | T | 0.1309 |
| rs11108209 | 12 | 96109855 | C | T | 0.1539 |
| rs7134060 | 12 | 96717095 | G | A | 0.0999 |
| rs34318061 | 12 | 104891832 | A | G | 0.088 |
| rs141391620 | 12 | 110681099 | A | G | 0.2237 |
| rs12184466 | 12 | 111281636 | T | C | 0.2207 |
| rs3898618 | 12 | 120813921 | C | T | 0.2532 |
| rs3213545 | 12 | 121471337 | A | G | 0.097 |
| rs181522314 | 12 | 122060212 | C | T | 0.3271 |
| rs28498002 | 12 | 122599796 | C | T | 0.1359 |
| rs1271309 | 12 | 124820705 | G | A | 0.1786 |
| rs12828318 | 12 | 133766122 | A | G | 0.1048 |
| rs55641580 | 13 | 25257917 | T | C | 0.1544 |
| rs56312513 | 13 | 38249726 | G | A | 0.0979 |
| rs9549297 | 13 | 41397482 | G | A | 0.1472 |
| rs12583637 | 13 | 50564490 | A | G | 0.1671 |
| rs9563529 | 13 | 58316637 | T | G | 0.1173 |
| rs9597982 | 13 | 59911420 | C | G | 0.0837 |
| rs3861113 | 13 | 72364382 | A | C | 0.1806 |
| rs1215469 | 13 | 80707408 | C | A | 0.1219 |
| rs55684003 | 13 | 97988689 | A | G | 0.1193 |
| rs650724 | 13 | 110804809 | G | A | 0.182 |
| rs36152087 | 13 | 114454891 | T | G | 0.1122 |
| rs7350752 | 14 | 21841154 | G | A | 0.1438 |
| rs17880989 | 14 | 23313633 | A | G | 0.3648 |
| rs1950500 | 14 | 24830850 | T | C | 0.1212 |
| rs2273171 | 14 | 31381351 | C | T | 0.0868 |
| rs4424827 | 14 | 35110857 | C | T | 0.0948 |
| rs35329957 | 14 | 41564491 | C | G | 0.0701 |
| rs11628933 | 14 | 60700903 | G | C | 0.1338 |
| rs731681 | 14 | 68010224 | G | C | 0.0943 |
| rs57786342 | 14 | 69260028 | A | G | 0.1141 |
| rs227426 | 14 | 70456664 | T | G | 0.0893 |
| rs4903064 | 14 | 73279420 | T | C | 0.1448 |
| rs1811739 | 14 | 77529375 | A | G | 0.1001 |
| rs117754243 | 14 | 84867375 | G | T | 0.1653 |
| rs34161718 | 14 | 104620193 | C | T | 0.1192 |
| rs10873612 | 15 | 26105602 | C | T | 0.0828 |
| rs2925345 | 15 | 41311799 | T | C | 0.1602 |
| rs4924570 | 15 | 41974660 | C | T | 0.1548 |
| rs2447195 | 15 | 43931449 | G | A | 0.1272 |
| rs11857515 | 15 | 49606320 | A | T | 0.1222 |
| rs62004794 | 15 | 68454523 | G | A | 0.0948 |
| rs11853359 | 15 | 71621524 | G | A | 0.1608 |
| rs2957743 | 15 | 72302030 | C | T | 0.0969 |
| rs61653296 | 15 | 74557817 | G | A | 0.1123 |
| rs983353 | 15 | 82186535 | G | A | 0.095 |
| rs7180952 | 15 | 85162551 | C | T | 0.0984 |
| rs78550103 | 15 | 90027296 | G | A | 0.1451 |
| rs873122 | 15 | 92702020 | C | G | 0.1147 |
| rs28533980 | 15 | 94207872 | T | C | 0.0883 |
| rs4984496 | 15 | 96635898 | T | G | 0.1584 |
| rs9932866 | 16 | 706067 | A | G | 0.1098 |
| rs28590346 | 16 | 2080653 | T | A | 0.1786 |
| rs9937170 | 16 | 10624905 | T | C | 0.0741 |
| rs57327054 | 16 | 14487036 | C | T | 0.0944 |
| rs59333122 | 16 | 19149996 | A | G | 0.1069 |
| rs9937801 | 16 | 21088130 | T | C | 0.1301 |
| rs61739285 | 16 | 27480797 | T | C | 0.2187 |
| rs6565174 | 16 | 30111904 | C | A | 0.1821 |
| rs10468291 | 16 | 49768046 | C | A | 0.0981 |
| rs9932220 | 16 | 51758116 | G | A | 0.1347 |
| rs12919839 | 16 | 56859216 | C | T | 0.1022 |
| rs45474499 | 16 | 66914492 | T | C | 0.3433 |
| rs7185555 | 16 | 69131281 | G | C | 0.1537 |
| rs3751765 | 16 | 77910418 | A | G | 0.0807 |
| rs8056413 | 16 | 84082650 | G | T | 0.0902 |
| rs7215084 | 17 | 3880148 | T | C | 0.1196 |
| rs35552228 | 17 | 4626318 | G | T | 0.0966 |
| rs8069739 | 17 | 8078765 | C | T | 0.1082 |
| rs940851 | 17 | 8929764 | C | T | 0.0825 |
| rs9894621 | 17 | 12000584 | G | A | 0.0752 |
| rs1465249 | 17 | 12639294 | T | C | 0.0806 |
| rs230919 | 17 | 15115717 | C | T | 0.0918 |
| rs9893005 | 17 | 16225506 | G | C | 0.1058 |
| rs12938803 | 17 | 19204432 | T | C | 0.1482 |
| rs11080134 | 17 | 29161503 | G | A | 0.0974 |
| rs11077961 | 17 | 81012749 | A | G | 0.0946 |
| rs334414 | 18 | 7124612 | C | T | 0.0914 |
| rs11665020 | 18 | 10879503 | G | C | 0.1371 |
| rs4800420 | 18 | 20158965 | A | G | 0.1114 |
| rs12605662 | 18 | 25751666 | A | G | 0.0734 |
| rs10164193 | 18 | 31161426 | G | T | 0.182 |
| rs34163044 | 18 | 51851616 | A | C | 0.1488 |
| rs72930904 | 18 | 52607301 | C | T | 0.1139 |
| rs12605156 | 18 | 53498114 | A | T | 0.1277 |
| rs7235890 | 18 | 55732115 | G | T | 0.1222 |
| rs28625082 | 19 | 3108544 | A | G | 0.0891 |
| rs2009733 | 19 | 8398714 | A | G | 0.1054 |
| rs66502847 | 19 | 12615646 | A | G | 0.0885 |
| rs117533700 | 19 | 13176728 | C | T | 0.2474 |
| rs3745318 | 19 | 16436262 | T | C | 0.1213 |
| rs1077795 | 19 | 17222584 | A | G | 0.1764 |
| rs6511291 | 19 | 21950402 | C | T | 0.1002 |
| rs1821295 | 19 | 32590773 | C | T | 0.1347 |
| rs12983238 | 19 | 39438532 | G | A | 0.1095 |
| rs2125578 | 19 | 44746657 | C | T | 0.0779 |
| rs55706574 | 19 | 47556180 | G | A | 0.1008 |
| rs1232482 | 20 | 11886643 | C | T | 0.1155 |
| rs2618647 | 20 | 17882452 | G | A | 0.0983 |
| rs4462845 | 20 | 21498864 | G | C | 0.0902 |
| rs13042148 | 20 | 32298286 | C | T | 0.1543 |
| rs4810332 | 20 | 40268334 | T | A | 0.1562 |
| rs2425757 | 20 | 44792921 | C | T | 0.092 |
| rs237485 | 20 | 48004238 | A | G | 0.089 |
| rs113824370 | 20 | 49182690 | C | A | 0.2458 |
| rs35213536 | 20 | 62694319 | T | G | 0.1884 |
| rs2834253 | 21 | 35093213 | C | T | 0.0842 |
| rs8128234 | 21 | 36470865 | T | C | 0.1071 |
| rs62229372 | 21 | 37692507 | T | C | 0.1446 |
| rs1571737 | 21 | 43530671 | T | C | 0.119 |
| rs5992929 | 22 | 18451977 | T | C | 0.1424 |
| rs9609429 | 22 | 32517431 | T | C | 0.1094 |
| rs240069 | 22 | 34083187 | G | C | 0.1236 |
| rs2012831 | 22 | 36837260 | A | G | 0.0781 |
| rs9627391 | 22 | 46447097 | T | C | 0.0883 |

